# Integrated genomic analysis of small intestinal neuroendocrine tumors provides further insights into molecular subgroups and identifies putative driver genes

**DOI:** 10.1101/2025.03.10.25323484

**Authors:** Samuel Backman, Elham Barazeghi, Olov Norlén, Per Hellman, Peter Stålberg

## Abstract

Small intestinal neuroendocrine tumors (SI-NETs) are the most common tumor of the small intestine. Despite typically being metastatic at the time of diagnosis, the tumors are usually indolent and associated with a relatively long survival. Nevertheless, some patients fare worse and succumb to the disease soon after diagnosis despite similar stage and grade. To date, molecular prognostic markers are lacking, and our understanding of the disease biology is incomplete. Only *CDKN1B* is recurrently mutated in a subset of cases. Copy number alterations, including loss of chromosome 18, are common. However, the oncogenic mediators of these alterations remain unknown. The aim of this study was to identify molecular derangements in SI-NETs and identify potential prognostic markers. Forty tumor samples (including ten paired metastatic and primary tumors) from thirty patients with unusually long or unusually short survival after diagnosis were included for whole genome sequencing. Twenty of the samples were also included for RNA Sequencing. We validate known gene mutations and copy number alterations, as well as previously described molecular subgroups. We identify potential prognostic markers based on gene expression. Finally, we identify genes whose expression is affected by the most common copy number alterations in these tumors.

## Introduction

Neuroendocrine tumors of the small intestine (SI-NETs) are presumed to develop from the enterochromaffin cells that are scattered in the intestinal mucosa (Kulke and Mayer, 1999). The tumor cells often secrete a number of endocrine mediators including serotonin and tachykinins which, when they reach the peripheral circulation, cause the carcinoid syndrome: Flush, diarrhoea and sometimes fibrosis of the cardiac valves (Ito et al., 2018). The disease is relatively rare with an estimated incidence of approximately 1/100000/year (Dasari et al., 2017), which has been shown to increase over the past decades (Yao et al., 2008). Despite typically advanced stage at diagnosis the tumors are associated with a comparatively indolent disease course and a median overall survival of more than 8 years (Norlén et al., 2012). Nevertheless, some patients fare worse and succumb soon after diagnosis. Established favourable clinical prognostic factors include young age at diagnosis, absence of carcinoid syndrome, low grade and stage, absence of metastases to mesenteric/distant abdominal lymph nodes and liver, as well as absence of peritoneal carcinomatosis (Norlén et al., 2012). Several studies have attempted to identify molecular prognostic factors; downregulation of mir-375 (Arvidsson et al., 2018) in metastatic lesions, and amplification of chromosome 14 (Andersson et al., 2009) have both been associated with poor outcome. Based on DNA methylation and copy number profiles, three subgroups have been defined. A subgroup characterized by isolated loss of chromosome 18 has been demonstrated to have better prognosis than the other groups, characterized by multiple copy number alterations and no copy number alterations, respectively (Karpathakis et al., 2016). However, none of these biomarkers have reached clinical use.

Despite several large-scale efforts, the genetic drivers of SI-NETs remain largely obscure. Mutations in *CDKN1B* are found in ∼9% of cases (Francis et al., 2013, Crona et al., 2015), while loss of chromosome 18 has been pinpointed as the most common aberration and is found in 50-80% of cases (Löllgen et al., 2001, Kytölä et al., 2001, Karpathakis et al., 2016). Additional common chromosomal aberrations include gain of chromosomes 4, 5 and 20, as well as loss of chromosomes 9, 11 and 16. No further recurrently mutated genes have been identified in the two published exome sequencing studies (Banck et al., 2013, Francis et al., 2013). The mechanisms through which these copy number alterations drive tumor development have not been clearly identified.

In an attempt to shed light on the genetic drivers of the disease and identify potential markers of poor prognosis we performed whole genome sequencing of a cohort of 30 patients selected for very long or short disease-specific survival (14 and 16 cases in each group, respectively). For five patients in each group we sequenced a primary tumor-metastasis pair. For a subset of twenty samples, we generated both DNA sequencing data and transcriptome data, allowing us to identify genes directly affected by copy number alterations.

## Methods

### Ethics

Ethical permission for the present study was obtained from the regional ethical review board (Regionala etikprövningsnämnden i Uppsala, decision no 2011/375 with amendments). Patients provided consent biobanking of specimens and inclusion in genetic studies.

### Tissue samples and DNA preparation

All included patients underwent resection for SI-NET at the Endocrine Surgery unit at Uppsala University Hospital between 1988 and 2009. Following tumor resection, a sample was taken from the tumor and snap-frozen in liquid nitrogen. Samples were stored at -70 C.

Cryosections (∼20 sections of 20 um thickness) were obtained using a Leica cryostat. DNA was extracted from cryosections using Qiagen DNEasy Blood&Tissue kit (Qiagen, Hilden, Germany) according to the manufacturer’s instructions. For each sample two 6um cryosections (one before and one after the sections used for DNA extraction) were stained using Hematoxylin and Eosin to ensure sufficient tumor purity. Whenever deemed possible, samples were macrodissected to minimize the presence of normal tissue in the samples. Clinical data were extracted from patient records as previously described (Norlén et al., 2012).

### RNA preparation

Tissue sections were obtained as described above. RNA was extracted using the Qiagen RNEasy Mini kit. The RNA was DNAse-treated with Invitrogen Turbo-Free DNase kit.

### Whole genome sequencing

Sequencing libraries were prepared from 1 microgram of DNA using the TruSeq PCR-free DNA sample preparation kit (cat# FC-121-3001/3002, Illumina Inc.). An insert size of 350bp was used. Paired-end 150bp reads were sequenced on an Illumina HiSeqX. A minimum mean coverage of 59X was obtained.

### RNA-Seq

Libraries were prepared from 1 µg total RNA using the TruSeq stranded total RNA kit and RiboZero gold kit according to the manufacturer’s instructions. Paired-end sequencing with 125 bp read length was performed on five lanes of an Illumina HiSeq2500 sequenced using v4 chemistry in the high throughput mode.

### Expression analyses

FASTQ files resulting from the RNA-Seq libraries were analysed using the nf-core/rnaseq pipeline (Ewels et al., 2020) v 3.9. Quality control was carried out using FastQC v. 0.11.9. Adapter sequences and poor quality bases were trimmed using TrimGalore! v 0.6.7. Subsequently, the remaining reads were aligned to the genome (ENSEMBL version 112) using STAR (Dobin et al., 2013) v 2.6.1d. Transcript quantification was performed using Salmon (Patro et al., 2017) v 1.5.2. Read counts were imported to R using tximport (Soneson et al., 2015). Normalization as well as differential expression analyses were carried out using DeSeq2 (Love et al., 2014). Prior to differential expression analysis and clustering, a filter was employed to only include features with at least five counts in at least five samples. For the PCA and clustering analyses, the data were transformed using the vst function, and for the clustering also scaled using the R scale function. Consensus clustering was carried out using the ConsensusClusterPlus R package on the top 30% most variably expressed genes, using 10000 repetitions, pItem=0.8 and pFeature=0.8.

### Gene ontology enrichment analysis

Gene ontology enrichment analyses were performed using the enrichGO function from the R package clusterProfiler (Wu et al., 2021). Up- and downregulated genes (absolute fold change >0.5) were considered separately.

### Variant calling

Variants were called using FreeBayes (Garrison and Marth, 2012). Variants with quality scores of 20 and less, or read depth of 9 or less were filtered out. Vcfanno (Pedersen et al., 2016) was used to annotate variants with allele count and allele frequency from the gnomAD (Karczewski et al., 2020) v2.0.2 exome and genome datasets as well as the SweFreq dataset (Ameur et al., 2017). Variants with an allele frequency of 0.0001 or higher in either of the GnomAD genome or exome datasets as well as variants with an allele count of 1 or higher in SweFreq were excluded. Variants were annotated using snpEff and VEP using the nf-core/sarek pipeline (Ewels et al., 2020). The COSMIC Cancer Gene Census was used as a reference to identify variants in known cancer genes. Remaining variants were manually curated; variants in long and highly polymorphic regions (e.g. trinucleotide repeats and poly-T regions) were excluded, as were variants not found in gnomAD/SweFreq but reported as common in dbSNP, and variants consistently predicted to be benign by SIFT and Polyphen.

### Copy number analysis

Copy number analysis of WGS data was performed using Control-FREEC (v 11.6) (Boeva et al., 2012) with. GC correction was performed using the reference genome sequence. Correction for normal cell contamination was employed, and the contribution of normal cells was automatically inferred. Visualisation was carried out using the makeGraph.R script. Very small CNVs, and greatly variable copy numbers located in telomeric and centromeric regions were not considered in downstream analyses as they were deemed likely results of germline variation and read mapping issues, respectively. For the clustering analysis, chromosome arms were determined to be lost or gained if 80% of the informative windows on the chromosome arm was affected. A sample by chromosome arm matrix was created, and hierarchical clustering and visualization was done using the R heatmap.2 function with the Manhattan metric as the distance metric.

## Results

### Cohort overview

Forty samples from 30 patients were included (Table 1). The mean age at diagnosis was 58 (range 25-85) and 14 of the patients were female. All had stage 4 disease, and all patients for whom Ki67 staining was available in the pathology report had Grade 1-2 disease. The fourteen patients in the short survival group had a mean survival (from diagnosis to death) of 1 year (range 0-2 years) while those in the long survival group had a mean survival of 18 years (range 10-30 years). The two groups differed in age at diagnosis with the long survival group being younger (median 47 vs 68.5, p=0.0001 by Mann-Whitney U test).

**Table 1.** Overview of the studied cohort.

| Patient no | Primary WGS | Metastasis WSG | RNA-Seq | Age at diagnosis | Survival group | Stage | WHO grade |
| --- | --- | --- | --- | --- | --- | --- | --- |
| 1 |  | Included | Metastasis | 76-80 | Short | IV | 2 |
| 2 |  | Included |  | 56-60 | Short | IV | N/A |
| 3 | Included | Included | Metastasis | 56-60 | Short | IV | N/A |
| 4 |  | Included |  | 61-65 | Short | IV | 1 |
| 5 | Included | Included | Primary | 66-70 | Short | IV | N/A |
| 6 | Included |  |  | 66-70 | Short | IV | N/A |
| 7 | Included | Included | Primary | 71-75 | Short | IV | N/A |
| 8 | Included |  |  | 66-70 | Short | IV | N/A |
| 9 |  | Included | Metastasis | 66-70 | Short | IV | 2 |
| 10 | Included |  | Primary | 71-75 | Short | IV | N/A |
| 11 | Included | Included | Metastasis | 81-85 | Short | IV | 1 |
| 12 | Included |  | Primary | 61-65 | Short | IV | N/A |
| 13 | Included |  | Primary | 66-70 | Short | IV | 1 |
| 14 | Included | Included | Metastasis | 61-65 | Short | IV | N/A |
| 15 |  | Included |  | 66-70 | Long | IV | N/A |
| 16 | Included |  | Primary | 36-40 | Long | IV | N/A |
| 17 | Included | Included |  | 71-75 | Long | IV | N/A |
| 18 |  | Included | Metastasis | 61-65 | Long | IV | N/A |
| 19 |  | Included | Metastasis | 56-60 | Long | IV | 2 |
| 20 | Included |  | Primary | 41-45 | Long | IV | 1 |
| 21 | Included |  |  | 35-40 | Long | IV | 1 |
| 22 | Included | Included | Metastasis | 46-50 | Long | IV | 1 |
| 23 |  | Included | Metastasis | 46-50 | Long | IV | N/A |
| 24 | Included | Included | Metastasis | 21-25 | Long | IV | N/A |
| 25 | Included |  | Primary | 46-50 | Long | IV | 1 |
| 26 | Included | Included | Metastasis | 46-50 | Long | IV | 1 |
| 27 |  | Included |  | 46-50 | Long | IV | N/A |
| 28 | Included |  |  | 41-45 | Long | IV | N/A |
| 29 |  | Included |  | 41-45 | Long | IV | N/A |
| 30 | Included | Included | Metastasis | 56-60 | Long | IV | N/A |

### Copy number aberrations

Copy number aberrations were found in 37 of the samples. The most common aberration was loss of chromosome 18, which was found in 19 of the samples with an additional sample with a small deletion. Additional recurrent alterations included loss of 1p, gain of chromosomes 4 and 5, loss of parts of chromosome 11q, gain of chromosome 14 and gain of chromosome 20 (Supplementary figures).

In order to recreate the previously published biological subgroups (Karpathakis et al., 2016) we performed hierarchical clustering based on chromosome arm level copy number events (Fig 1A). We identified a subgroup characterized by chromosome 18 deletion (Loss18), a subgroup with numerous CNVs (MultiCNV), and a subgroup with few alterations (Silent), establishing the validity of these groups. Notably, in 4/10 cases with paired primary and metastatic tumors, the paired tumors belonged to different copy number clusters. While the Loss18 group has previously been associated with good outcome, samples from the two survival groups in our cohort were evenly distributed across the clusters and did not show significant differences in the number of arm level copy number alterations (Fig 1B).

**Figure 1.**
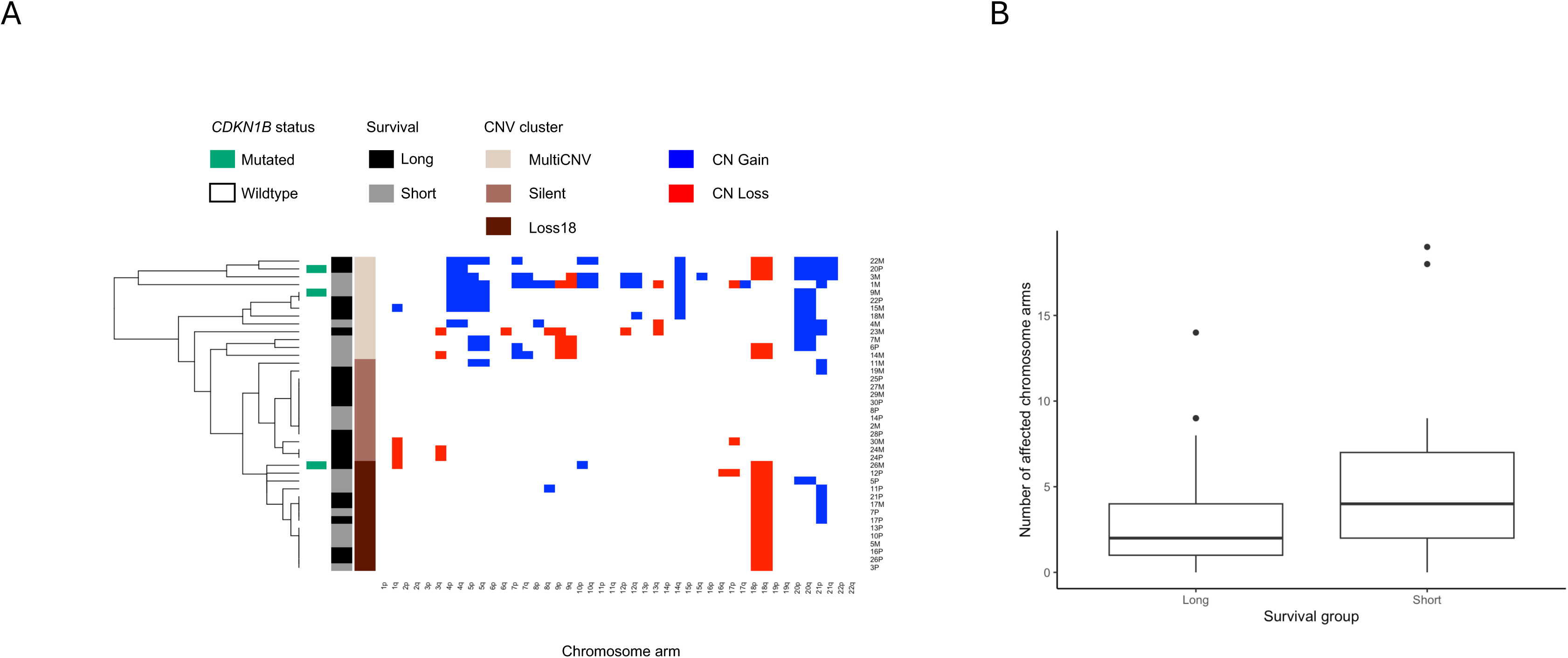
A) Hierarchical clustering based on arm-level somatic copy number alterations identifies known subgroups: A subgroup characterized by multiple CNVs (MultiCNV), a group with no or few alterations (Silent) and a group characterized by isolated loss of chromosome 18 (Loss18). B) The number of arm-level copy number alterations does not significantly differ between the groups with long and short survival.

### Protein altering mutations

First, we identified mutations in the only known SI-NET gene, *CDKN1B*. Three samples had mutations in this gene: 26M carried a p.Val90fs mutation, which was not called in the paired primary tumor 26P. 20P carried a p.Glu54* mutation, and 9M a p.Glu40* mutation. The fraction of samples with *CDKN1B* mutations was similar to that previously described. Two of the *CDKN1B*-mutated samples belonged to the MultiCNV-group, and one to the Loss18 group. These mutations were previously detected by Sanger sequencing and reported in Crona et al 2015.

We next expanded the analysis to selected known neuroendocrine cancer genes. The paired primary and metastatic tumors 14P and 14M carried a p.Leu212fs mutation in *NF1*, which was coupled with a focal deletion on chromosome 17 suggestive of biallelic inactivation. One additional sample carried an in-frame deletion in *NF1,* which was not associated with deletion of the other allele. RNA-Sequencing data revealed a higher than average expression of *NF1* in this sample, which can be contrasted with 14M which had the lowest expression of *NF1* of all studied samples.

2M carried a p.Gln62* frameshift mutation in *MAX*. Two additional samples from one patient, 7P and 7M, carried a missense variant of uncertain significance in the same gene which was predicted to be benign by both Polyphen and SIFT.

### Transcriptome clustering

We performed unsupervised clustering of the RNA-sequencing samples based on the top 10% of genes with the most variable expression across the cohort (Figure 2A). While the predefined clinical groups with long and short survival were dispersed throughout the dendrogram, a distinct cluster was noted comprised of the primary tumor samples from the Loss18 copy number group. Similar results were obtained using consensus clustering where all Loss18 samples clustered together (Figure 2B). On the PCA plot separation of the primary tumor and the metastasis samples was observed, as well as some separation of the copy number subgroups (Figure 2C-D).

**Figure 2.**
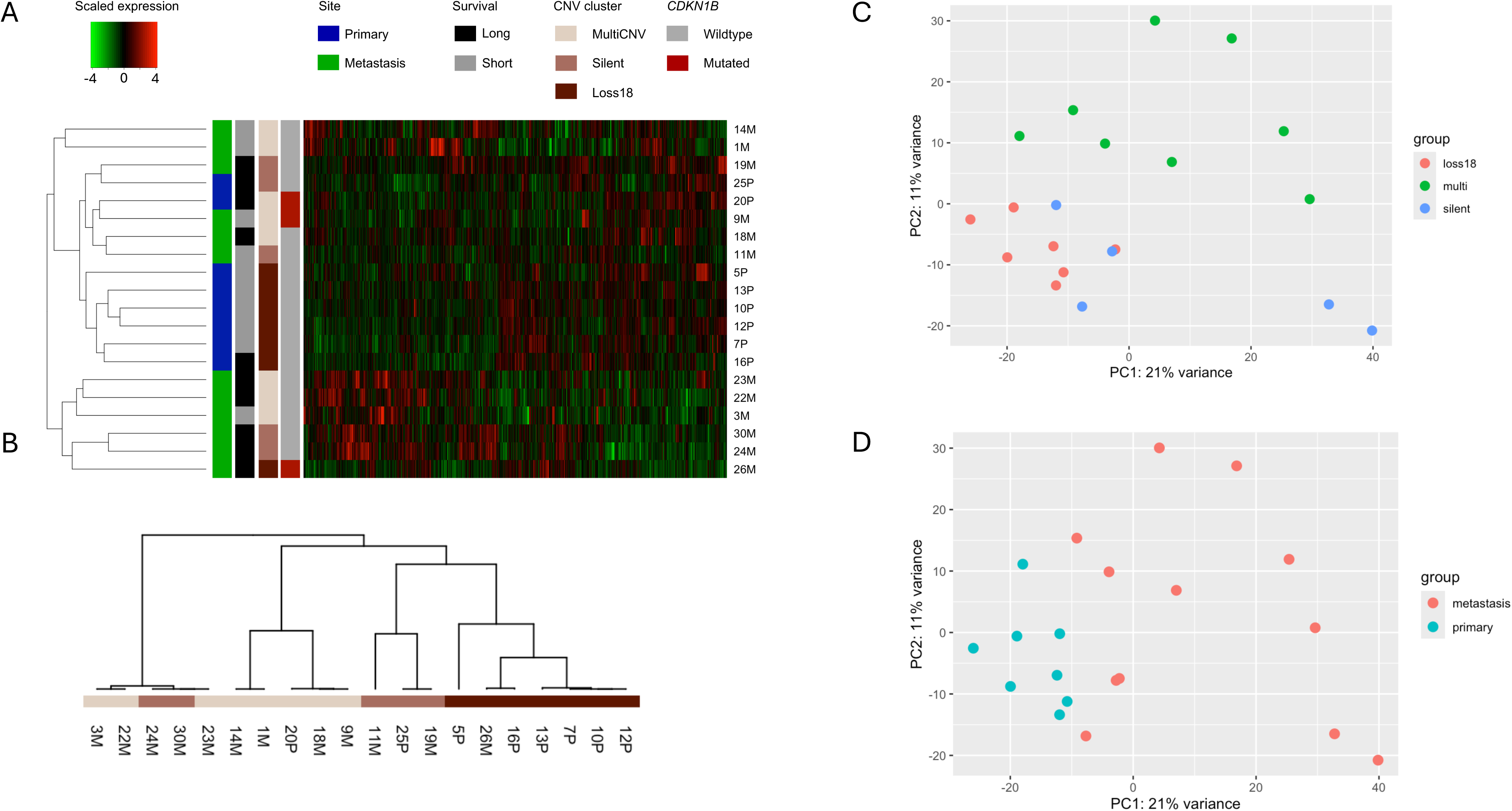
A) Hierarchical clustering based on the top 10% most variably expressed genes in the cohort clusters all primary tumor samples from the Loss18 group together. B) Consensus Clustering results C-D) PCA separates primary and metastatic tumors, and the different subgroups

In order to further understand the biological differences between these groups of tumors, we performed differential expression analyses of the tumors in each group against all other tumors (Fig 3A-C). The Loss18 had 791 transcriptome-wide significantly differentially expressed genes (Supplementary table 1). Gene ontology analyses revealed that the differentially expressed genes were enriched for involvement in developmental processes and neuronal apoptosis (Fig 3D-E). The MultiCNV had 1278 differentially expressed genes (Supplementary table 2). No gene ontologies were significantly enriched among either the upregulated or downregulated genes. The Silent group had 433 differentially expressed genes, (Supplementary table 3), and among the upregulated genes, “cilium movement involved in cell motility” was the top overrepresented Gene ontology (Figure 3F. Notably, the MultiCNV group overexpressed *EZH2*, previously demonstrated to have a therapeutically targetable oncogenic role in SI-NETs (Barazeghi et al., 2021).

**Figure 3.**
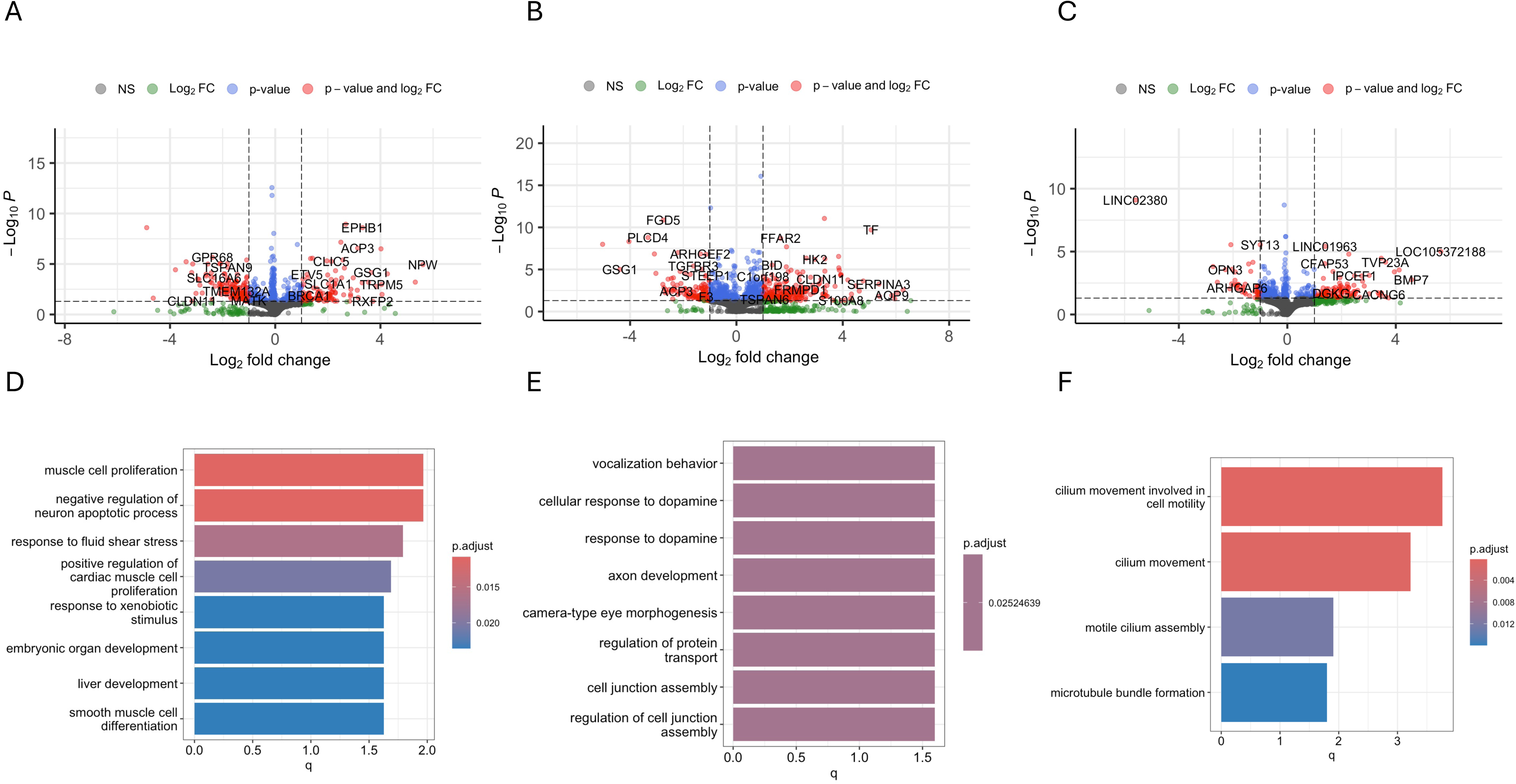
Volcano plots of genes differentially expressed in the A) Loss18 B) MultiCNV and C) Silent groups. D) Top gene ontologies enriched among genes downregulated in the Loss18 group E) Top gene ontologies enriched among genes upregulated in the Loss18 F) Top gene ontologies among genes upregulated in the silent group

### Gene expression differences between the long-survivors and the short-survivors

In order to identify potential biomarkers of survival a differential expression analysis comparing lesions from those with long survival with lesions from those with short survival was performed. A total of 126 genes were found differentially expressed (Supplementary table 4).

### Transcriptome of CDKN1B mutated tumors

Comparing three tumors with *CDKN1B* mutations to those without, 32 genes were differentially expressed (Supplementary table 5), although most with very small effect sizes after fold-change shrinkage.

### Gene dosage

As the mechanism through which chromosomal copy number alterations contributes to SI-NET development remains incompletely understood, we utilized our paired copy number and gene expression data to identify genes affected by gene dosage. In the RNA-Seq cohort, a total of 11 samples had loss of chromosome 18. The samples with loss of chromosome 18 had a total of 546 differentially expressed genes (Figure 4A, Supplementary Table 6). Of these, 17 genes were located in the previously described minimally overlapping region 18q22-18qter and had lower expression in the tumors with loss of the chromosome (Fig 4B). Across the entire chromosome, 123 genes were affected by gene dosage, including previously investigated candidate tumor suppressor gene *PTPRM* (Barazeghi et al., 2019).

**Figure 4.**
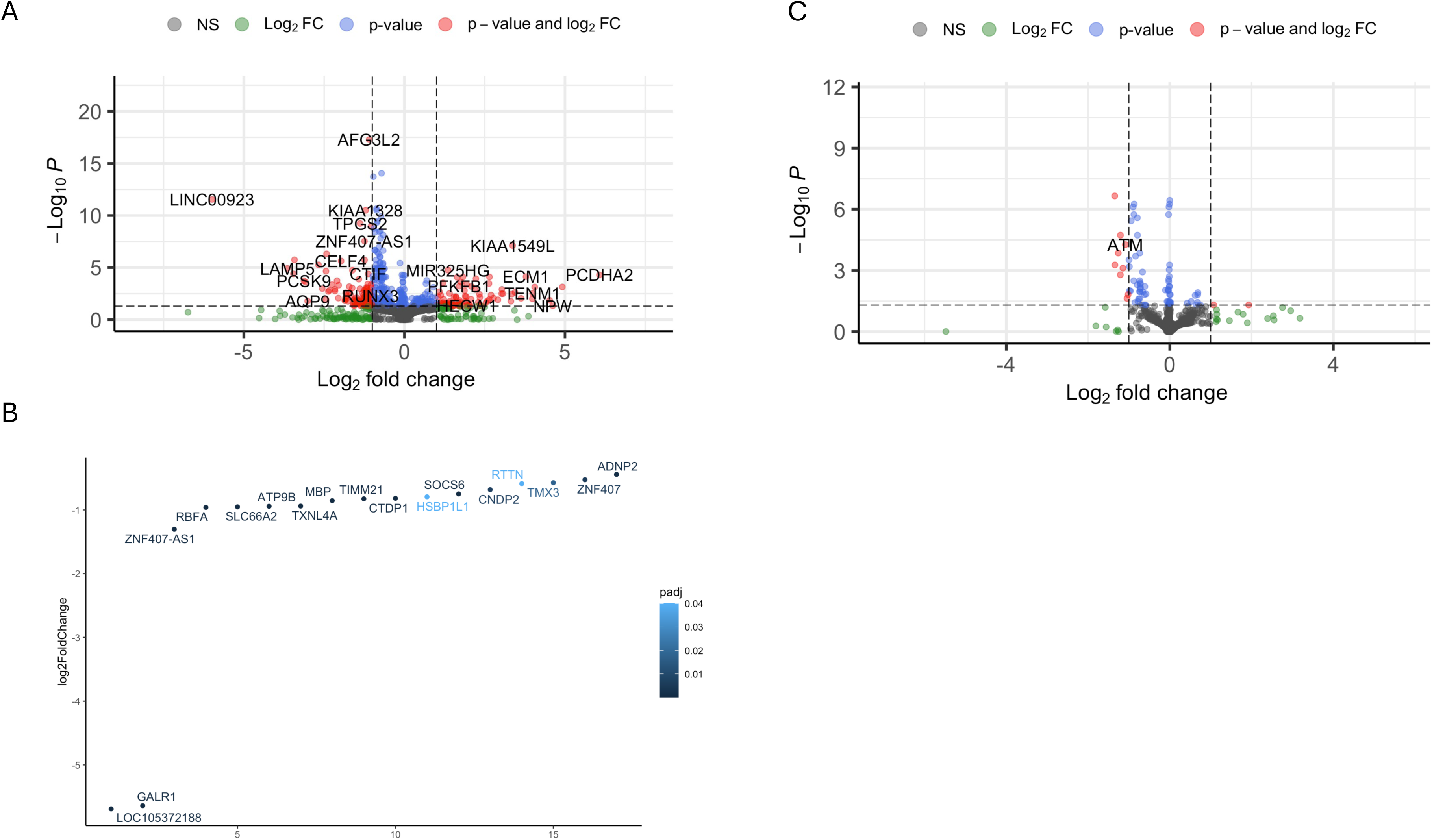
A) Gene expression differences between tumors with loss of chromosome 18 and those without. B) Genes on chromosome 18q22-18qter affected by gene dosage. C) *ATM* is affected by gene dosage in SI-NETs with deletions on chromosome 11

Similarly, out of 80 genes differentially expressed in tumors with loss of chromosome 11, 42 were located on chromosome 11 and 41 of these had lower expression in the tumors with losses, including established tumor suppressor gene *ATM* (Figure 4C, Supplementary table 7) and cell-adhesion molecule *NCAM1*. In several of the cases, the chromosome 11-deletion was focal, spanning only a region of the chromosome, overlapping the *ATM* locus (Supplementary figures).

## Discussion

In the present paper we have studied genetic variations, chromosomal aberrations and gene expression variation in small intestine neuroendocrine tumors. We identify genes that are differentially expressed between tumors from patients with long survival and patients with short survival. Moreover, we were able to identify and verify previously described molecular subgroups based on CNV data. Our findings suggests that the subgroup characterized by loss of chromosome 18 in the context of a relatively stable genome exhibits a distinct transcriptomic signature. In half of the cases where multiple tumors from the same patient were sequenced, the tumors belonged to different copy number clusters. This demonstrates the heterogenous biology of these tumors, which has previously been demonstrated both with regards to *CDKN1B* mutations and copy number profiles. SI-NETs are commonly multi-focal, and it has previously been shown that the tumors arise independently (Elias et al., 2021). It is thus likely that the tumors with discordant cluster assignment are not clonally related.

Limitations of the present study includes the lack of paired normal tissue for effective filtering of germline variants, and it cannot be excluded that variants originating in the germline remain after filtration against public databases. Consequently, the analyses focused on known cancer genes. Additionally, the included cohort is heterogeneous and includes metastatic as well as primary tumor material. Moreover, the two groups of long and short survivors, although similar in tumor characteristics such as grade and stage, differed significantly in age at diagnosis. The long-survivors were significantly younger when diagnosed, which has been previously described as a positive prognostic factor. Nevertheless, we identify a number of genes that were differentially expressed between the two groups, and which represent potential prognostic markers that warrant investigation in a separate validation cohort.

Devoid of recurrent somatic mutations, SI-NETs appear to be largely driven by copy number variation and epigenetic changes. Despite being the most prevalent genetic aberration in SI-NETs, the mechanism through which loss of chromosome 18 contributes to tumorigenesis remains unknown. Previous studies have exhaustively screened for mutations in the deleted regions (Delgado Verdugo et al., 2015) without findings. Our integrative analyses identify several genes located on chromosome 18 affected by gene dosage. One of these genes, *PTPRM*, has previously been shown to be epigenetically silenced in SI-NETs (Barazeghi et al., 2019), and its overexpression has been shown to inhibit cell proliferation in SI-NET cell lines, suggestive of a tumor suppressor role. While our descriptive data cannot conclusively establish a tumorigenic role for any of these genes, we believe that our results may guide future functional studies.

Loss of parts, or all of chromosome 11q is also a recurrent and relatively prevalent aberration in these tumors. Our integrative analysis of CNVs and gene expression suggests that this leads to a reduced expression of tumor suppressor gene *ATM* which encodes a serine/threonine kinase which is involved in the response to DNA damage and may activate cell cycle arrest and apoptosis (Cremona and Behrens, 2014). Germline mutations in *ATM* underlie Ataxia Telangiectasia, an autosomal recessive neurodegenerative disorder which is associated with an increased lifetime risk of cancer of approximately 30% (McKinnon, 2012), and somatic mutations of the gene have been reported (Ding et al., 2008).

In conclusion, we validate established genomic aberrations in SI-NETs. Additionally, through integration of genomic data with gene expression, we provide additional insight into previously established molecular subgroups. We also identify putative prognostic markers and genes which are affected by gene dosage in these tumors. Future studies in validation cohorts, and functional studies are required to corroborate these findings.

## Acknowledgments

The computations were enabled by resources provided by the National Academic Infrastructure for Supercomputing in Sweden (NAISS), partially funded by the Swedish Research Council through grant agreement no. 2022-06725.

## Author contributions

SB performed laboratory work and bioinformatics. PS and SB planned the study. PS, PH and ON provided study oversight. PS, PH, ON, EB and SB interpreted the data. SB drafted the manuscript. All authors critically reviewed and approved the final manuscript.

## Data availability

Code to reproduce key figures are available at sabackman/SINET2024: Code and data associated with Backman et al. 2024 (github.com). Requests for access to source data may be directed to the corresponding author and considered on an individual basis with regard to applicable legislation.

