## Supplementary figures for "Integrated genomic analysis of small intestinal neuroendocrine tumors provides further insights into molecular subgroups and identifies putative driver genes"

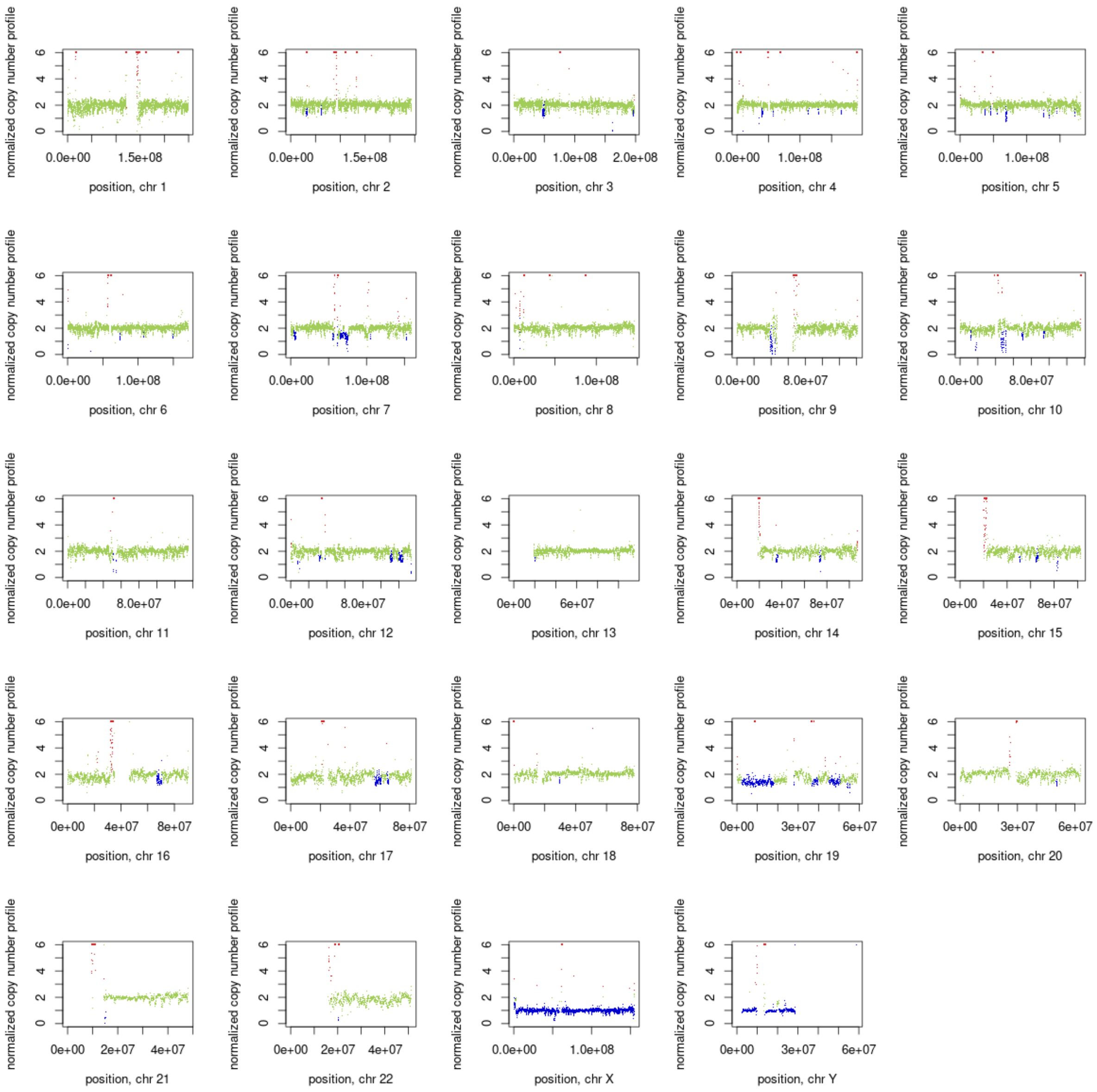
normalized copy number profile
position, chr 1
position, chr 2
position, chr 3
position, chr 4
position, chr 5
position, chr 6
position, chr 7
position, chr 8
position, chr 9
position, chr 10
position, chr 11
position, chr 12
position, chr 13
position, chr 14
position, chr 15
position, chr 16
position, chr 17
position, chr 18
position, chr 19
position, chr 20
position, chr 21
position, chr 22
position, chr X
position, chr Y

29M

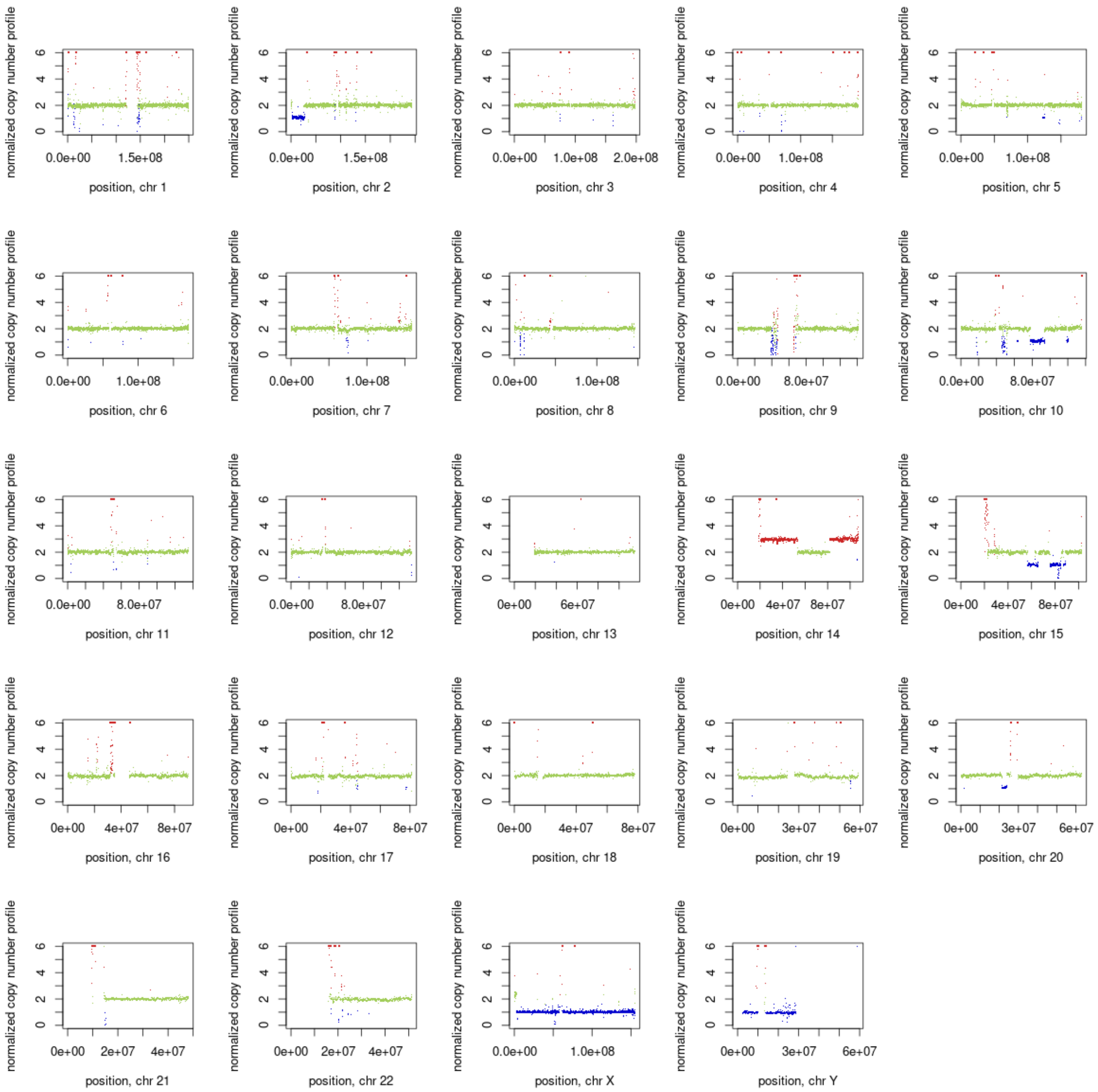
normalized copy number profile
0
2
4
6
0.0e+00 1.5e+08
position, chr 1
0.0e+00 1.5e+08
position, chr 2
0.0e+00 1.0e+08 2.0e+08
position, chr 3
0.0e+00 1.0e+08
position, chr 4
0.0e+00 1.0e+08
position, chr 5
0.0e+00 1.0e+08
position, chr 6
0.0e+00 1.0e+08
position, chr 7
0.0e+00 1.0e+08
position, chr 8
0.0e+00 8.0e+07
position, chr 9
0.0e+00 8.0e+07
position, chr 10
0.0e+00 8.0e+07
position, chr 11
0.0e+00 8.0e+07
position, chr 12
0e+00 6e+07
position, chr 13
0e+00 4e+07 8e+07
position, chr 14
0e+00 4e+07 8e+07
position, chr 15
0e+00 4e+07 8e+07
position, chr 16
0e+00 4e+07 8e+07
position, chr 17
0e+00 4e+07 8e+07
position, chr 18
0e+00 3e+07 6e+07
position, chr 19
0e+00 3e+07 6e+07
position, chr 20
0e+00 2e+07 4e+07
position, chr 21
0e+00 2e+07 4e+07
position, chr 22
0.0e+00 1.0e+08
position, chr X
0e+00 3e+07 6e+07
position, chr Y

2M

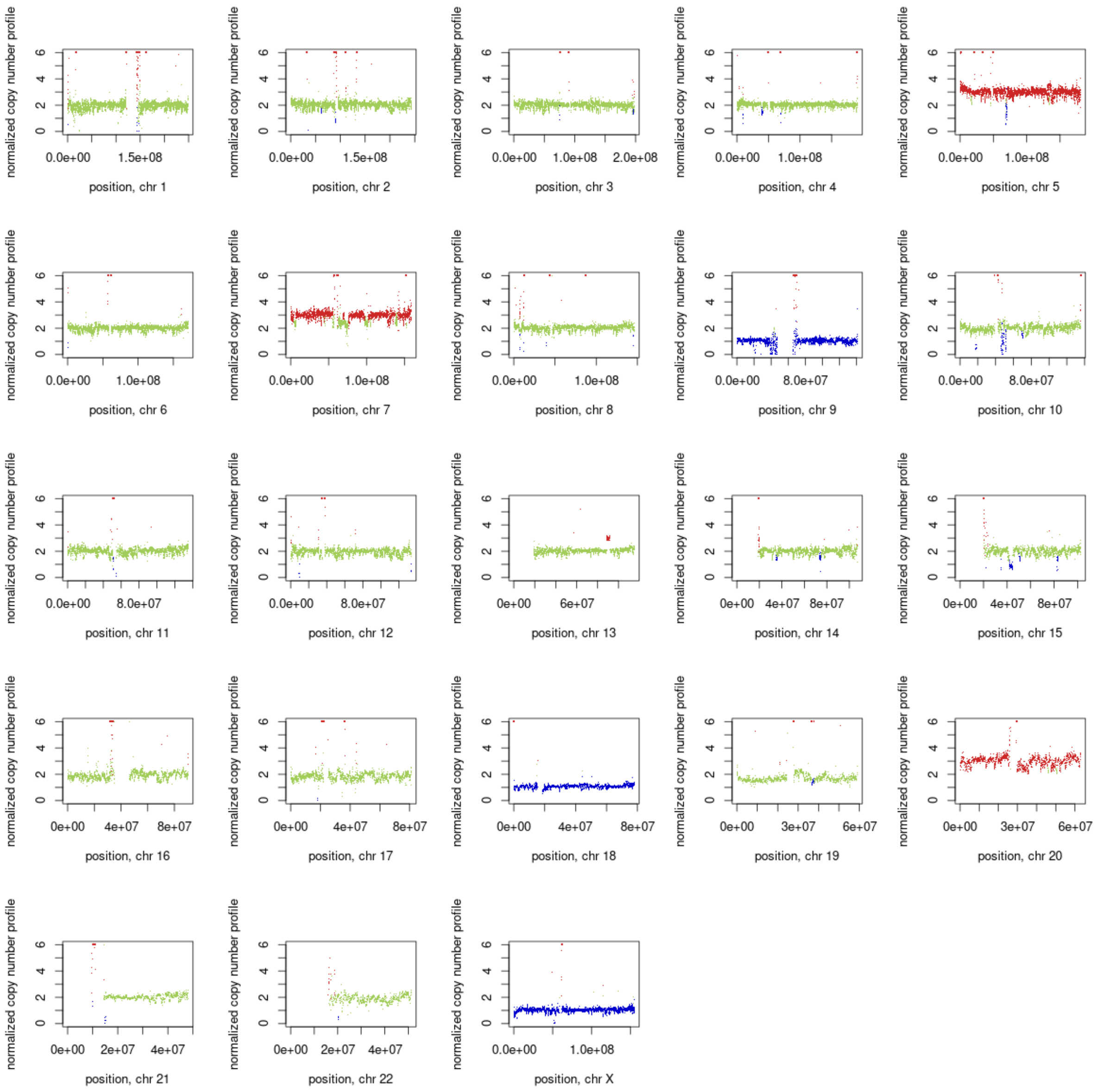
normalized copy number profile
position, chr 1
position, chr 2
position, chr 3
position, chr 4
position, chr 5
position, chr 6
position, chr 7
position, chr 8
position, chr 9
position, chr 10
position, chr 11
position, chr 12
position, chr 13
position, chr 14
position, chr 15
position, chr 16
position, chr 17
position, chr 18
position, chr 19
position, chr 20
position, chr 21
position, chr 22
position, chr X

6P

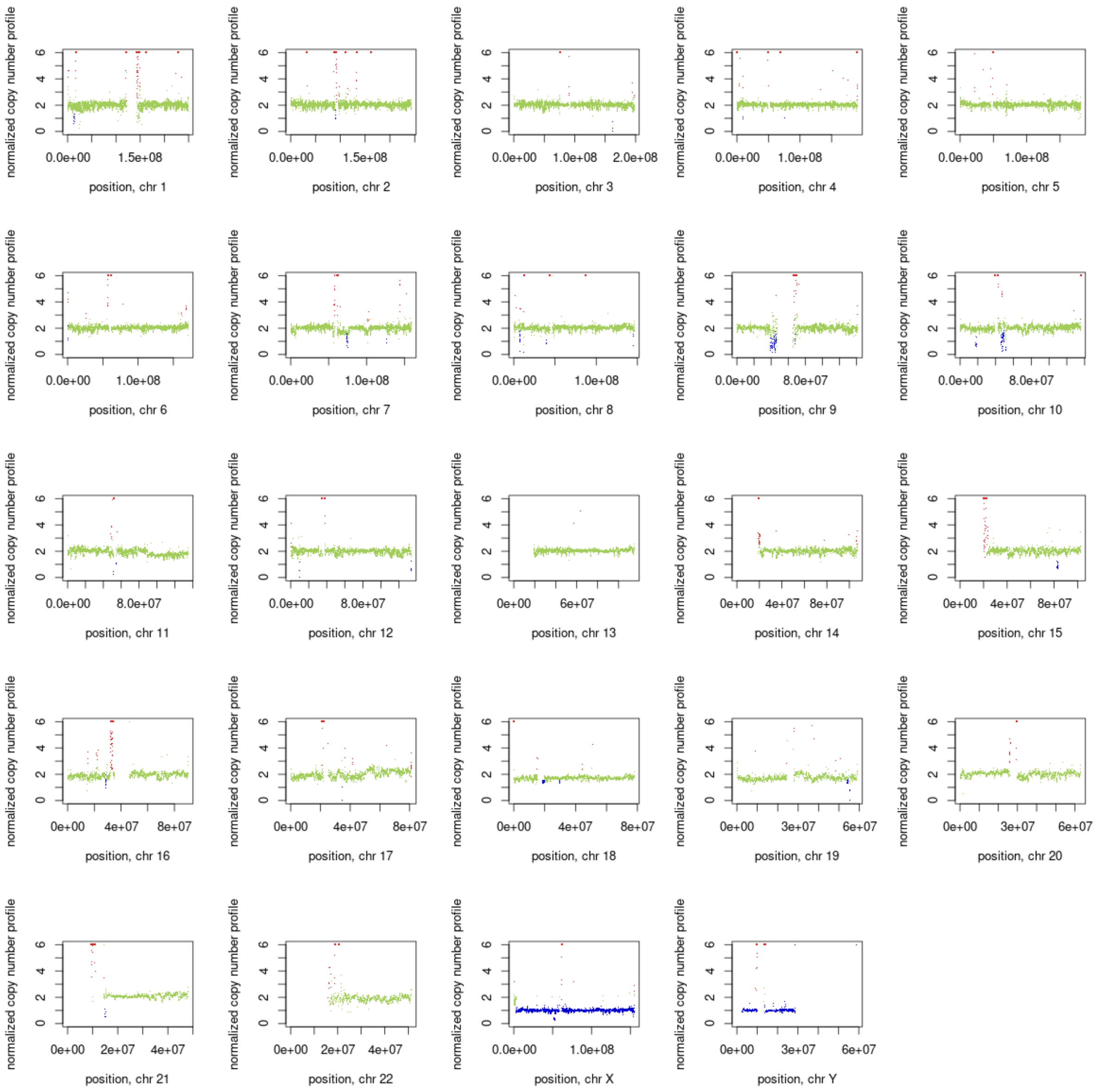
normalized copy number profile
0
2
4
6
0.0e+00
1.5e+08
position, chr 1
0.0e+00
1.5e+08
position, chr 2
0.0e+00
1.0e+08
2.0e+08
position, chr 3
0.0e+00
1.0e+08
position, chr 4
0.0e+00
1.0e+08
position, chr 5
0.0e+00
1.0e+08
position, chr 6
0.0e+00
1.0e+08
position, chr 7
0.0e+00
1.0e+08
position, chr 8
0.0e+00
8.0e+07
position, chr 9
0.0e+00
8.0e+07
position, chr 10
0.0e+00
8.0e+07
position, chr 11
0.0e+00
8.0e+07
position, chr 12
0e+00
6e+07
position, chr 13
0e+00
4e+07
8e+07
position, chr 14
0e+00
4e+07
8e+07
position, chr 15
0e+00
4e+07
8e+07
position, chr 16
0e+00
4e+07
8e+07
position, chr 17
0e+00
4e+07
8e+07
position, chr 18
0e+00
3e+07
6e+07
position, chr 19
0e+00
3e+07
6e+07
position, chr 20
0e+00
2e+07
4e+07
position, chr 21
0e+00
2e+07
4e+07
position, chr 22
0.0e+00
1.0e+08
position, chr X
0e+00
3e+07
6e+07
position, chr Y

27M

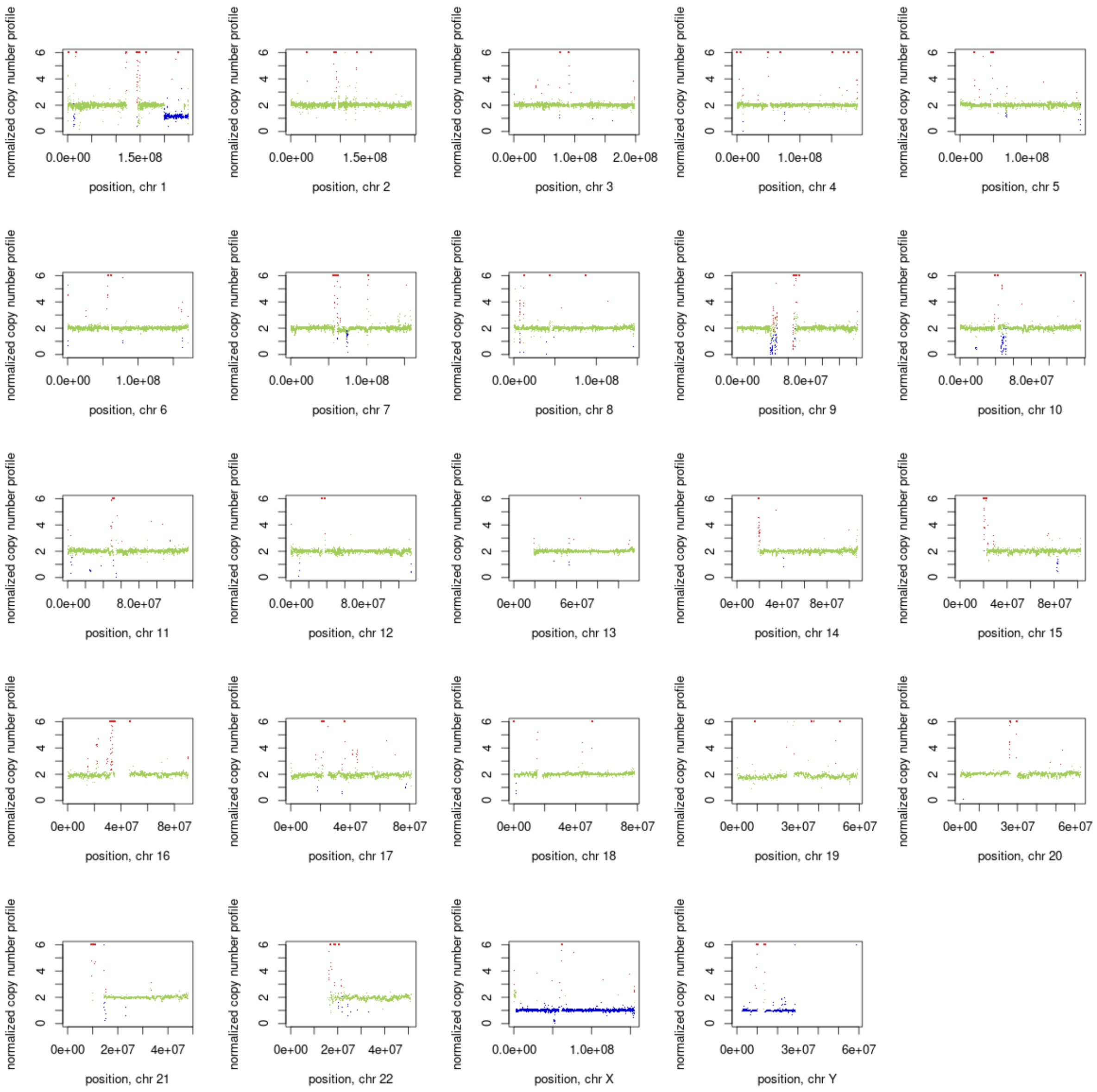
normalized copy number profile
0 2 4 6
0.0e+00 1.5e+08
position, chr 1
0.0e+00 1.5e+08
position, chr 2
0.0e+00 1.0e+08 2.0e+08
position, chr 3
0.0e+00 1.0e+08
position, chr 4
0.0e+00 1.0e+08
position, chr 5
0.0e+00 1.0e+08
position, chr 6
0.0e+00 1.0e+08
position, chr 7
0.0e+00 1.0e+08
position, chr 8
0.0e+00 8.0e+07
position, chr 9
0.0e+00 8.0e+07
position, chr 10
0.0e+00 8.0e+07
position, chr 11
0.0e+00 8.0e+07
position, chr 12
0e+00 6e+07
position, chr 13
0e+00 4e+07 8e+07
position, chr 14
0e+00 4e+07 8e+07
position, chr 15
0e+00 4e+07 8e+07
position, chr 16
0e+00 4e+07 8e+07
position, chr 17
0e+00 4e+07 8e+07
position, chr 18
0e+00 3e+07 6e+07
position, chr 19
0e+00 3e+07 6e+07
position, chr 20
0e+00 2e+07 4e+07
position, chr 21
0e+00 2e+07 4e+07
position, chr 22
0.0e+00 1.0e+08
position, chr X
0e+00 3e+07 6e+07
position, chr Y

30P

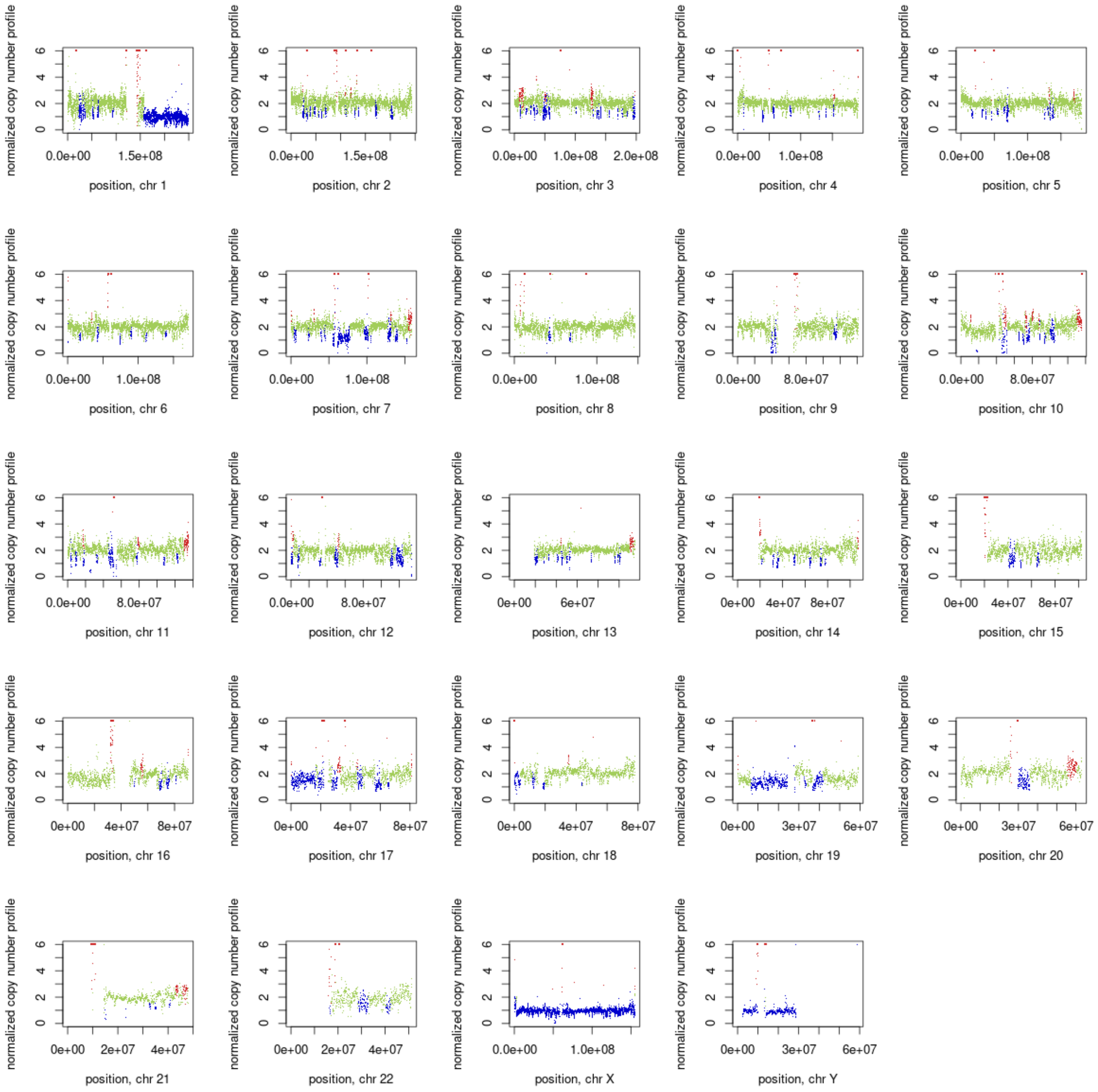
normalized copy number profile
position, chr 1
position, chr 2
position, chr 3
position, chr 4
position, chr 5
position, chr 6
position, chr 7
position, chr 8
position, chr 9
position, chr 10
position, chr 11
position, chr 12
position, chr 13
position, chr 14
position, chr 15
position, chr 16
position, chr 17
position, chr 18
position, chr 19
position, chr 20
position, chr 21
position, chr 22
position, chr X
position, chr Y

30M

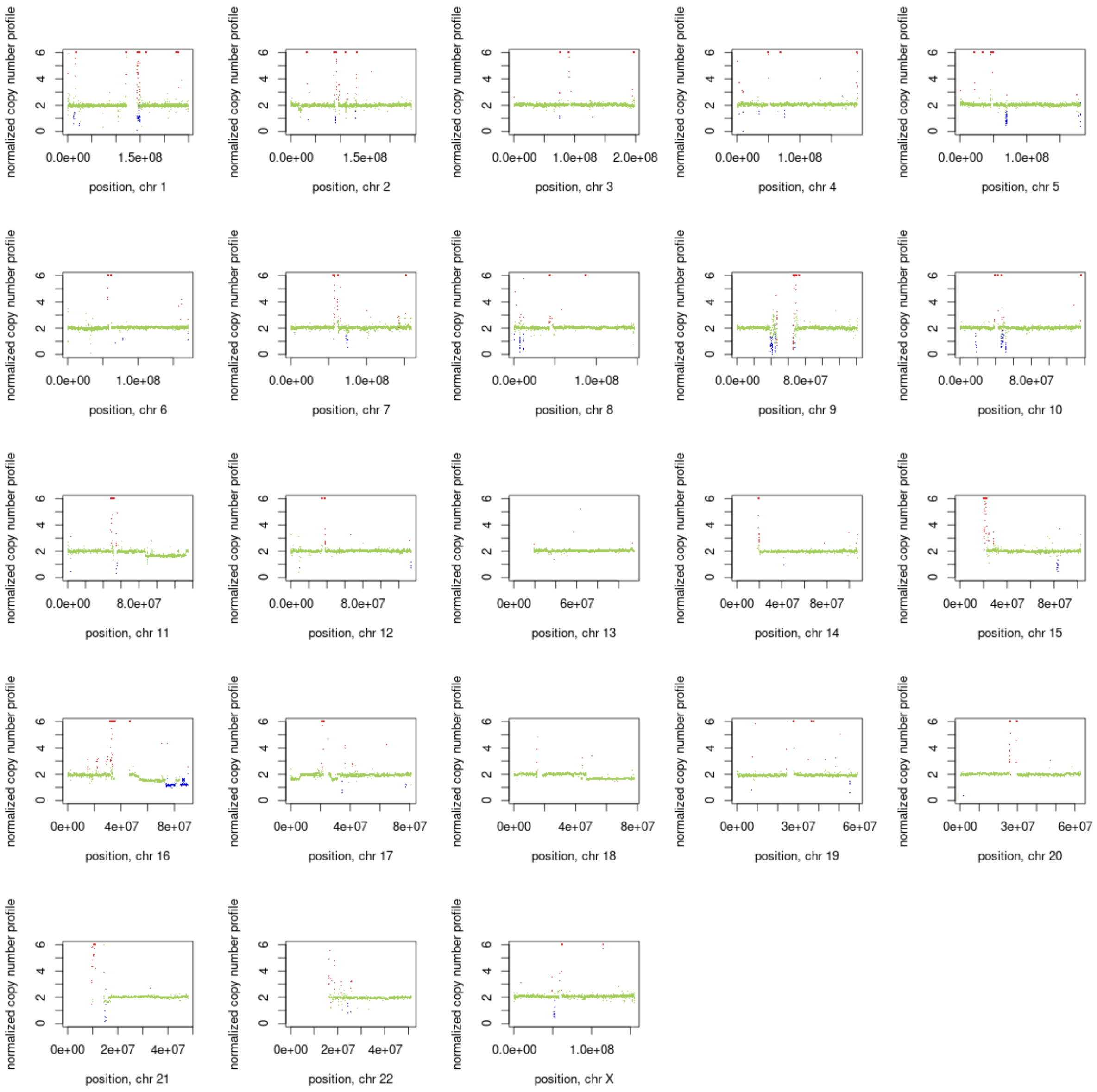
normalized copy number profile
0
2
4
6
0.0e+00
1.5e+08
position, chr 1
position, chr 2
1.0e+08
2.0e+08
position, chr 3
position, chr 4
position, chr 5
position, chr 6
position, chr 7
position, chr 8
8.0e+07
position, chr 9
position, chr 10
position, chr 11
position, chr 12
0e+00
6e+07
position, chr 13
4e+07
8e+07
position, chr 14
position, chr 15
position, chr 16
position, chr 17
position, chr 18
3e+07
6e+07
position, chr 19
position, chr 20
2e+07
position, chr 21
position, chr 22
position, chr X

28P

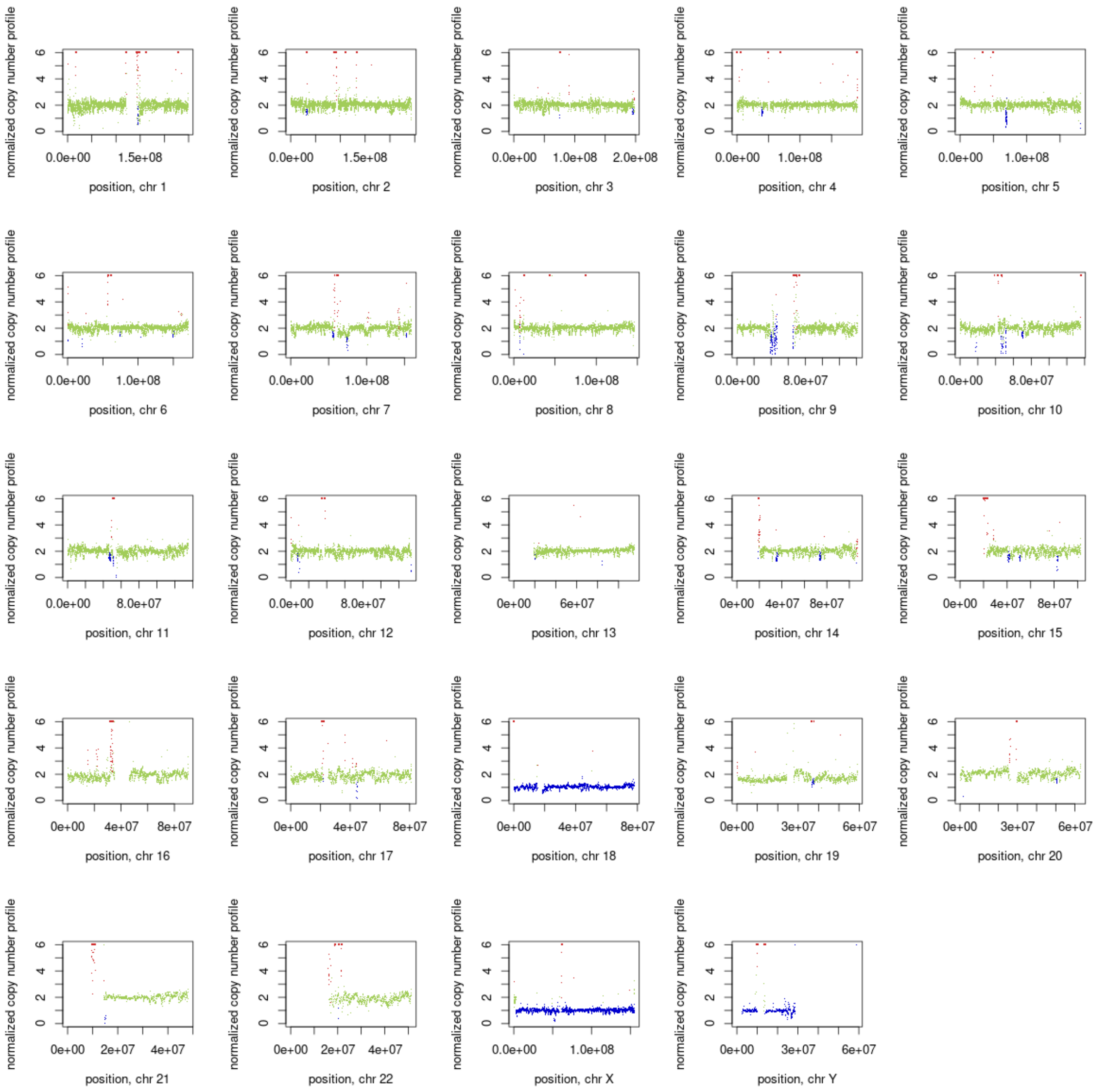
normalized copy number profile
position, chr 1
position, chr 2
position, chr 3
position, chr 4
position, chr 5
position, chr 6
position, chr 7
position, chr 8
position, chr 9
position, chr 10
position, chr 11
position, chr 12
position, chr 13
position, chr 14
position, chr 15
position, chr 16
position, chr 17
position, chr 18
position, chr 19
position, chr 20
position, chr 21
position, chr 22
position, chr X
position, chr Y

17P

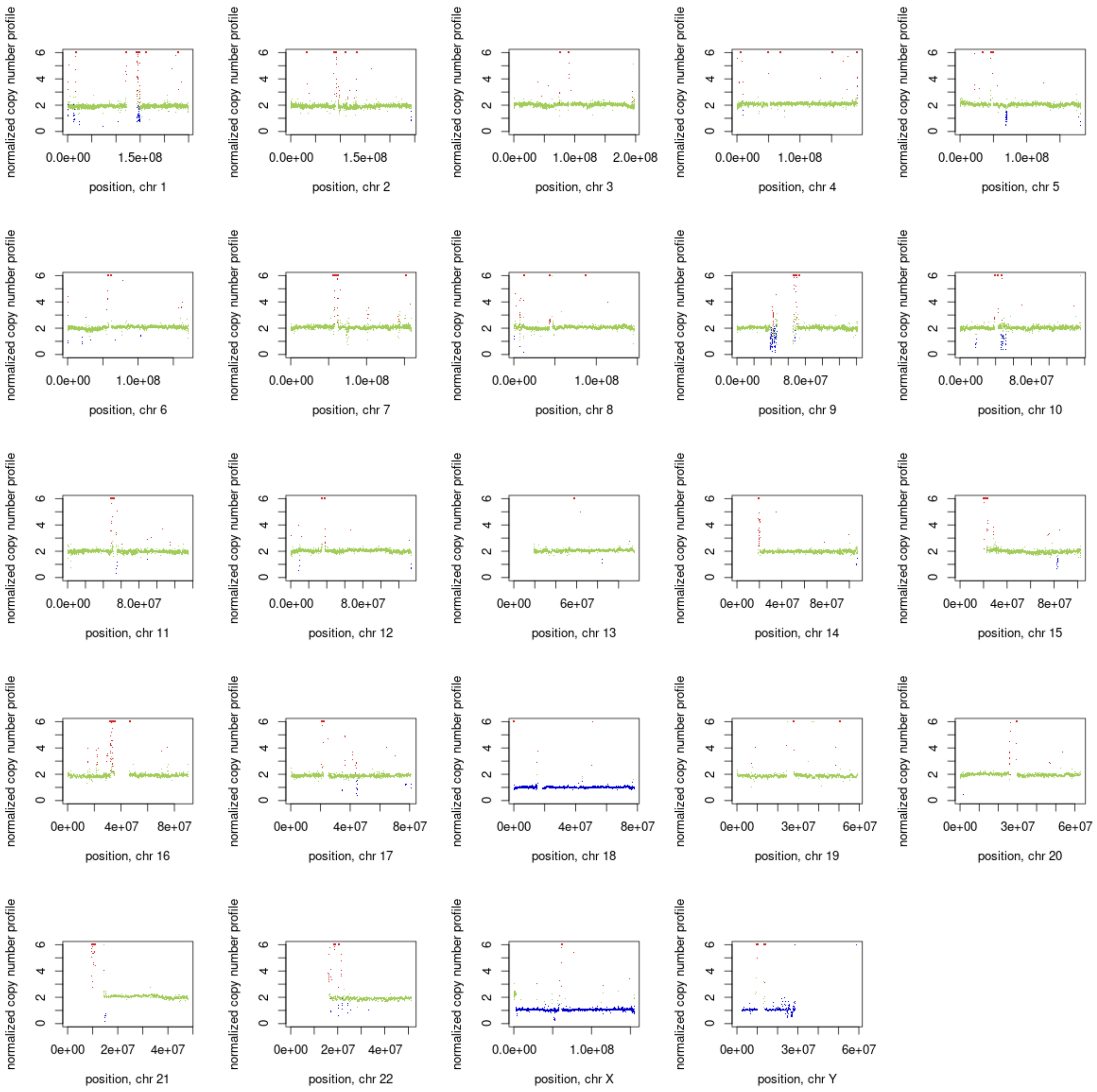
normalized copy number profile
position, chr 1
position, chr 2
position, chr 3
position, chr 4
position, chr 5
position, chr 6
position, chr 7
position, chr 8
position, chr 9
position, chr 10
position, chr 11
position, chr 12
position, chr 13
position, chr 14
position, chr 15
position, chr 16
position, chr 17
position, chr 18
position, chr 19
position, chr 20
position, chr 21
position, chr 22
position, chr X
position, chr Y

17M

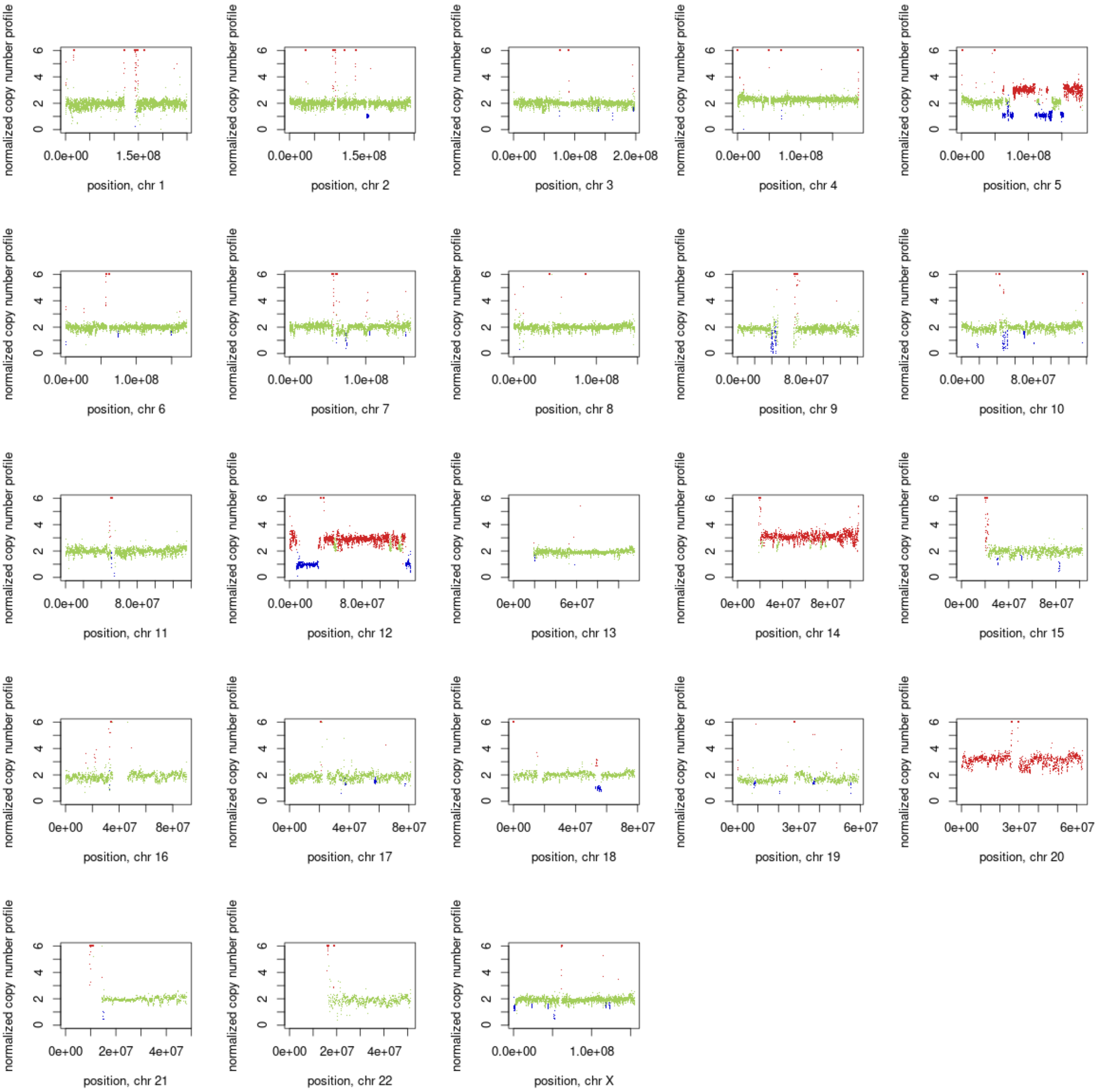
normalized copy number profile
0 2 4 6
0.0e+00 1.5e+08
position, chr 1
0.0e+00 1.5e+08
position, chr 2
0.0e+00 1.0e+08 2.0e+08
position, chr 3
0.0e+00 1.0e+08
position, chr 4
0.0e+00 1.0e+08
position, chr 5
0.0e+00 1.0e+08
position, chr 6
0.0e+00 1.0e+08
position, chr 7
0.0e+00 1.0e+08
position, chr 8
0.0e+00 8.0e+07
position, chr 9
0.0e+00 8.0e+07
position, chr 10
0.0e+00 8.0e+07
position, chr 11
0.0e+00 8.0e+07
position, chr 12
0e+00 6e+07
position, chr 13
0e+00 4e+07 8e+07
position, chr 14
0e+00 4e+07 8e+07
position, chr 15
0e+00 4e+07 8e+07
position, chr 16
0e+00 4e+07 8e+07
position, chr 17
0e+00 4e+07 8e+07
position, chr 18
0e+00 3e+07 6e+07
position, chr 19
0e+00 3e+07 6e+07
position, chr 20
0e+00 2e+07 4e+07
position, chr 21
0e+00 2e+07 4e+07
position, chr 22
0.0e+00 1.0e+08
position, chr X

18M

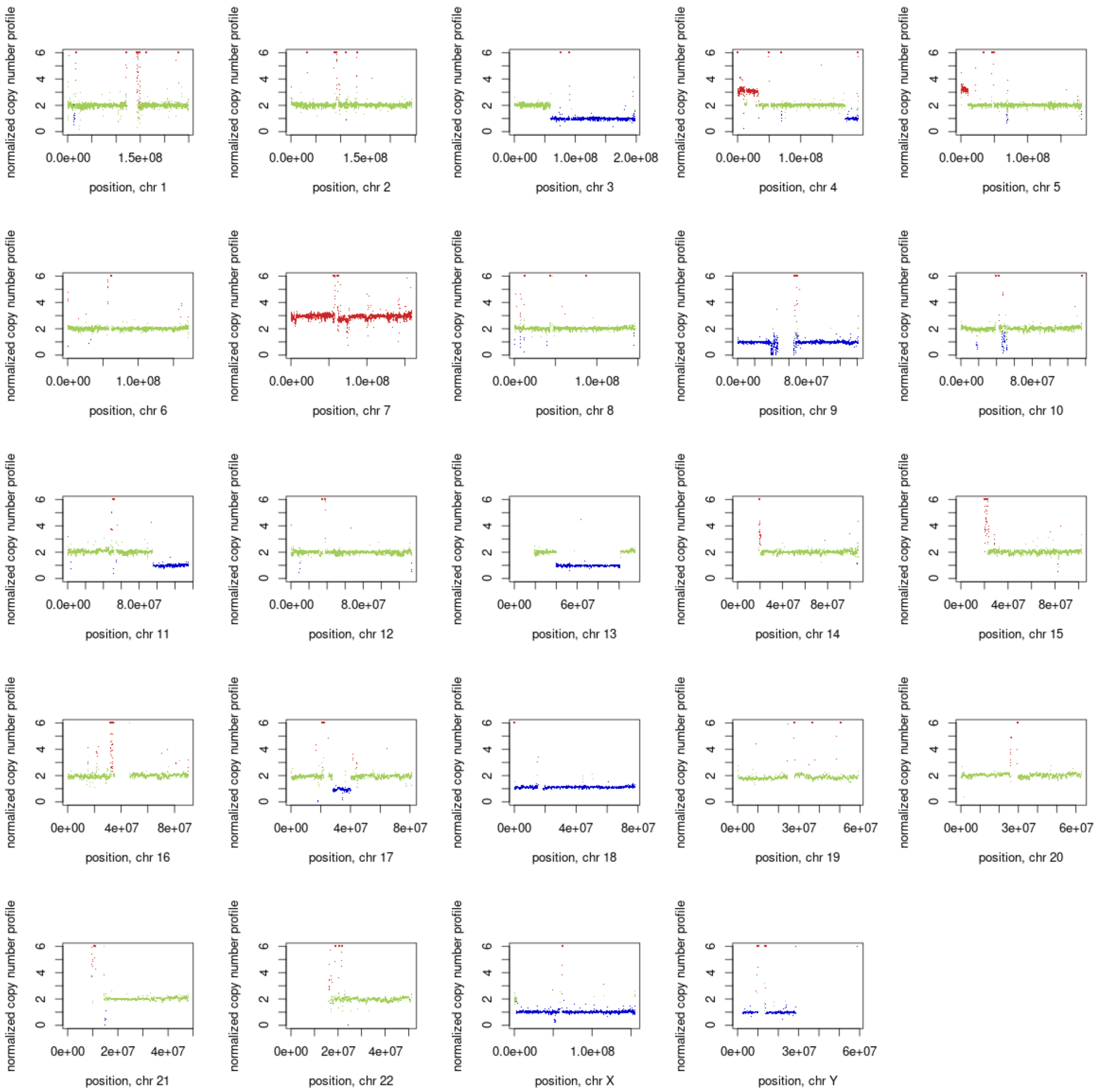

normalized copy number profile
position, chr 1
position, chr 2
position, chr 3
position, chr 4
position, chr 5
position, chr 6
position, chr 7
position, chr 8
position, chr 9
position, chr 10
position, chr 11
position, chr 12
position, chr 13
position, chr 14
position, chr 15
position, chr 16
position, chr 17
position, chr 18
position, chr 19
position, chr 20
position, chr 21
position, chr 22
position, chr X
position, chr Y


14M

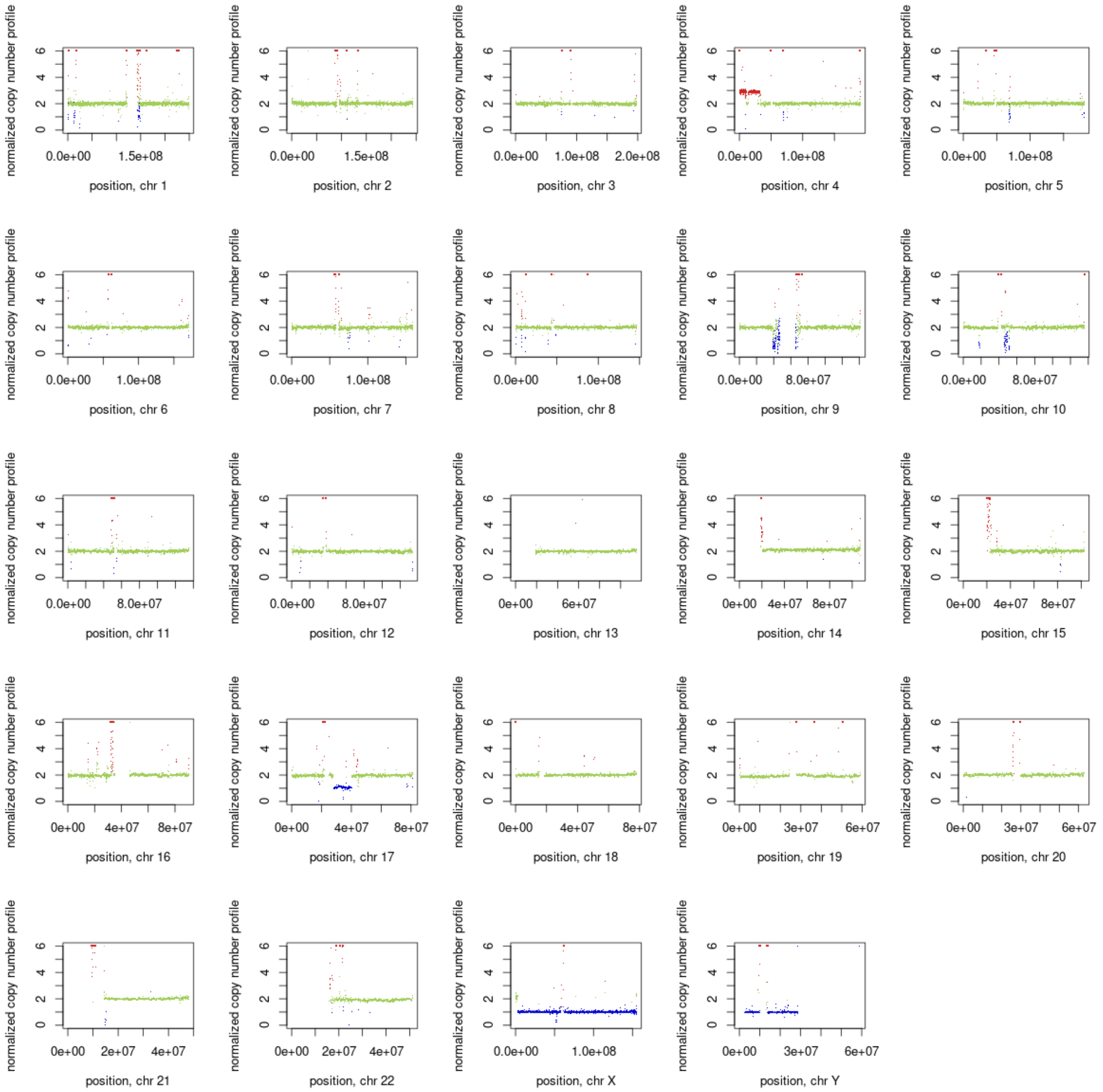
normalized copy number profile
0 2 4 6
0.0e+00 1.5e+08
position, chr 1
0.0e+00 1.5e+08
position, chr 2
0.0e+00 1.0e+08 2.0e+08
position, chr 3
0.0e+00 1.0e+08
position, chr 4
0.0e+00 1.0e+08
position, chr 5
0.0e+00 1.0e+08
position, chr 6
0.0e+00 1.0e+08
position, chr 7
0.0e+00 1.0e+08
position, chr 8
0.0e+00 8.0e+07
position, chr 9
0.0e+00 8.0e+07
position, chr 10
0.0e+00 8.0e+07
position, chr 11
0.0e+00 8.0e+07
position, chr 12
0e+00 6e+07
position, chr 13
0e+00 4e+07 8e+07
position, chr 14
0e+00 4e+07 8e+07
position, chr 15
0e+00 4e+07 8e+07
position, chr 16
0e+00 4e+07 8e+07
position, chr 17
0e+00 4e+07 8e+07
position, chr 18
0e+00 3e+07 6e+07
position, chr 19
0e+00 3e+07 6e+07
position, chr 20
0e+00 2e+07 4e+07
position, chr 21
0e+00 2e+07 4e+07
position, chr 22
0.0e+00 1.0e+08
position, chr X
0e+00 3e+07 6e+07
position, chr Y

14P

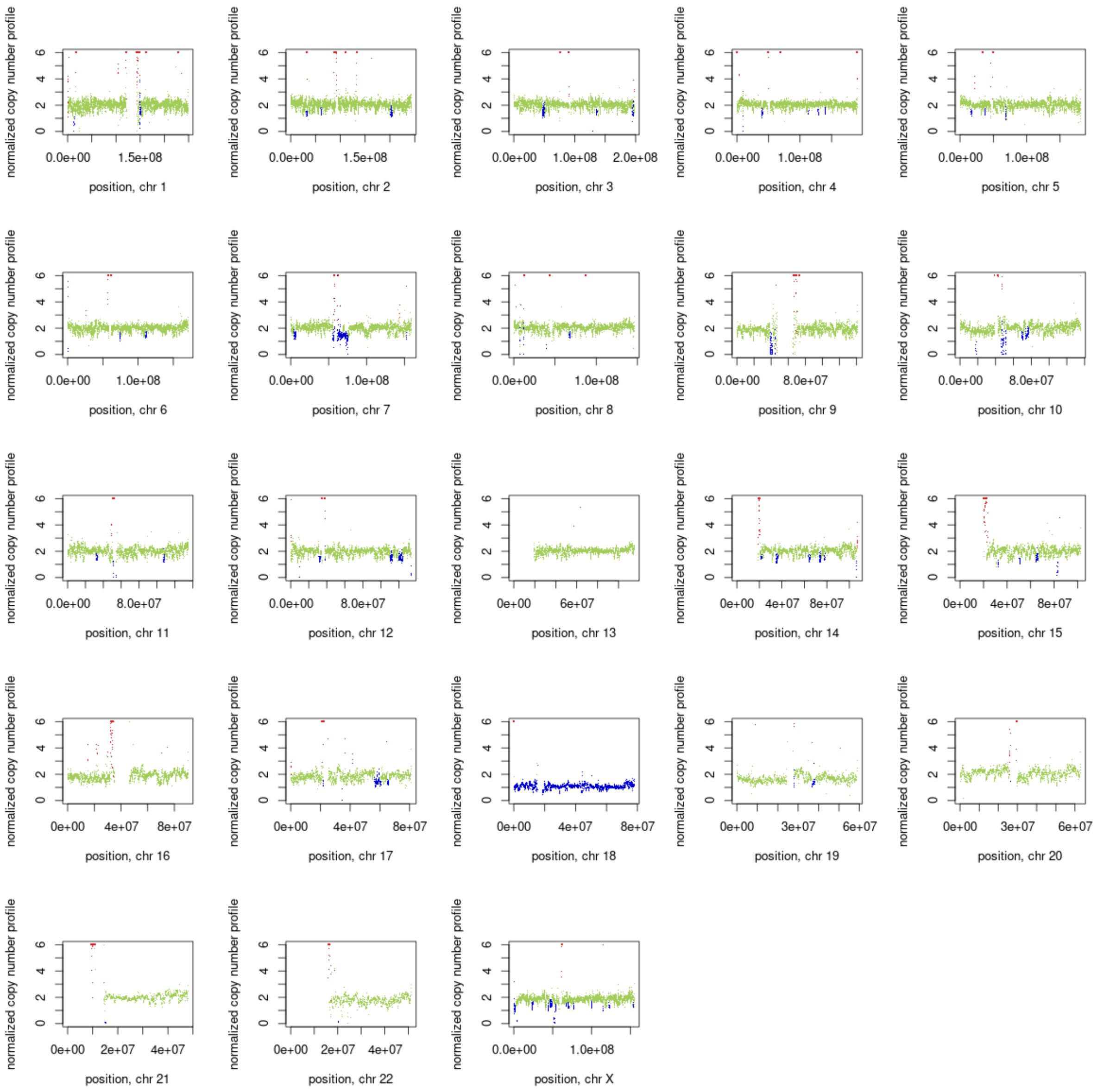
normalized copy number profile
0
2
4
6
0.0e+00
1.5e+08
position, chr 1
position, chr 2
1.0e+08
2.0e+08
position, chr 3
position, chr 4
position, chr 5
position, chr 6
position, chr 7
position, chr 8
8.0e+07
position, chr 9
position, chr 10
position, chr 11
position, chr 12
0e+00
6e+07
position, chr 13
4e+07
8e+07
position, chr 14
position, chr 15
position, chr 16
position, chr 17
position, chr 18
3e+07
6e+07
position, chr 19
position, chr 20
2e+07
position, chr 21
position, chr 22
position, chr X

3P

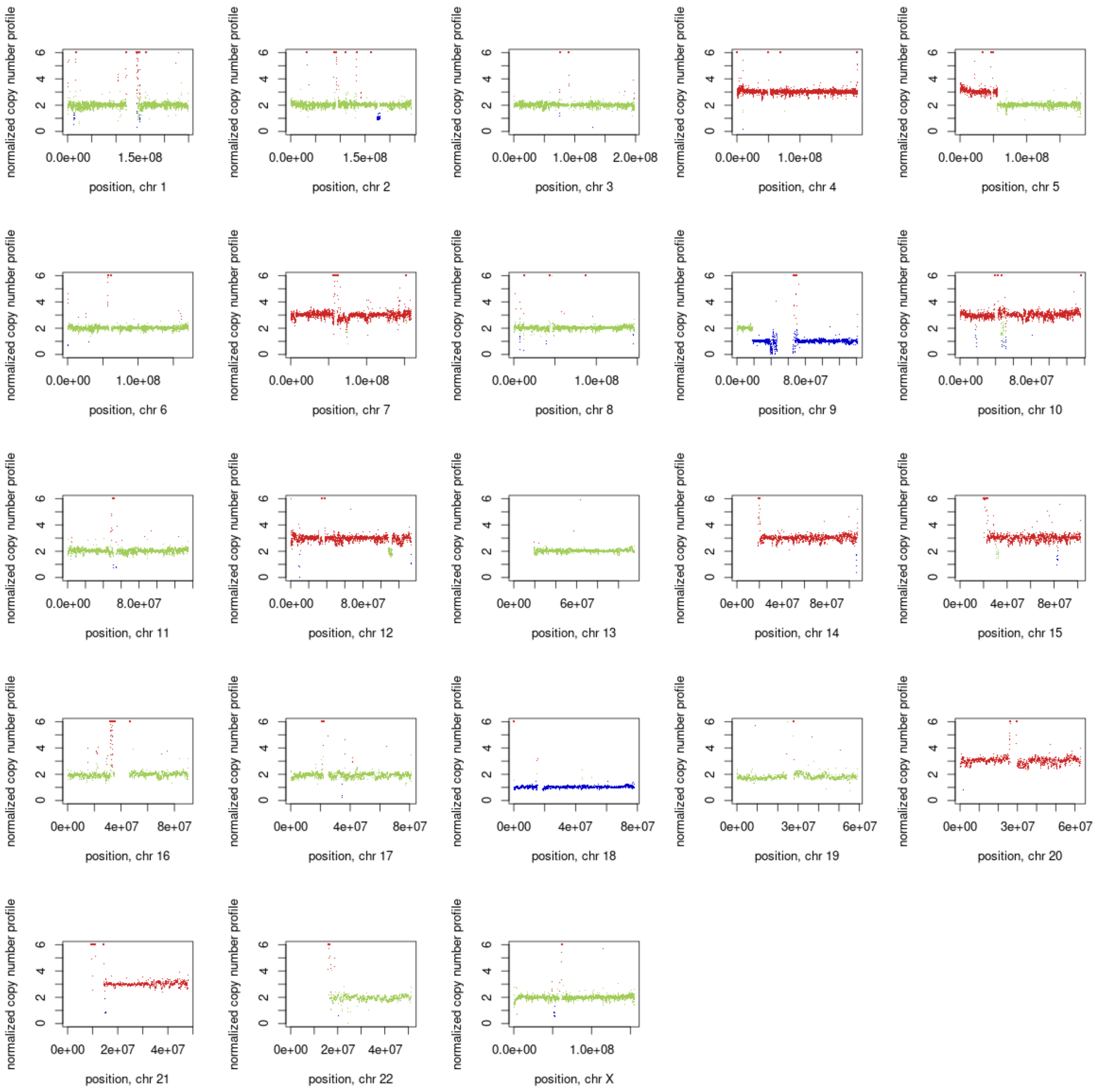
normalized copy number profile
0
2
4
6
0.0e+00
1.5e+08
position, chr 1
0.0e+00
1.5e+08
position, chr 2
0.0e+00
1.0e+08
2.0e+08
position, chr 3
0.0e+00
1.0e+08
position, chr 4
0.0e+00
1.0e+08
position, chr 5
0.0e+00
1.0e+08
position, chr 6
0.0e+00
1.0e+08
position, chr 7
0.0e+00
1.0e+08
position, chr 8
0.0e+00
8.0e+07
position, chr 9
0.0e+00
8.0e+07
position, chr 10
0.0e+00
8.0e+07
position, chr 11
0.0e+00
8.0e+07
position, chr 12
0e+00
6e+07
position, chr 13
0e+00
4e+07
8e+07
position, chr 14
0e+00
4e+07
8e+07
position, chr 15
0e+00
4e+07
8e+07
position, chr 16
0e+00
4e+07
8e+07
position, chr 17
0e+00
4e+07
8e+07
position, chr 18
0e+00
3e+07
6e+07
position, chr 19
0e+00
3e+07
6e+07
position, chr 20
0e+00
2e+07
4e+07
position, chr 21
0e+00
2e+07
4e+07
position, chr 22
0.0e+00
1.0e+08
position, chr X

3M

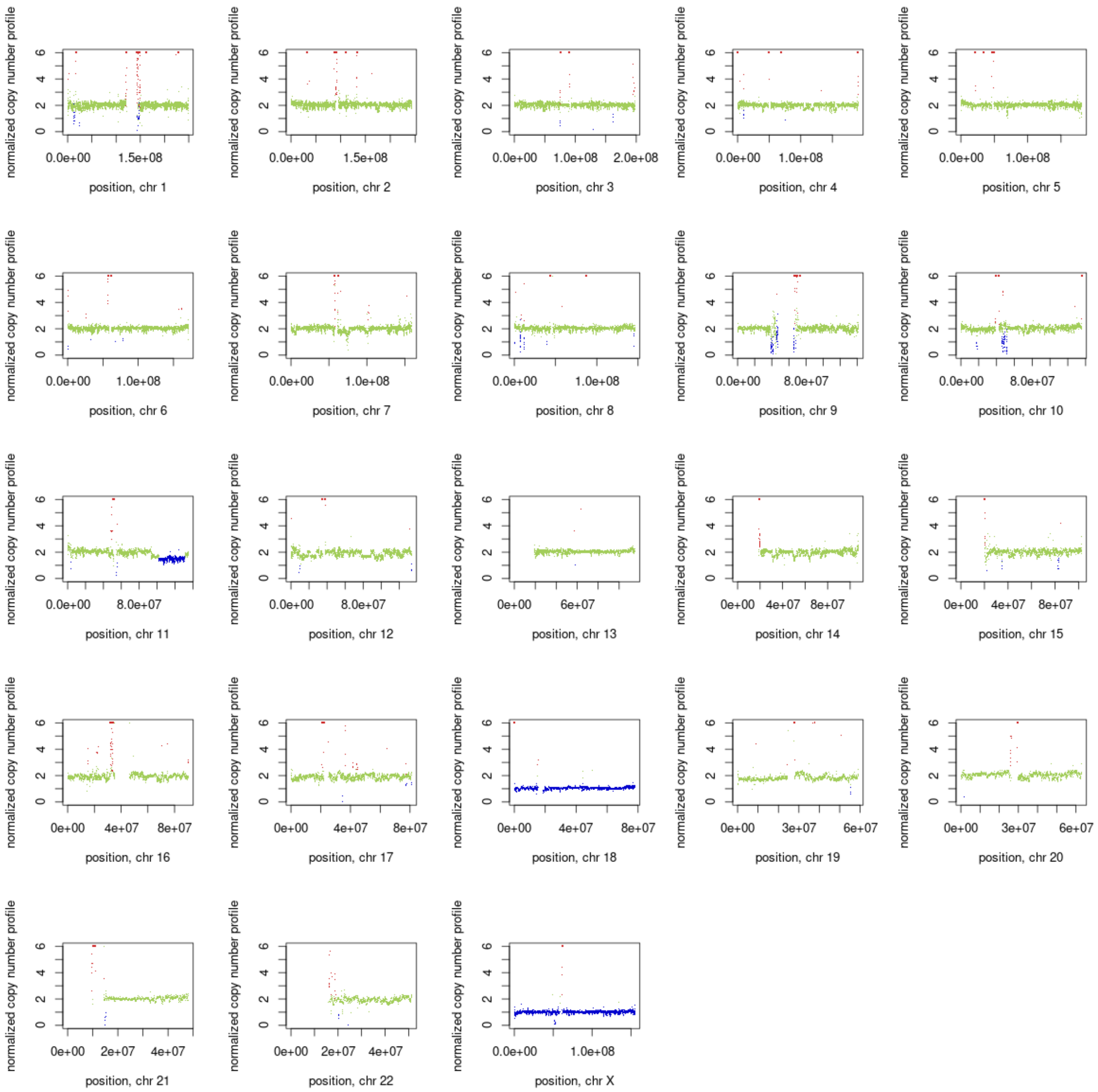
normalized copy number profile
0 2 4 6
0.0e+00 1.5e+08
position, chr 1
0.0e+00 1.5e+08
position, chr 2
0.0e+00 1.0e+08 2.0e+08
position, chr 3
0.0e+00 1.0e+08
position, chr 4
0.0e+00 1.0e+08
position, chr 5
0.0e+00 1.0e+08
position, chr 6
0.0e+00 1.0e+08
position, chr 7
0.0e+00 1.0e+08
position, chr 8
0.0e+00 8.0e+07
position, chr 9
0.0e+00 8.0e+07
position, chr 10
0.0e+00 8.0e+07
position, chr 11
0.0e+00 8.0e+07
position, chr 12
0e+00 6e+07
position, chr 13
0e+00 4e+07 8e+07
position, chr 14
0e+00 4e+07 8e+07
position, chr 15
0e+00 4e+07 8e+07
position, chr 16
0e+00 4e+07 8e+07
position, chr 17
0e+00 4e+07 8e+07
position, chr 18
0e+00 3e+07 6e+07
position, chr 19
0e+00 3e+07 6e+07
position, chr 20
0e+00 2e+07 4e+07
position, chr 21
0e+00 2e+07 4e+07
position, chr 22
0.0e+00 1.0e+08
position, chr X

26P

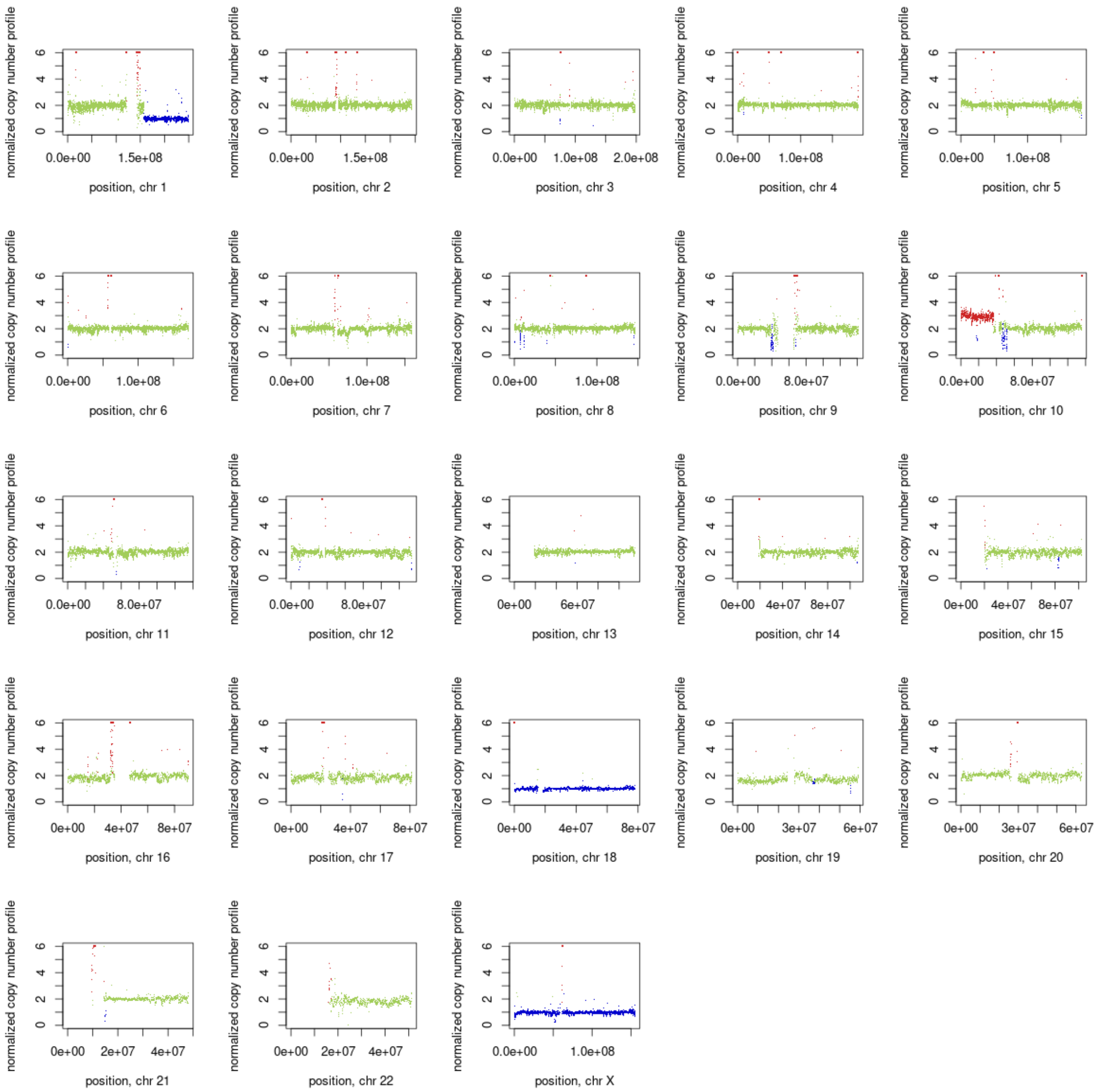
normalized copy number profile
position, chr 1
position, chr 2
position, chr 3
position, chr 4
position, chr 5
position, chr 6
position, chr 7
position, chr 8
position, chr 9
position, chr 10
position, chr 11
position, chr 12
position, chr 13
position, chr 14
position, chr 15
position, chr 16
position, chr 17
position, chr 18
position, chr 19
position, chr 20
position, chr 21
position, chr 22
position, chr X

26M

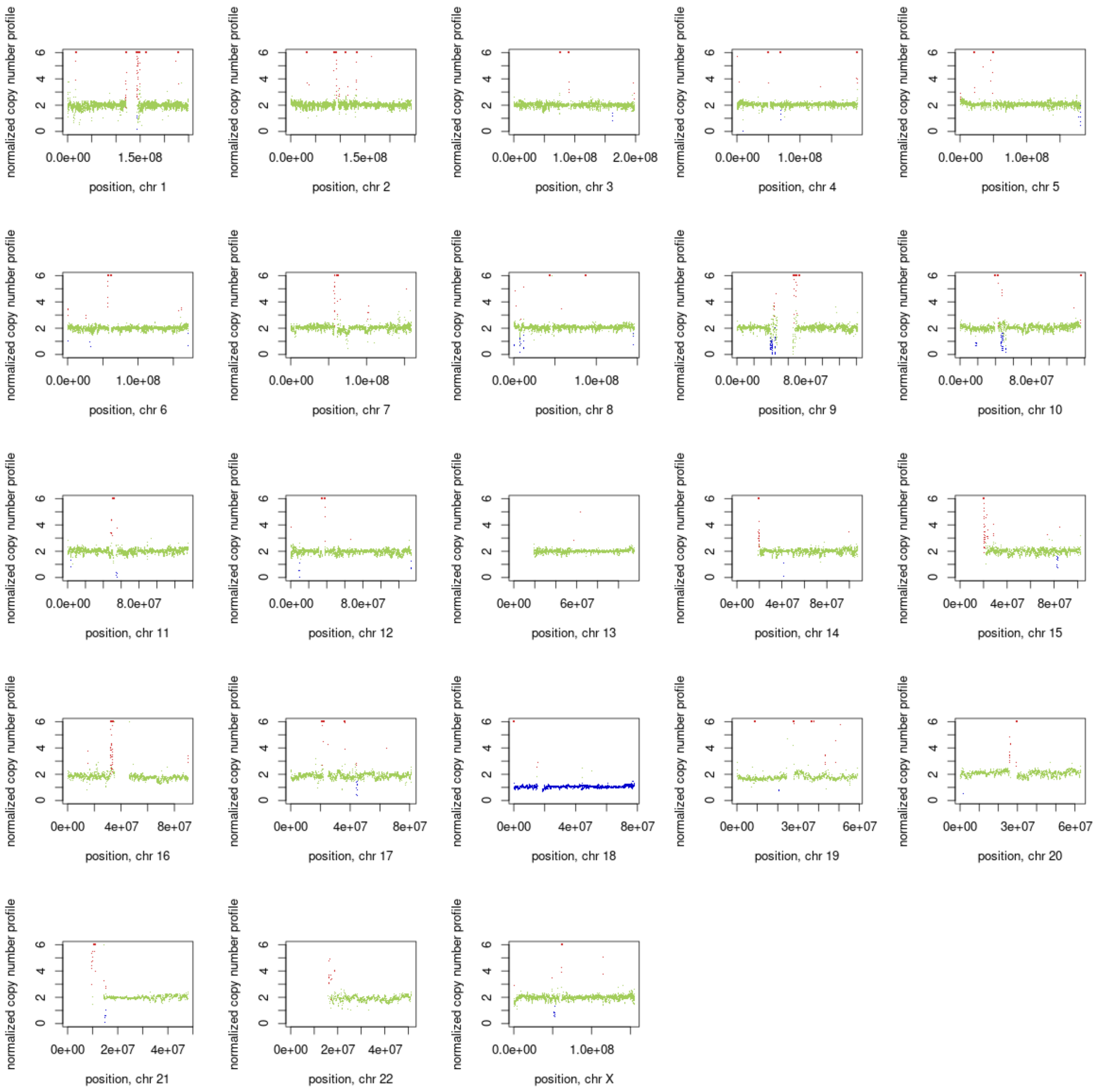
normalized copy number profile
0
2
4
6
0.0e+00 1.5e+08
position, chr 1
0.0e+00 1.5e+08
position, chr 2
0.0e+00 1.0e+08 2.0e+08
position, chr 3
0.0e+00 1.0e+08
position, chr 4
0.0e+00 1.0e+08
position, chr 5
0.0e+00 1.0e+08
position, chr 6
0.0e+00 1.0e+08
position, chr 7
0.0e+00 1.0e+08
position, chr 8
0.0e+00 8.0e+07
position, chr 9
0.0e+00 8.0e+07
position, chr 10
0.0e+00 8.0e+07
position, chr 11
0.0e+00 8.0e+07
position, chr 12
0e+00 6e+07
position, chr 13
0e+00 4e+07 8e+07
position, chr 14
0e+00 4e+07 8e+07
position, chr 15
0e+00 4e+07 8e+07
position, chr 16
0e+00 4e+07 8e+07
position, chr 17
0e+00 4e+07 8e+07
position, chr 18
0e+00 3e+07 6e+07
position, chr 19
0e+00 3e+07 6e+07
position, chr 20
0e+00 2e+07 4e+07
position, chr 21
0e+00 2e+07 4e+07
position, chr 22
0.0e+00 1.0e+08
position, chr X

10P

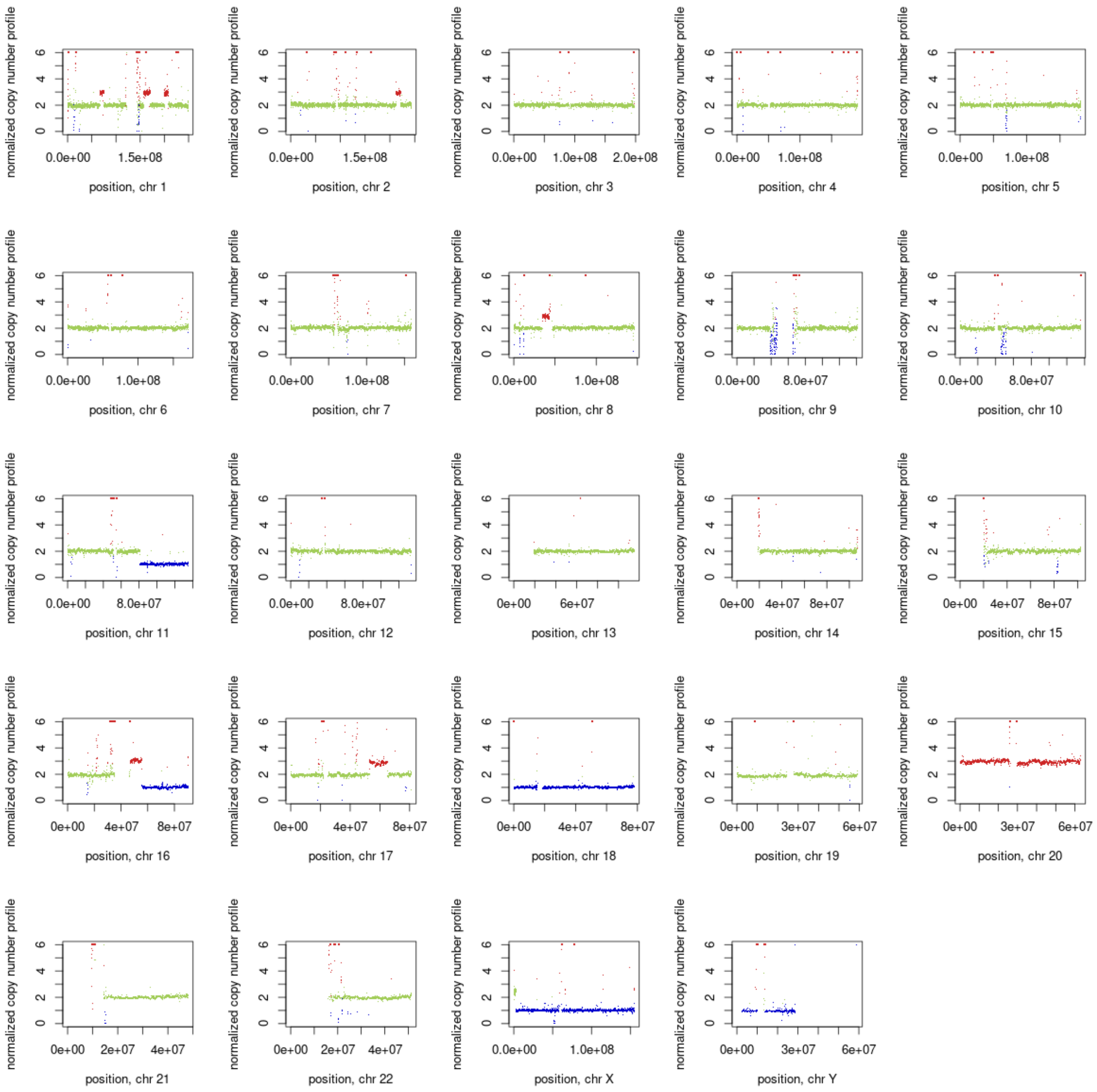
normalized copy number profile
position, chr 1
position, chr 2
position, chr 3
position, chr 4
position, chr 5
position, chr 6
position, chr 7
position, chr 8
position, chr 9
position, chr 10
position, chr 11
position, chr 12
position, chr 13
position, chr 14
position, chr 15
position, chr 16
position, chr 17
position, chr 18
position, chr 19
position, chr 20
position, chr 21
position, chr 22
position, chr X
position, chr Y

5P

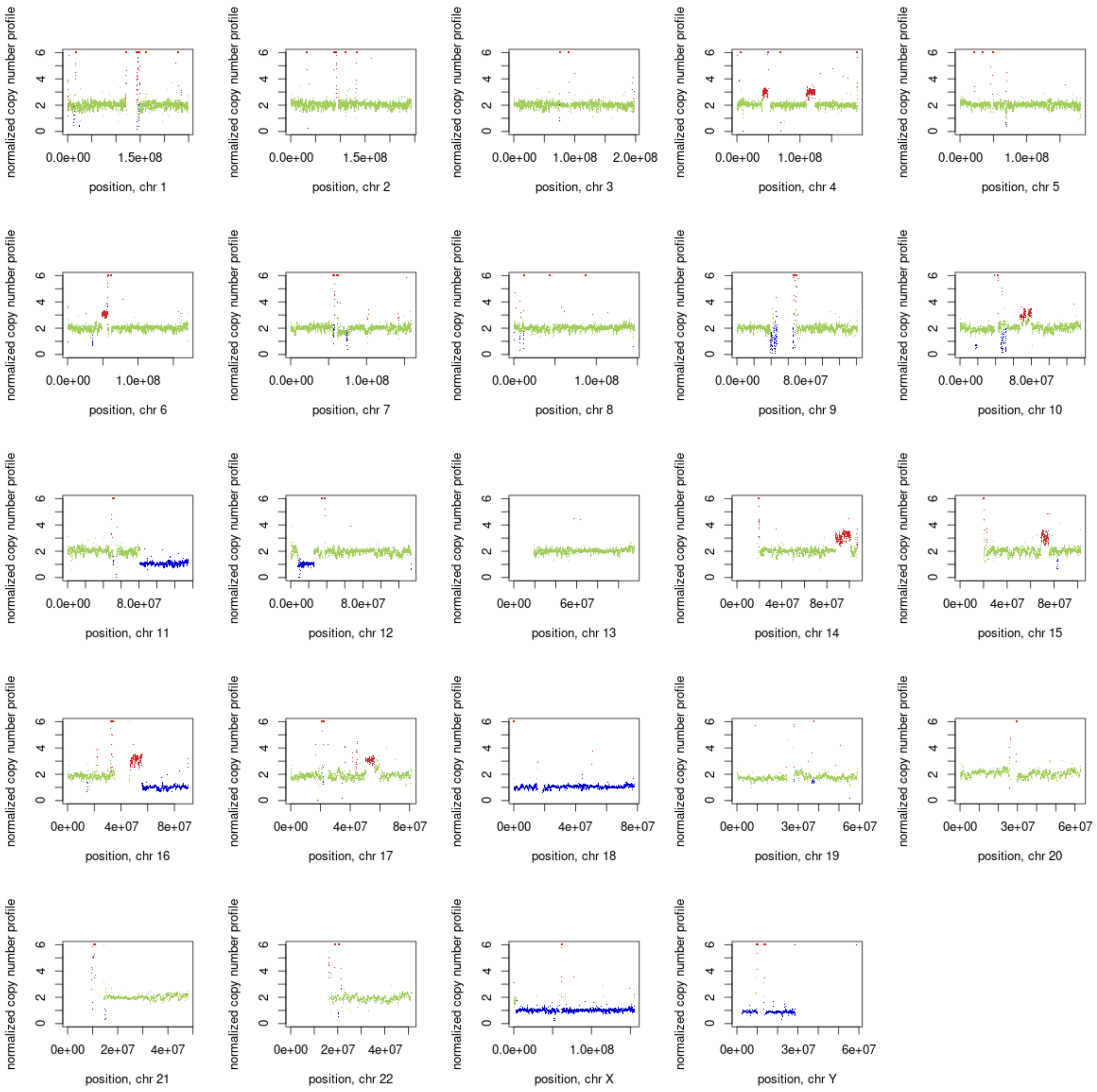
normalized copy number profile
position, chr 1
position, chr 2
position, chr 3
position, chr 4
position, chr 5
position, chr 6
position, chr 7
position, chr 8
position, chr 9
position, chr 10
position, chr 11
position, chr 12
position, chr 13
position, chr 14
position, chr 15
position, chr 16
position, chr 17
position, chr 18
position, chr 19
position, chr 20
position, chr 21
position, chr 22
position, chr X
position, chr Y

5M

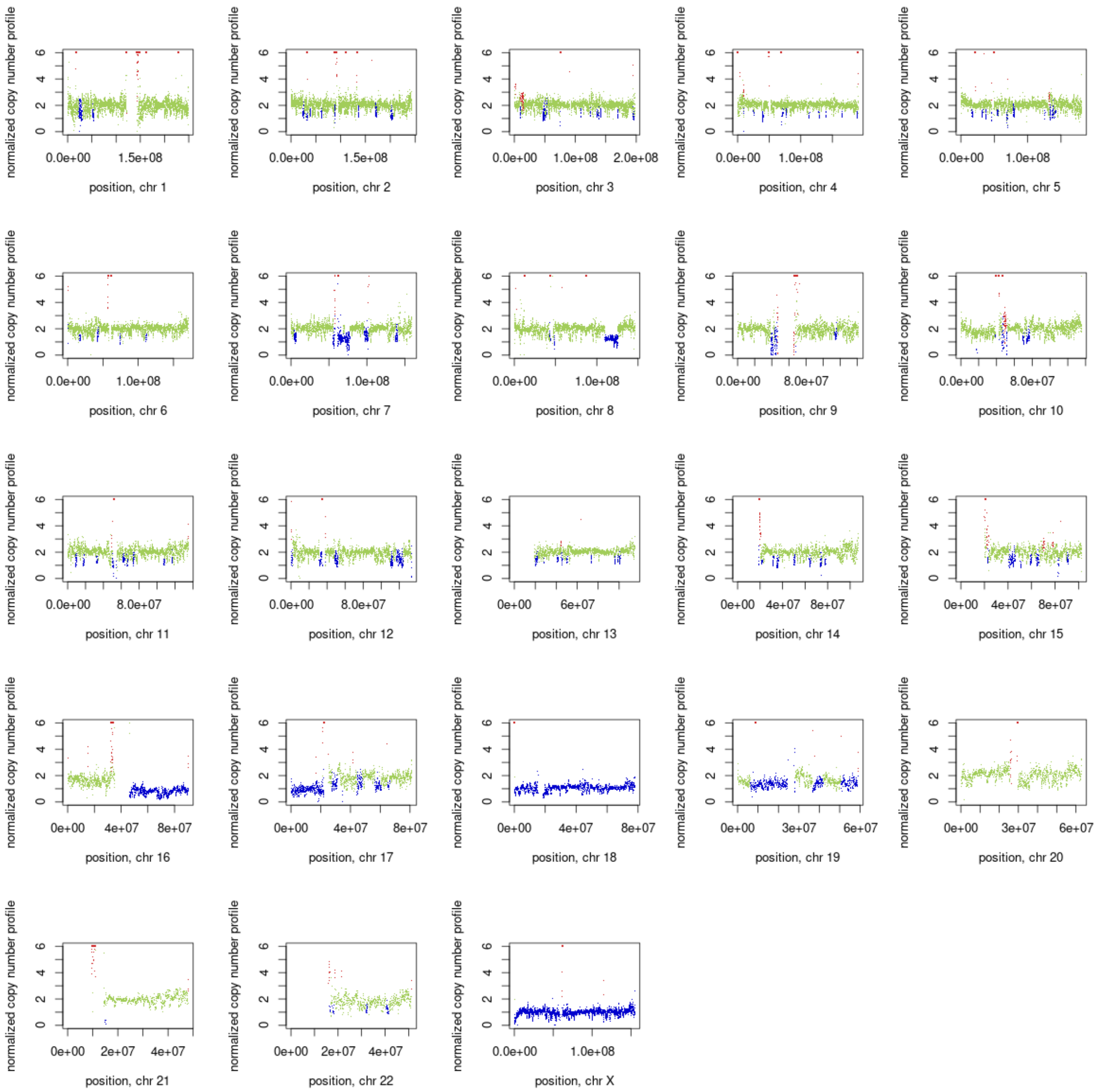
normalized copy number profile
0
2
4
6
0.0e+00
1.5e+08
position, chr 1
0.0e+00
1.5e+08
position, chr 2
0.0e+00
1.0e+08
2.0e+08
position, chr 3
0.0e+00
1.0e+08
position, chr 4
0.0e+00
1.0e+08
position, chr 5
0.0e+00
1.0e+08
position, chr 6
0.0e+00
1.0e+08
position, chr 7
0.0e+00
1.0e+08
position, chr 8
0.0e+00
8.0e+07
position, chr 9
0.0e+00
8.0e+07
position, chr 10
0.0e+00
8.0e+07
position, chr 11
0.0e+00
8.0e+07
position, chr 12
0e+00
6e+07
position, chr 13
0e+00
4e+07
8e+07
position, chr 14
0e+00
4e+07
8e+07
position, chr 15
0e+00
4e+07
8e+07
position, chr 16
0e+00
4e+07
8e+07
position, chr 17
0e+00
4e+07
8e+07
position, chr 18
0e+00
3e+07
6e+07
position, chr 19
0e+00
3e+07
6e+07
position, chr 20
0e+00
2e+07
4e+07
position, chr 21
0e+00
2e+07
4e+07
position, chr 22
0.0e+00
1.0e+08
position, chr X

12P

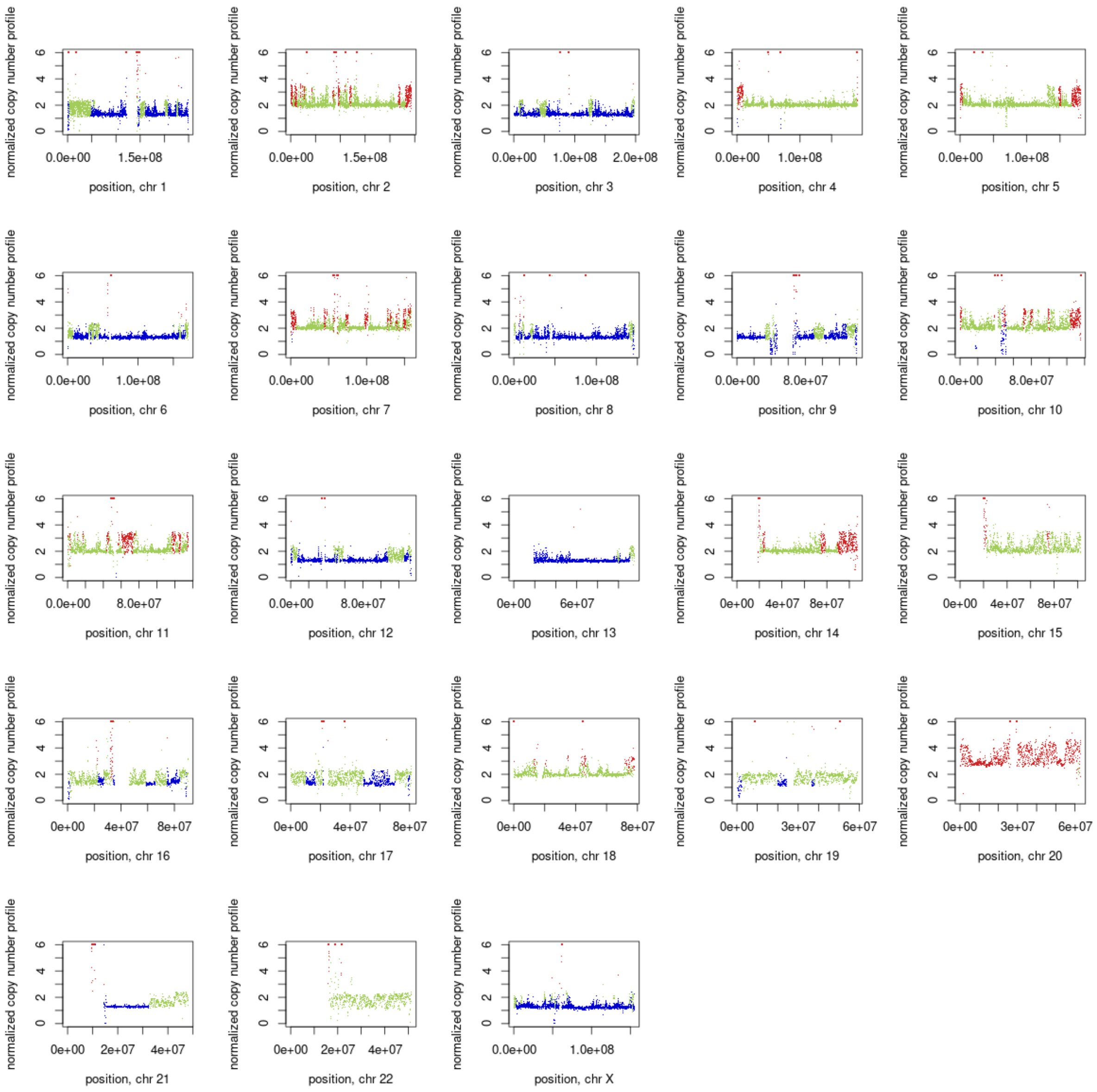
normalized copy number profile
0 2 4 6
0.0e+00 1.5e+08
position, chr 1
0.0e+00 1.5e+08
position, chr 2
0.0e+00 1.0e+08 2.0e+08
position, chr 3
0.0e+00 1.0e+08
position, chr 4
0.0e+00 1.0e+08
position, chr 5
0.0e+00 1.0e+08
position, chr 6
0.0e+00 1.0e+08
position, chr 7
0.0e+00 1.0e+08
position, chr 8
0.0e+00 8.0e+07
position, chr 9
0.0e+00 8.0e+07
position, chr 10
0.0e+00 8.0e+07
position, chr 11
0.0e+00 8.0e+07
position, chr 12
0e+00 6e+07
position, chr 13
0e+00 4e+07 8e+07
position, chr 14
0e+00 4e+07 8e+07
position, chr 15
0e+00 4e+07 8e+07
position, chr 16
0e+00 4e+07 8e+07
position, chr 17
0e+00 4e+07 8e+07
position, chr 18
0e+00 3e+07 6e+07
position, chr 19
0e+00 3e+07 6e+07
position, chr 20
0e+00 2e+07 4e+07
position, chr 21
0e+00 2e+07 4e+07
position, chr 22
0.0e+00 1.0e+08
position, chr X

23M

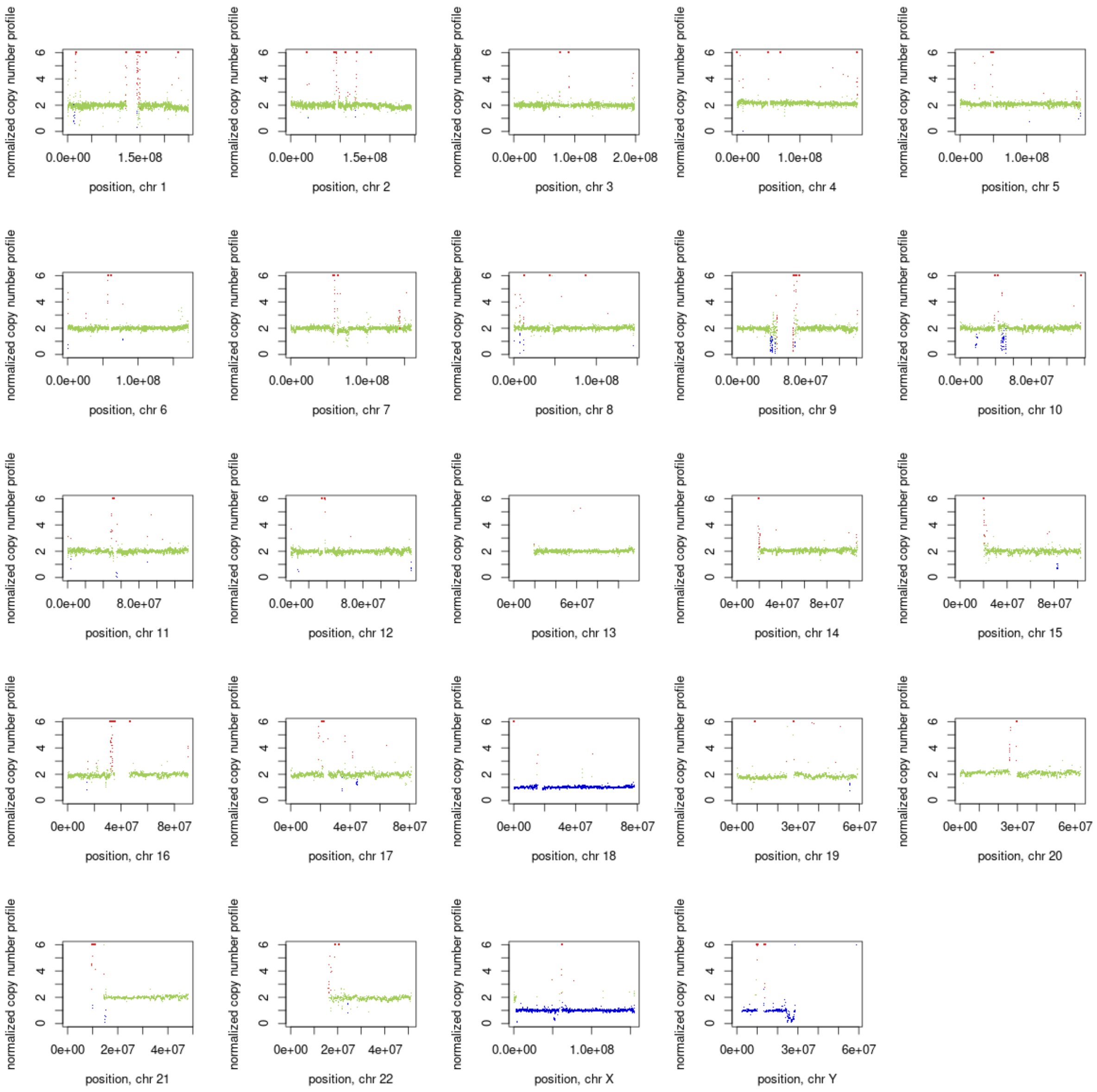
normalized copy number profile
0
2
4
6
position, chr 1
position, chr 2
position, chr 3
position, chr 4
position, chr 5
position, chr 6
position, chr 7
position, chr 8
position, chr 9
position, chr 10
position, chr 11
position, chr 12
position, chr 13
position, chr 14
position, chr 15
position, chr 16
position, chr 17
position, chr 18
position, chr 19
position, chr 20
position, chr 21
position, chr 22
position, chr X
position, chr Y
0.0e+00
1.5e+08
1.0e+08
2.0e+08
8.0e+07
0e+00
6e+07
4e+07
8e+07
3e+07
2e+07

16P

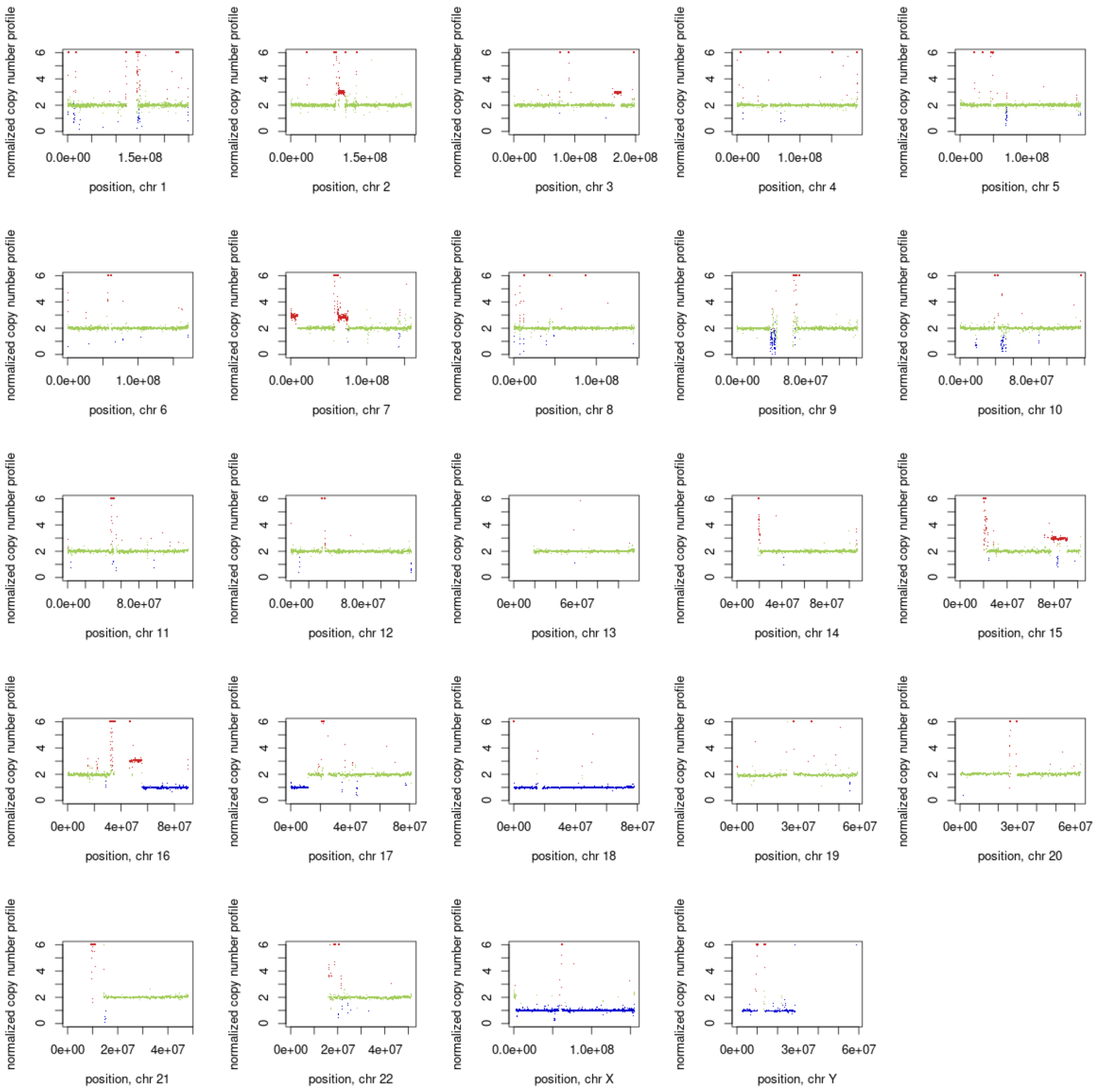
normalized copy number profile
0
2
4
6
0.0e+00
1.5e+08
position, chr 1
0.0e+00
1.5e+08
position, chr 2
0.0e+00
1.0e+08
2.0e+08
position, chr 3
0.0e+00
1.0e+08
position, chr 4
0.0e+00
1.0e+08
position, chr 5
0.0e+00
1.0e+08
position, chr 6
0.0e+00
1.0e+08
position, chr 7
0.0e+00
1.0e+08
position, chr 8
0.0e+00
8.0e+07
position, chr 9
0.0e+00
8.0e+07
position, chr 10
0.0e+00
8.0e+07
position, chr 11
0.0e+00
8.0e+07
position, chr 12
0e+00
6e+07
position, chr 13
0e+00
4e+07
8e+07
position, chr 14
0e+00
4e+07
8e+07
position, chr 15
0e+00
4e+07
8e+07
position, chr 16
0e+00
4e+07
8e+07
position, chr 17
0e+00
4e+07
8e+07
position, chr 18
0e+00
3e+07
6e+07
position, chr 19
0e+00
3e+07
6e+07
position, chr 20
0e+00
2e+07
4e+07
position, chr 21
0e+00
2e+07
4e+07
position, chr 22
0.0e+00
1.0e+08
position, chr X
0e+00
3e+07
6e+07
position, chr Y

7P

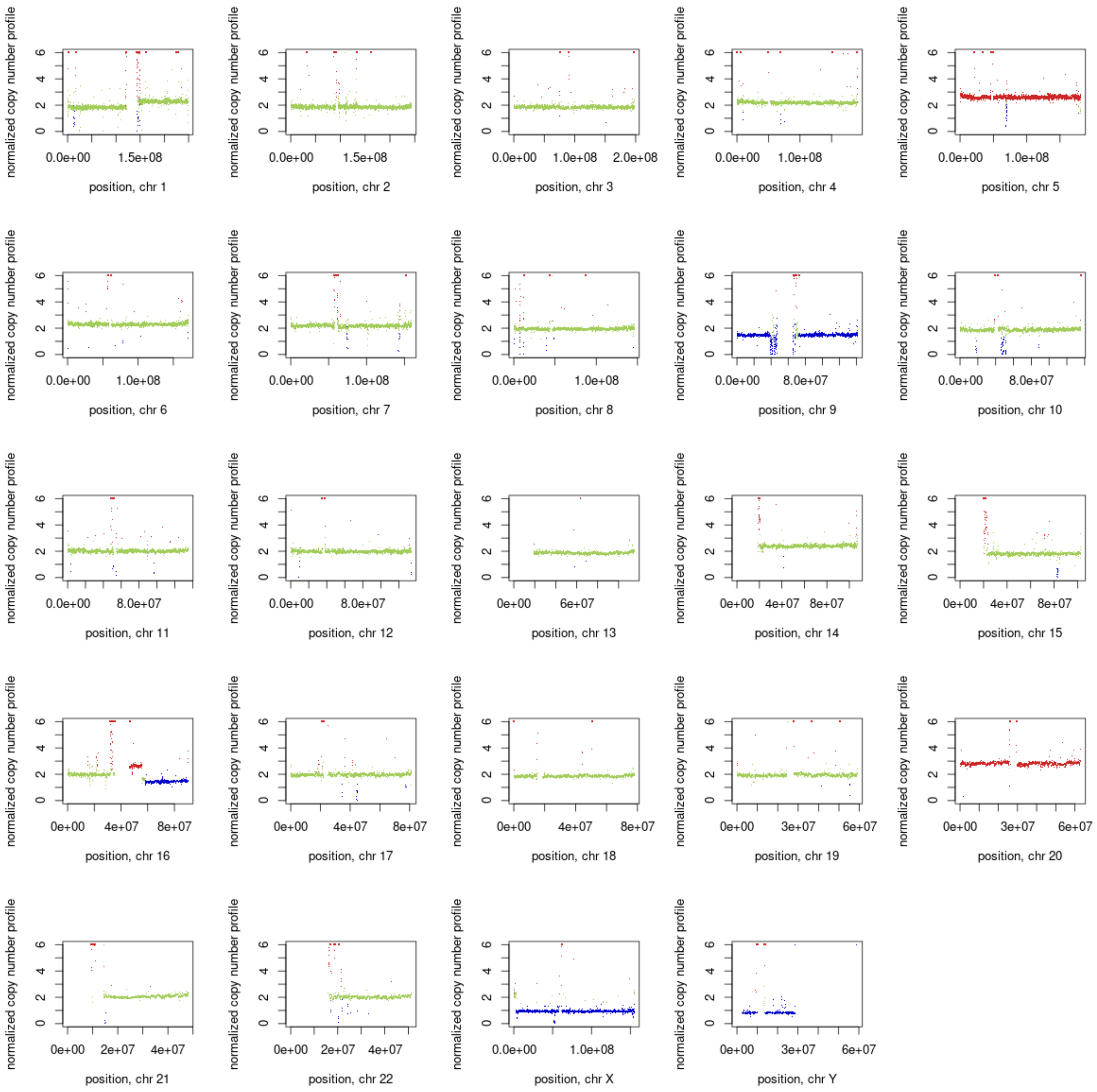
normalized copy number profile
0
2
4
6
0.0e+00 1.5e+08
position, chr 1
0.0e+00 1.5e+08
position, chr 2
0.0e+00 1.0e+08 2.0e+08
position, chr 3
0.0e+00 1.0e+08
position, chr 4
0.0e+00 1.0e+08
position, chr 5
0.0e+00 1.0e+08
position, chr 6
0.0e+00 1.0e+08
position, chr 7
0.0e+00 1.0e+08
position, chr 8
0.0e+00 8.0e+07
position, chr 9
0.0e+00 8.0e+07
position, chr 10
0.0e+00 8.0e+07
position, chr 11
0.0e+00 8.0e+07
position, chr 12
0e+00 6e+07
position, chr 13
0e+00 4e+07 8e+07
position, chr 14
0e+00 4e+07 8e+07
position, chr 15
0e+00 4e+07 8e+07
position, chr 16
0e+00 4e+07 8e+07
position, chr 17
0e+00 4e+07 8e+07
position, chr 18
0e+00 3e+07 6e+07
position, chr 19
0e+00 3e+07 6e+07
position, chr 20
0e+00 2e+07 4e+07
position, chr 21
0e+00 2e+07 4e+07
position, chr 22
0.0e+00 1.0e+08
position, chr X
0e+00 3e+07 6e+07
position, chr Y

7M

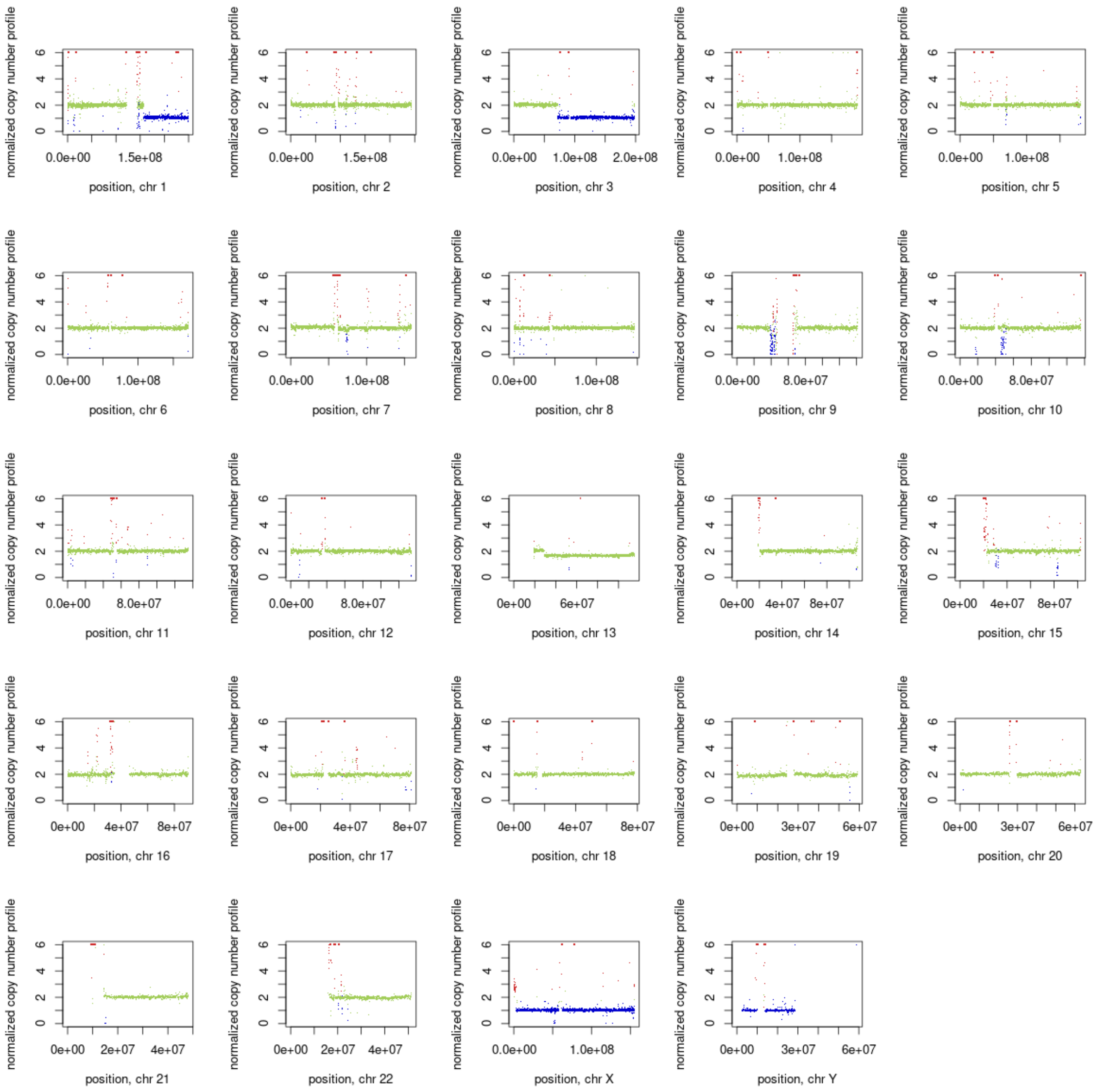
normalized copy number profile
position, chr 1
position, chr 2
position, chr 3
position, chr 4
position, chr 5
position, chr 6
position, chr 7
position, chr 8
position, chr 9
position, chr 10
position, chr 11
position, chr 12
position, chr 13
position, chr 14
position, chr 15
position, chr 16
position, chr 17
position, chr 18
position, chr 19
position, chr 20
position, chr 21
position, chr 22
position, chr X
position, chr Y

24P

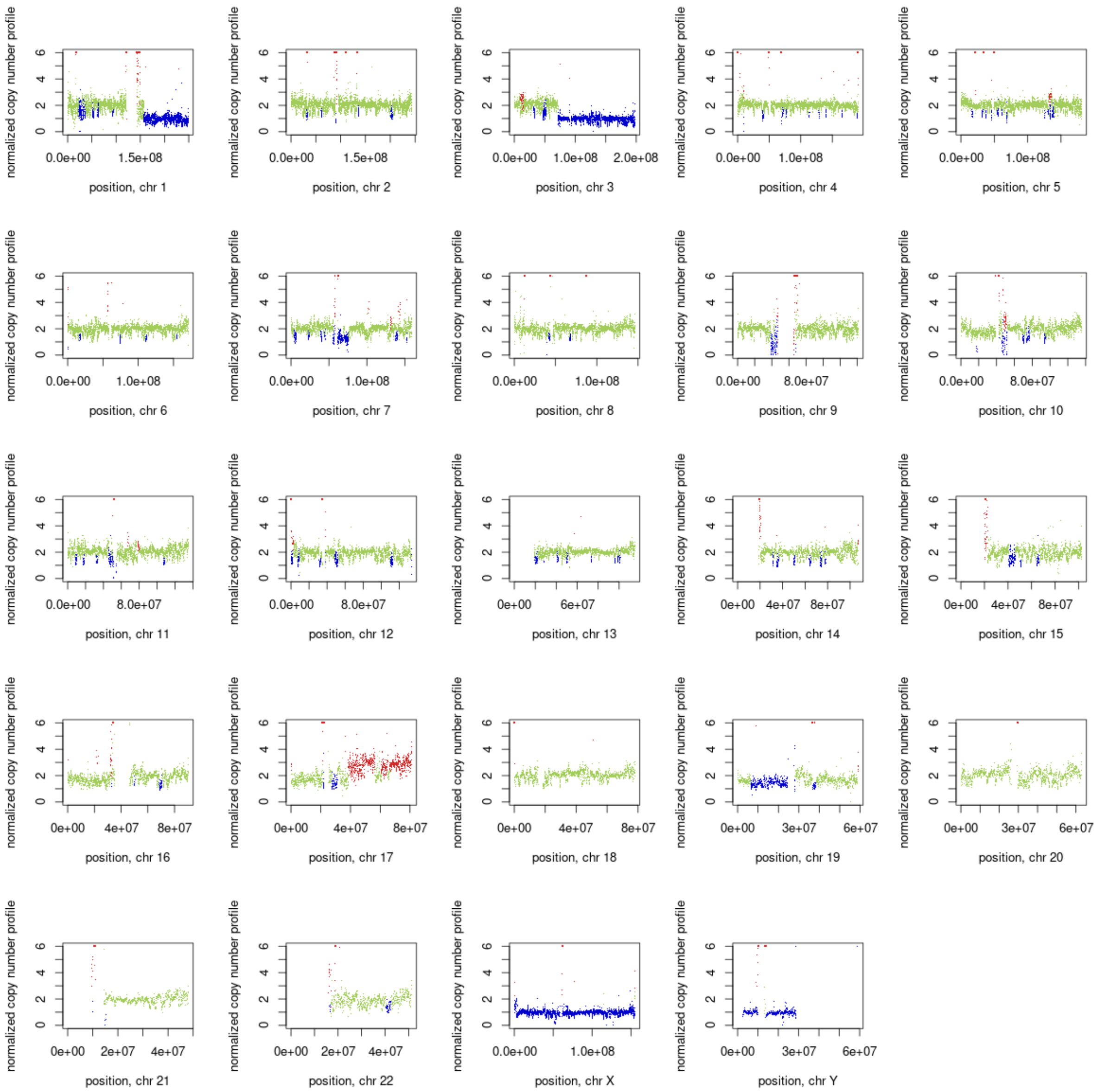
normalized copy number profile
position, chr 1
position, chr 2
position, chr 3
position, chr 4
position, chr 5
position, chr 6
position, chr 7
position, chr 8
position, chr 9
position, chr 10
position, chr 11
position, chr 12
position, chr 13
position, chr 14
position, chr 15
position, chr 16
position, chr 17
position, chr 18
position, chr 19
position, chr 20
position, chr 21
position, chr 22
position, chr X
position, chr Y

24M

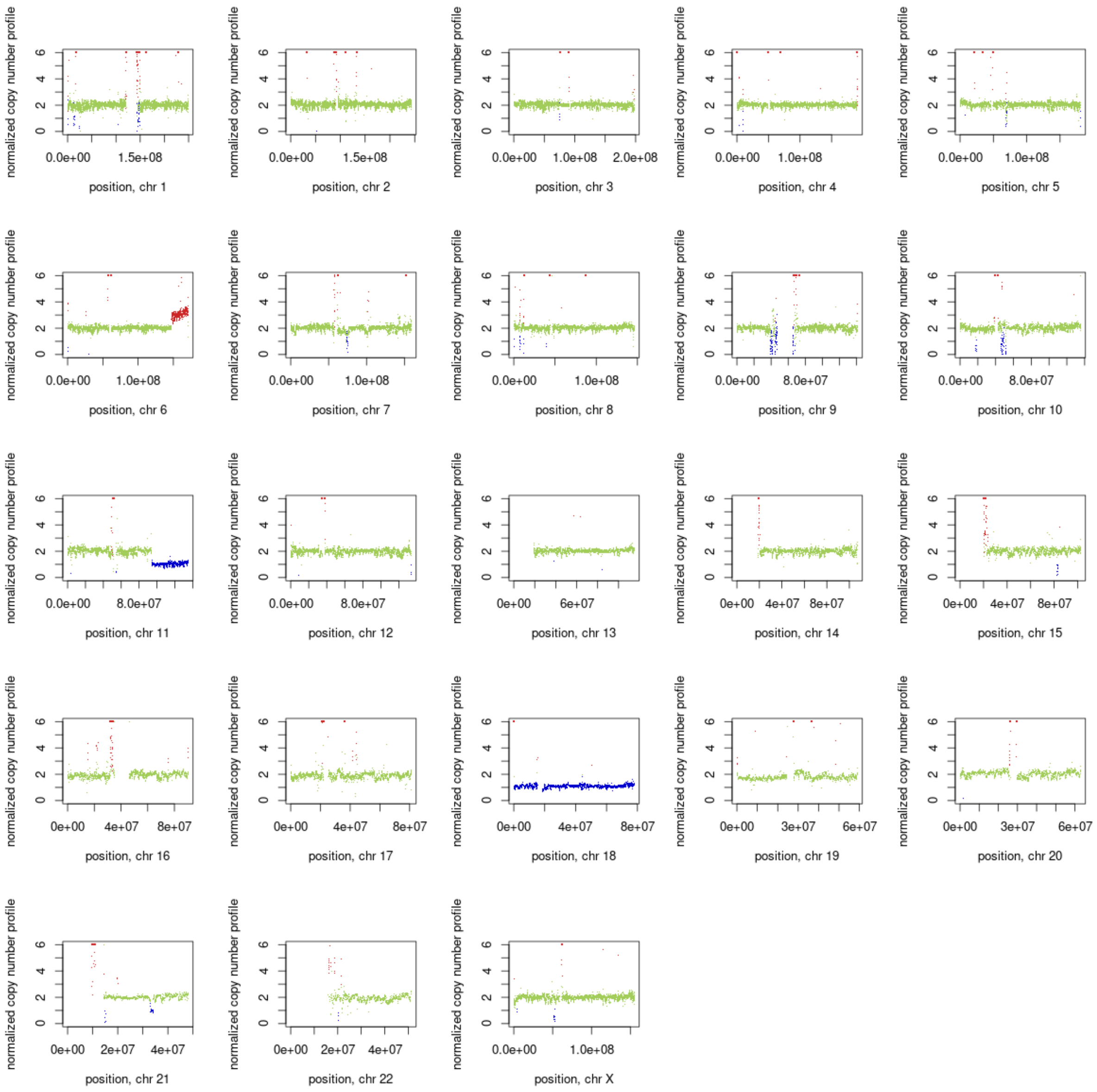
normalized copy number profile
0
2
4
6
0.0e+00
1.5e+08
position, chr 1
0.0e+00
1.5e+08
position, chr 2
0.0e+00
1.0e+08
2.0e+08
position, chr 3
0.0e+00
1.0e+08
position, chr 4
0.0e+00
1.0e+08
position, chr 5
0.0e+00
1.0e+08
position, chr 6
0.0e+00
1.0e+08
position, chr 7
0.0e+00
1.0e+08
position, chr 8
0.0e+00
8.0e+07
position, chr 9
0.0e+00
8.0e+07
position, chr 10
0.0e+00
8.0e+07
position, chr 11
0.0e+00
8.0e+07
position, chr 12
0e+00
6e+07
position, chr 13
0e+00
4e+07
8e+07
position, chr 14
0e+00
4e+07
8e+07
position, chr 15
0e+00
4e+07
8e+07
position, chr 16
0e+00
4e+07
8e+07
position, chr 17
0e+00
4e+07
8e+07
position, chr 18
0e+00
3e+07
6e+07
position, chr 19
0e+00
3e+07
6e+07
position, chr 20
0e+00
2e+07
4e+07
position, chr 21
0e+00
2e+07
4e+07
position, chr 22
0.0e+00
1.0e+08
position, chr X

21P

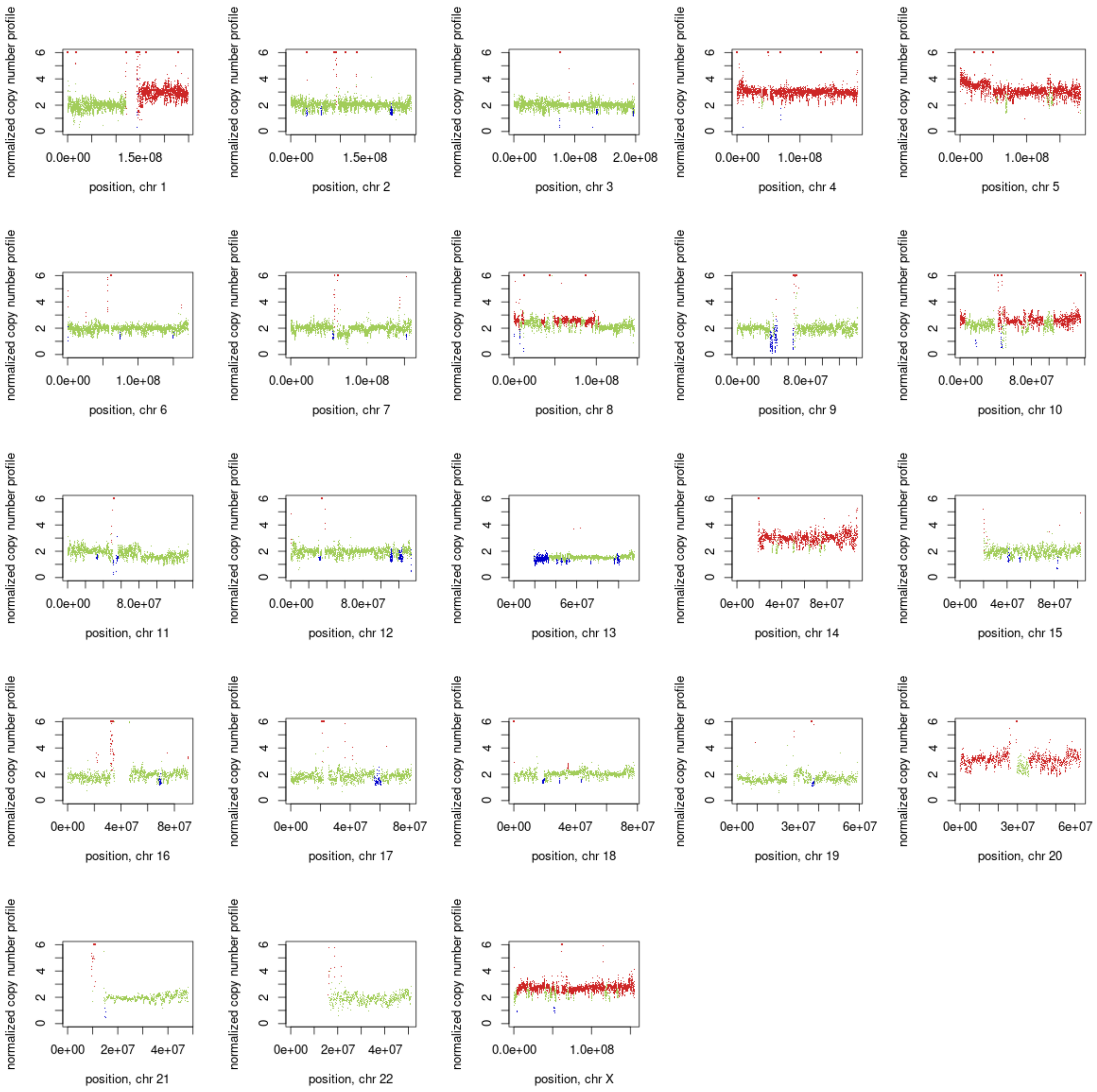
normalized copy number profile
0
2
4
6
0.0e+00
1.5e+08
position, chr 1
0.0e+00
1.5e+08
position, chr 2
0.0e+00
1.0e+08
2.0e+08
position, chr 3
0.0e+00
1.0e+08
position, chr 4
0.0e+00
1.0e+08
position, chr 5
0.0e+00
1.0e+08
position, chr 6
0.0e+00
1.0e+08
position, chr 7
0.0e+00
1.0e+08
position, chr 8
0.0e+00
8.0e+07
position, chr 9
0.0e+00
8.0e+07
position, chr 10
0.0e+00
8.0e+07
position, chr 11
0.0e+00
8.0e+07
position, chr 12
0e+00
6e+07
position, chr 13
0e+00
4e+07
8e+07
position, chr 14
0e+00
4e+07
8e+07
position, chr 15
0e+00
4e+07
8e+07
position, chr 16
0e+00
4e+07
8e+07
position, chr 17
0e+00
4e+07
8e+07
position, chr 18
0e+00
3e+07
6e+07
position, chr 19
0e+00
3e+07
6e+07
position, chr 20
0e+00
2e+07
4e+07
position, chr 21
0e+00
2e+07
4e+07
position, chr 22
0.0e+00
1.0e+08
position, chr X

15M

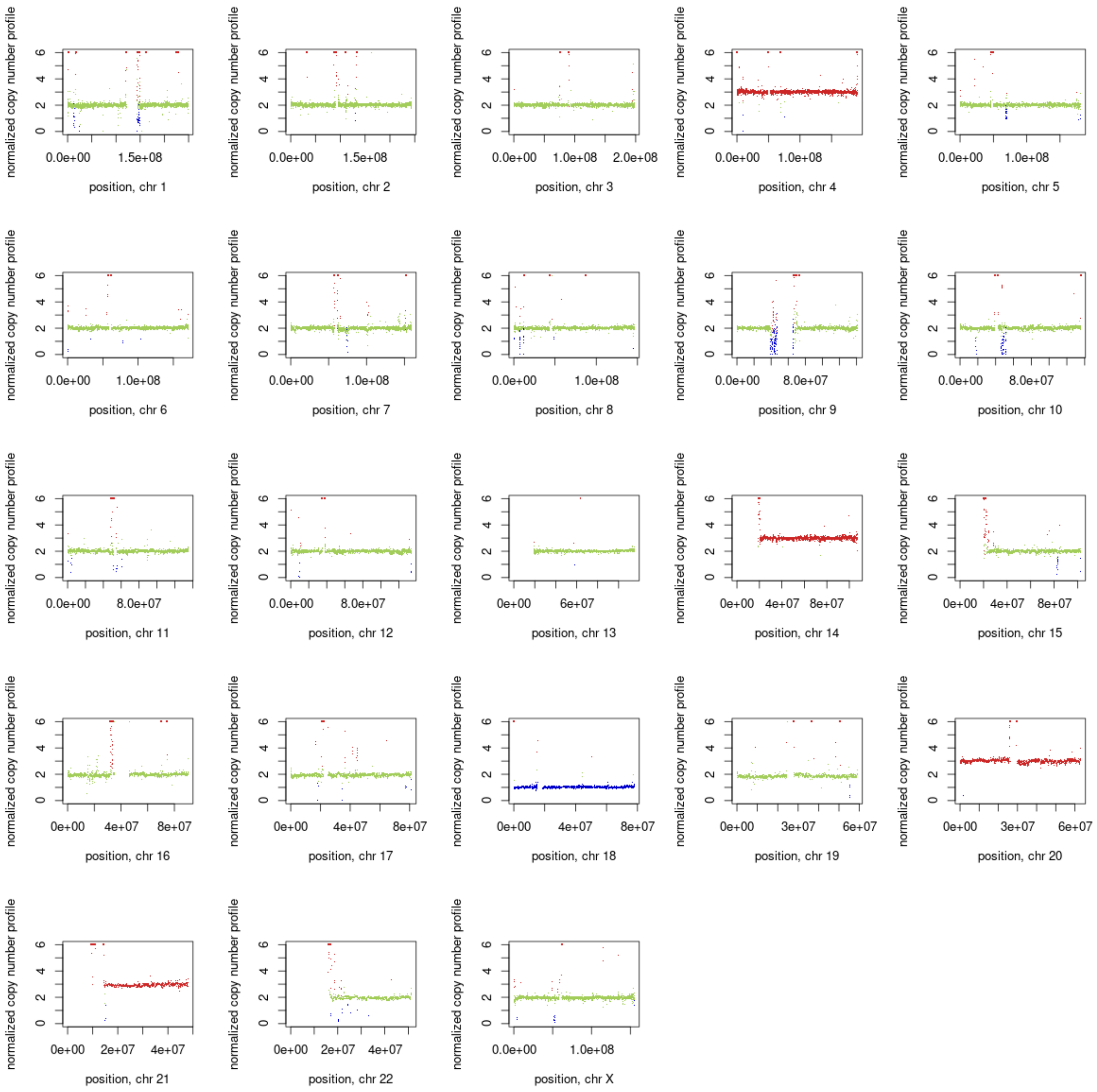
normalized copy number profile
position, chr 1
position, chr 2
position, chr 3
position, chr 4
position, chr 5
position, chr 6
position, chr 7
position, chr 8
position, chr 9
position, chr 10
position, chr 11
position, chr 12
position, chr 13
position, chr 14
position, chr 15
position, chr 16
position, chr 17
position, chr 18
position, chr 19
position, chr 20
position, chr 21
position, chr 22
position, chr X

20P

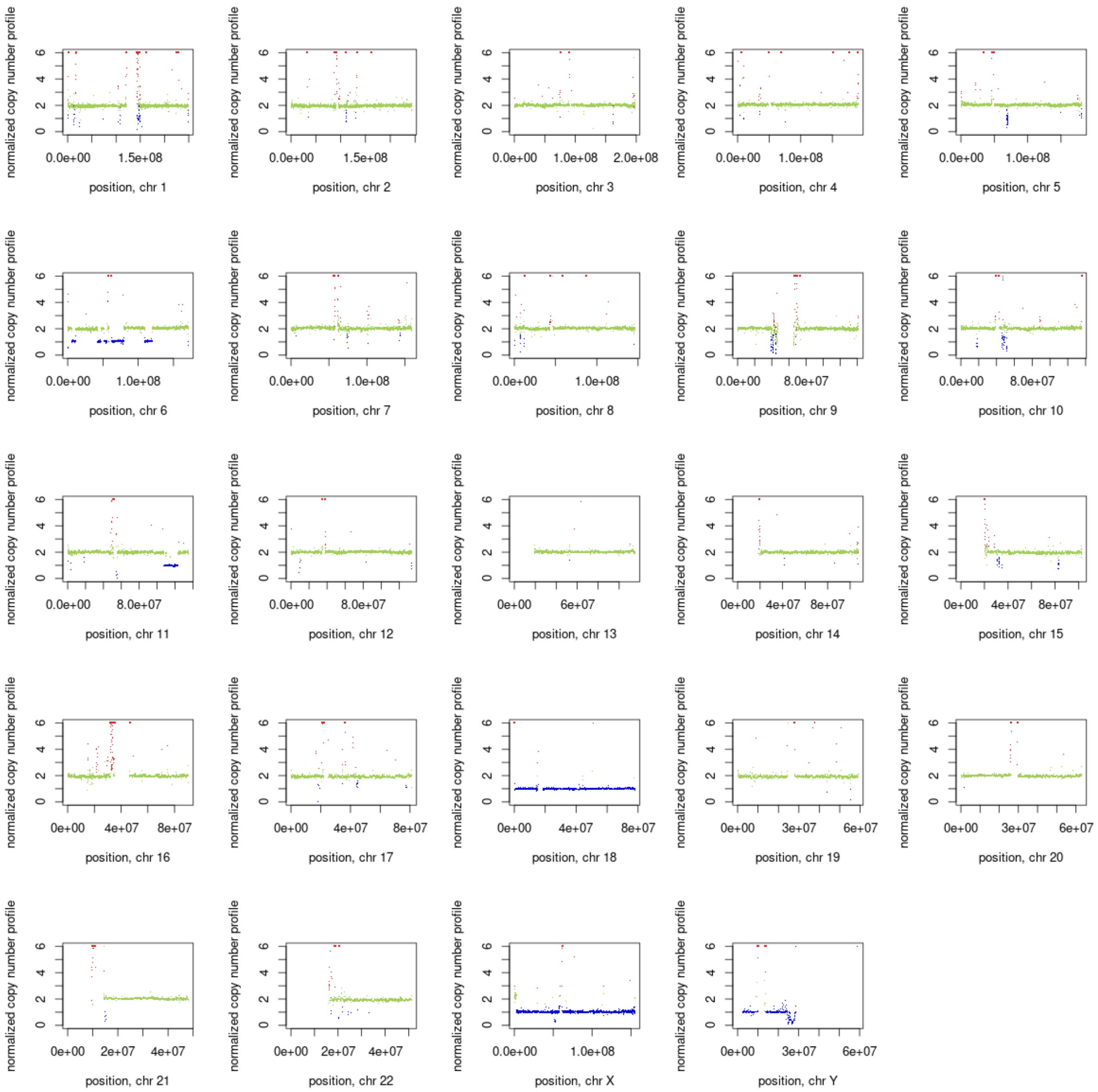
normalized copy number profile
position, chr 1
position, chr 2
position, chr 3
position, chr 4
position, chr 5
position, chr 6
position, chr 7
position, chr 8
position, chr 9
position, chr 10
position, chr 11
position, chr 12
position, chr 13
position, chr 14
position, chr 15
position, chr 16
position, chr 17
position, chr 18
position, chr 19
position, chr 20
position, chr 21
position, chr 22
position, chr X
position, chr Y

13P

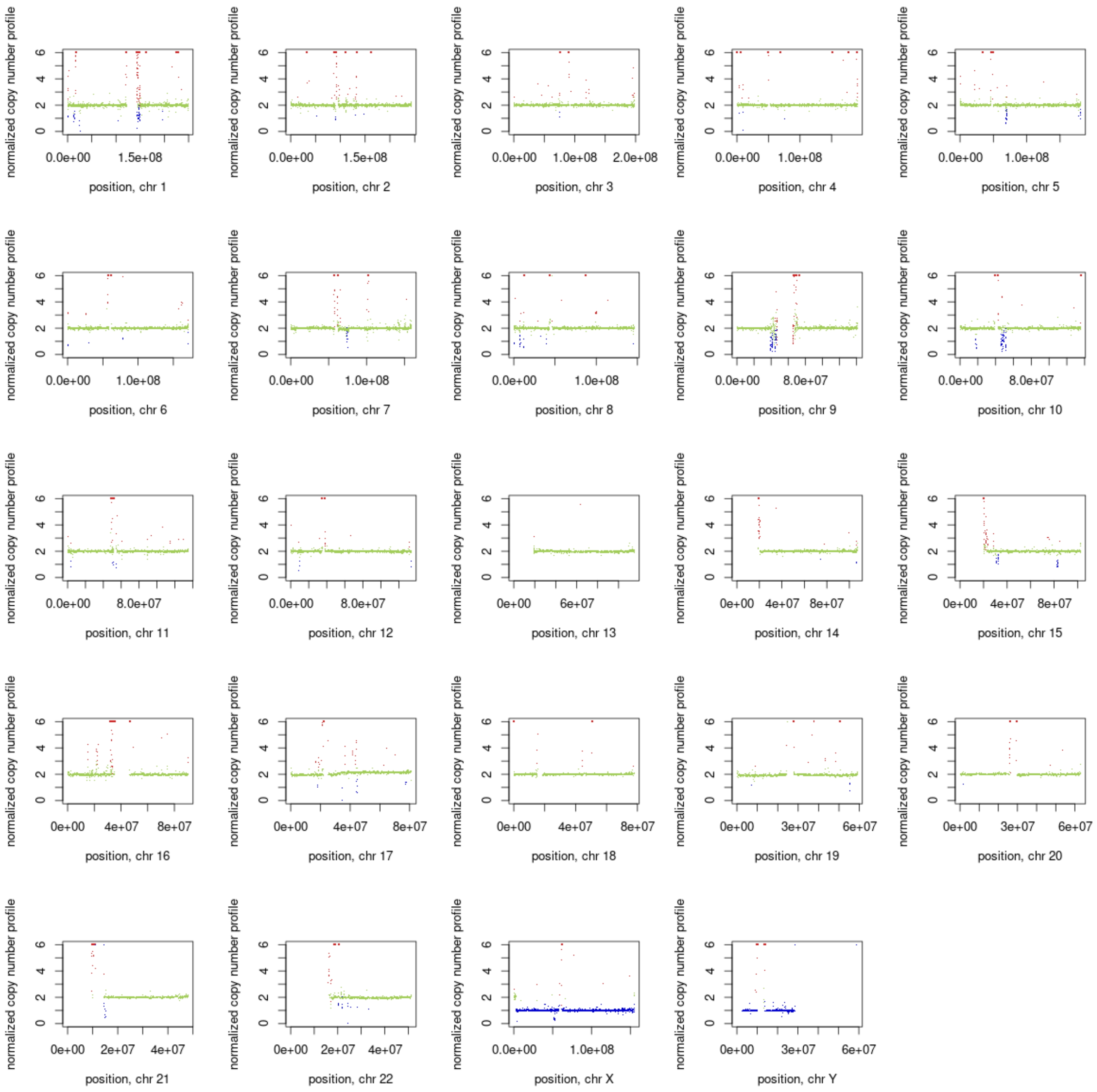
normalized copy number profile
0
2
4
6
0.0e+00 1.5e+08
position, chr 1
0.0e+00 1.5e+08
position, chr 2
0.0e+00 1.0e+08 2.0e+08
position, chr 3
0.0e+00 1.0e+08
position, chr 4
0.0e+00 1.0e+08
position, chr 5
0.0e+00 1.0e+08
position, chr 6
0.0e+00 1.0e+08
position, chr 7
0.0e+00 1.0e+08
position, chr 8
0.0e+00 8.0e+07
position, chr 9
0.0e+00 8.0e+07
position, chr 10
0.0e+00 8.0e+07
position, chr 11
0.0e+00 8.0e+07
position, chr 12
0e+00 6e+07
position, chr 13
0e+00 4e+07 8e+07
position, chr 14
0e+00 4e+07 8e+07
position, chr 15
0e+00 4e+07 8e+07
position, chr 16
0e+00 4e+07 8e+07
position, chr 17
0e+00 4e+07 8e+07
position, chr 18
0e+00 3e+07 6e+07
position, chr 19
0e+00 3e+07 6e+07
position, chr 20
0e+00 2e+07 4e+07
position, chr 21
0e+00 2e+07 4e+07
position, chr 22
0.0e+00 1.0e+08
position, chr X
0e+00 3e+07 6e+07
position, chr Y

8P

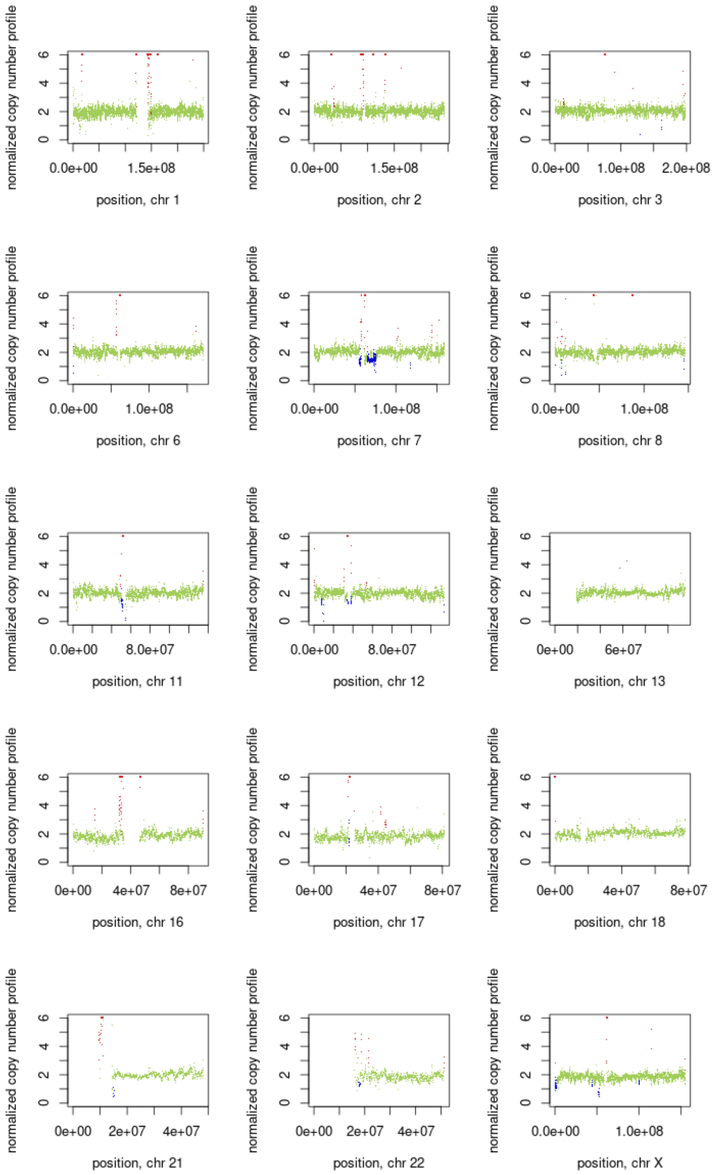
normalized copy number profile
position, chr 1
position, chr 2
position, chr 3
position, chr 4
position, chr 5
position, chr 6
position, chr 7
position, chr 8
position, chr 9
position, chr 10
position, chr 11
position, chr 12
position, chr 13
position, chr 14
position, chr 15
position, chr 16
position, chr 17
position, chr 18
position, chr 19
position, chr 20
position, chr 21
position, chr 22
position, chr X

9M

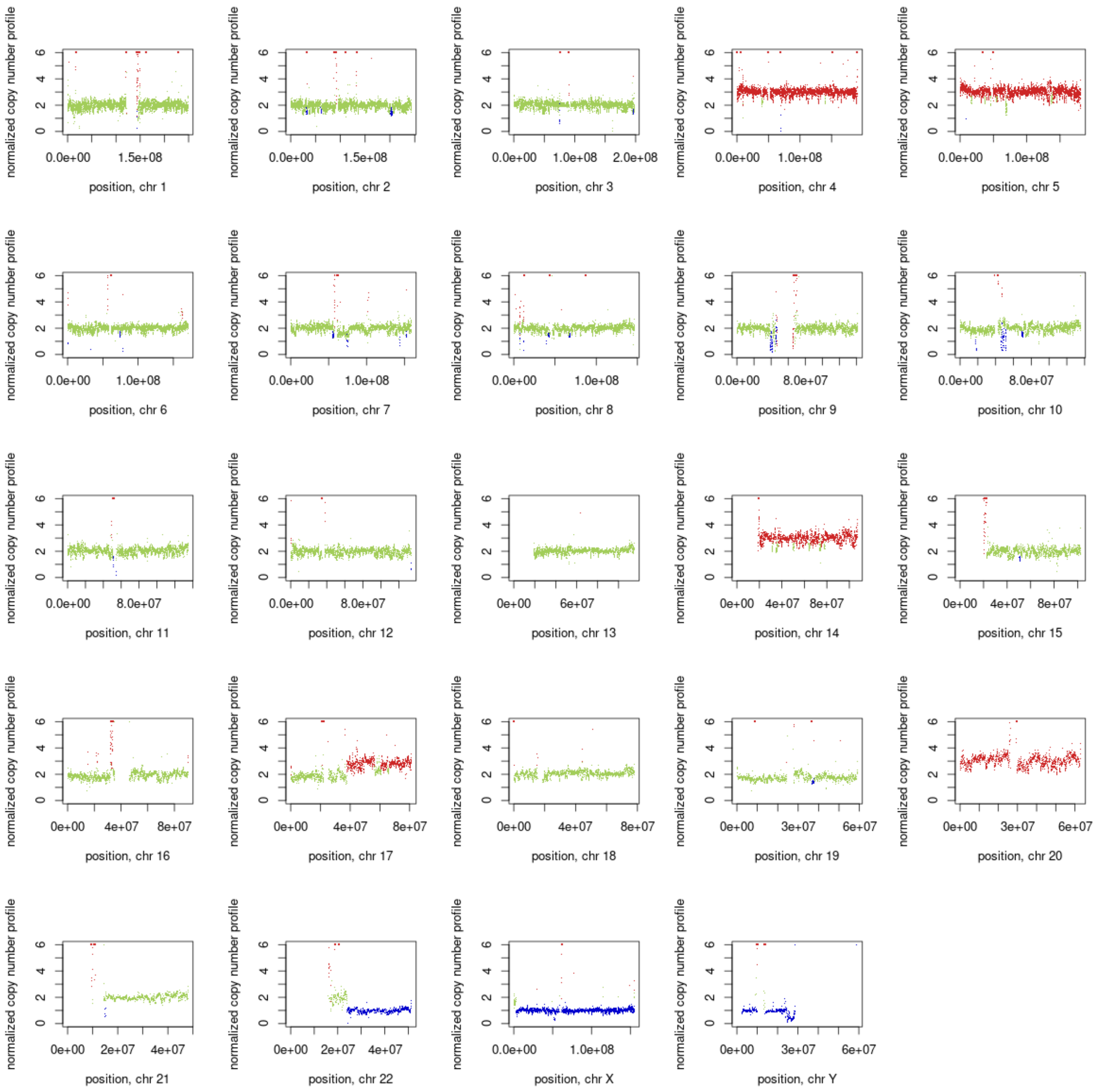
normalized copy number profile
0
2
4
6
0.0e+00 1.5e+08
position, chr 1
0.0e+00 1.5e+08
position, chr 2
0.0e+00 1.0e+08 2.0e+08
position, chr 3
0.0e+00 1.0e+08
position, chr 4
0.0e+00 1.0e+08
position, chr 5
0.0e+00 1.0e+08
position, chr 6
0.0e+00 1.0e+08
position, chr 7
0.0e+00 1.0e+08
position, chr 8
0.0e+00 8.0e+07
position, chr 9
0.0e+00 8.0e+07
position, chr 10
0.0e+00 8.0e+07
position, chr 11
0.0e+00 8.0e+07
position, chr 12
0e+00 6e+07
position, chr 13
0e+00 4e+07 8e+07
position, chr 14
0e+00 4e+07 8e+07
position, chr 15
0e+00 4e+07 8e+07
position, chr 16
0e+00 4e+07 8e+07
position, chr 17
0e+00 4e+07 8e+07
position, chr 18
0e+00 3e+07 6e+07
position, chr 19
0e+00 3e+07 6e+07
position, chr 20
0e+00 2e+07 4e+07
position, chr 21
0e+00 2e+07 4e+07
position, chr 22
0.0e+00 1.0e+08
position, chr X
0e+00 3e+07 6e+07
position, chr Y

22P

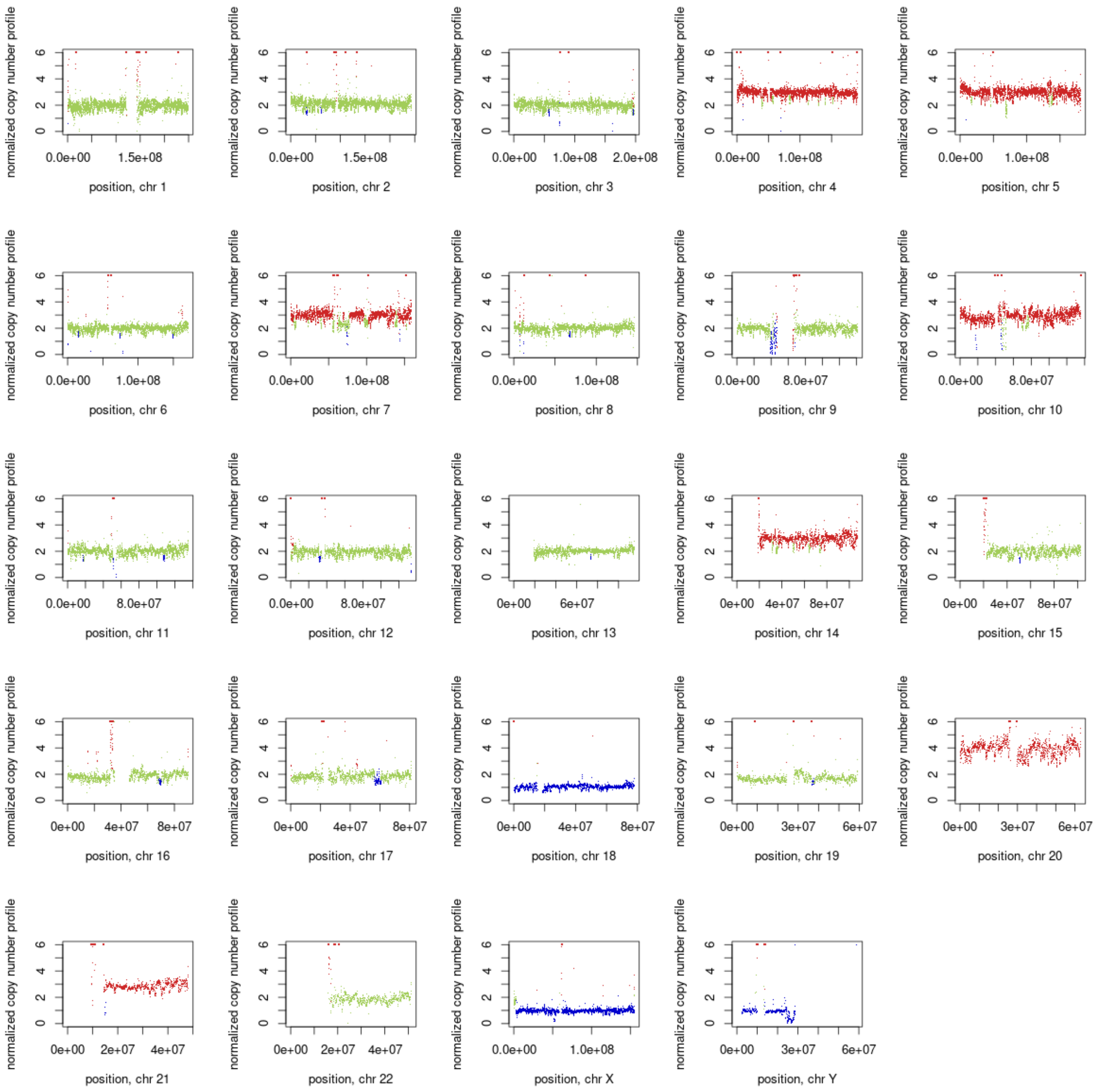
normalized copy number profile
0
2
4
6
0.0e+00 1.5e+08
position, chr 1
0.0e+00 1.5e+08
position, chr 2
0.0e+00 1.0e+08 2.0e+08
position, chr 3
0.0e+00 1.0e+08
position, chr 4
0.0e+00 1.0e+08
position, chr 5
0.0e+00 1.0e+08
position, chr 6
0.0e+00 1.0e+08
position, chr 7
0.0e+00 1.0e+08
position, chr 8
0.0e+00 8.0e+07
position, chr 9
0.0e+00 8.0e+07
position, chr 10
0.0e+00 8.0e+07
position, chr 11
0.0e+00 8.0e+07
position, chr 12
0e+00 6e+07
position, chr 13
0e+00 4e+07 8e+07
position, chr 14
0e+00 4e+07 8e+07
position, chr 15
0e+00 4e+07 8e+07
position, chr 16
0e+00 4e+07 8e+07
position, chr 17
0e+00 4e+07 8e+07
position, chr 18
0e+00 3e+07 6e+07
position, chr 19
0e+00 3e+07 6e+07
position, chr 20
0e+00 2e+07 4e+07
position, chr 21
0e+00 2e+07 4e+07
position, chr 22
0.0e+00 1.0e+08
position, chr X
0e+00 3e+07 6e+07
position, chr Y

22M

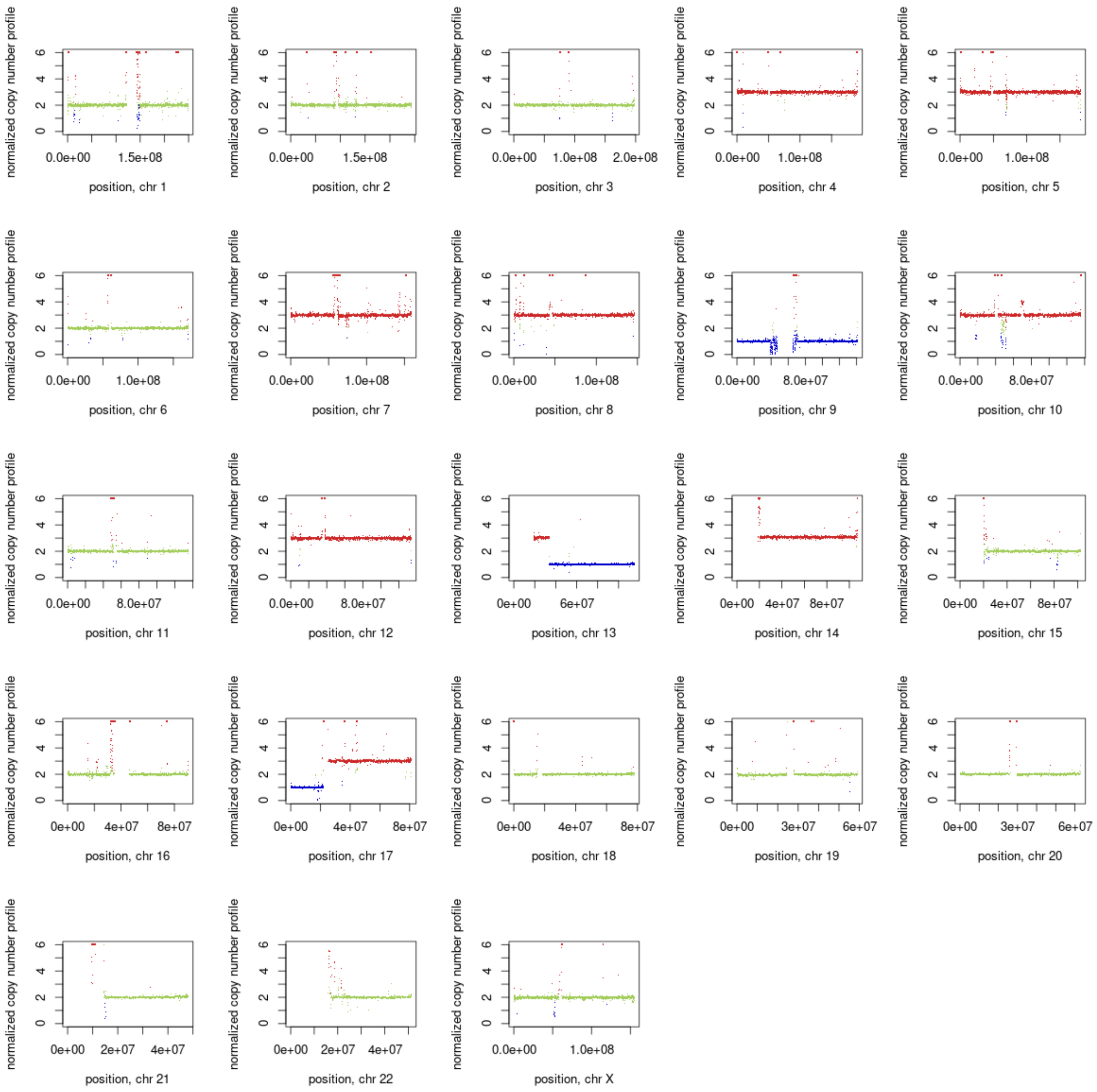
normalized copy number profile
0
2
4
6
0.0e+00
1.5e+08
position, chr 1
position, chr 2
1.0e+08
2.0e+08
position, chr 3
position, chr 4
position, chr 5
position, chr 6
position, chr 7
position, chr 8
8.0e+07
position, chr 9
position, chr 10
position, chr 11
position, chr 12
0e+00
6e+07
position, chr 13
4e+07
8e+07
position, chr 14
position, chr 15
position, chr 16
position, chr 17
position, chr 18
3e+07
6e+07
position, chr 19
position, chr 20
2e+07
position, chr 21
position, chr 22
position, chr X

1M

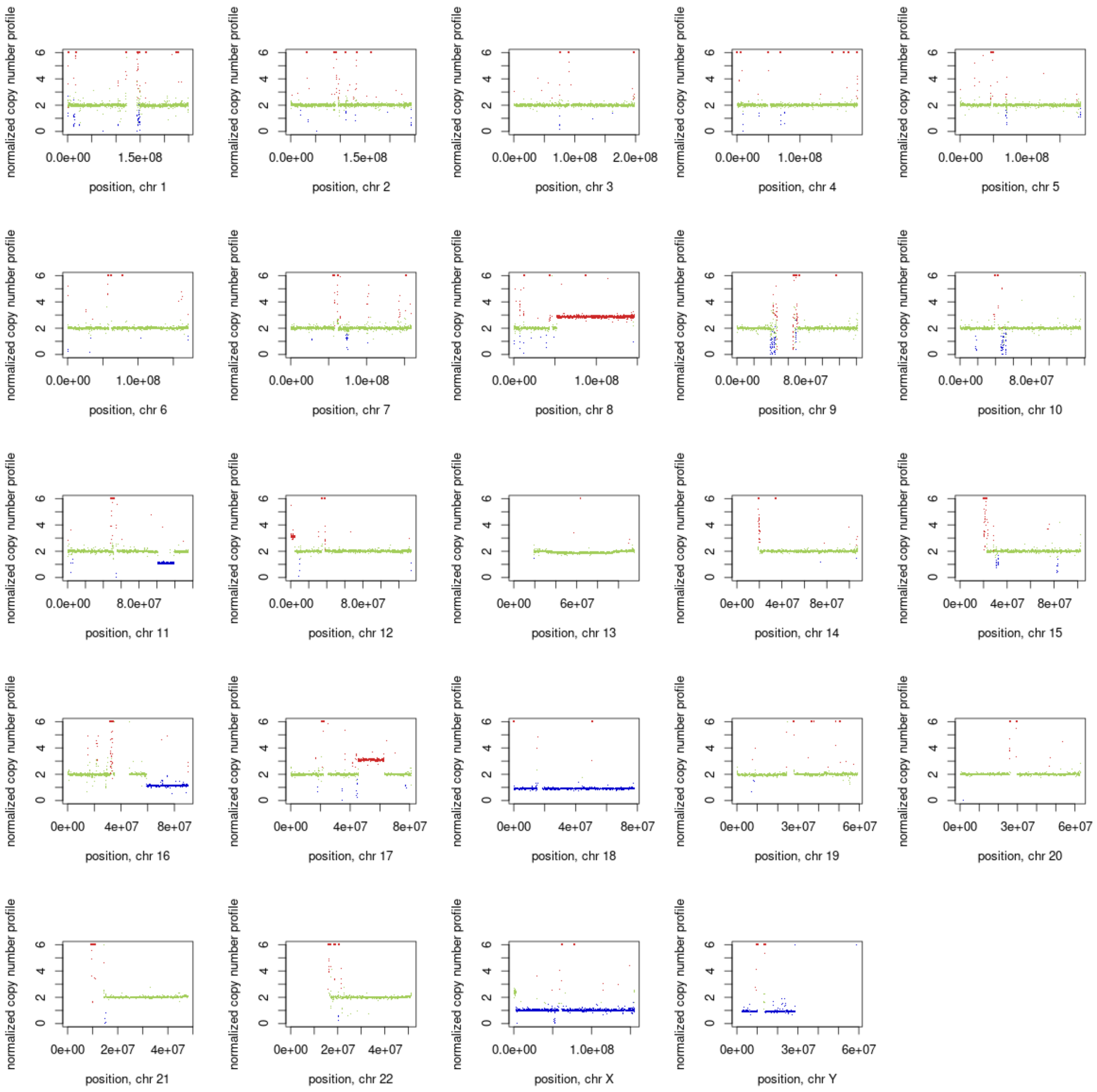
normalized copy number profile
position, chr 1
position, chr 2
position, chr 3
position, chr 4
position, chr 5
position, chr 6
position, chr 7
position, chr 8
position, chr 9
position, chr 10
position, chr 11
position, chr 12
position, chr 13
position, chr 14
position, chr 15
position, chr 16
position, chr 17
position, chr 18
position, chr 19
position, chr 20
position, chr 21
position, chr 22
position, chr X
position, chr Y

11P

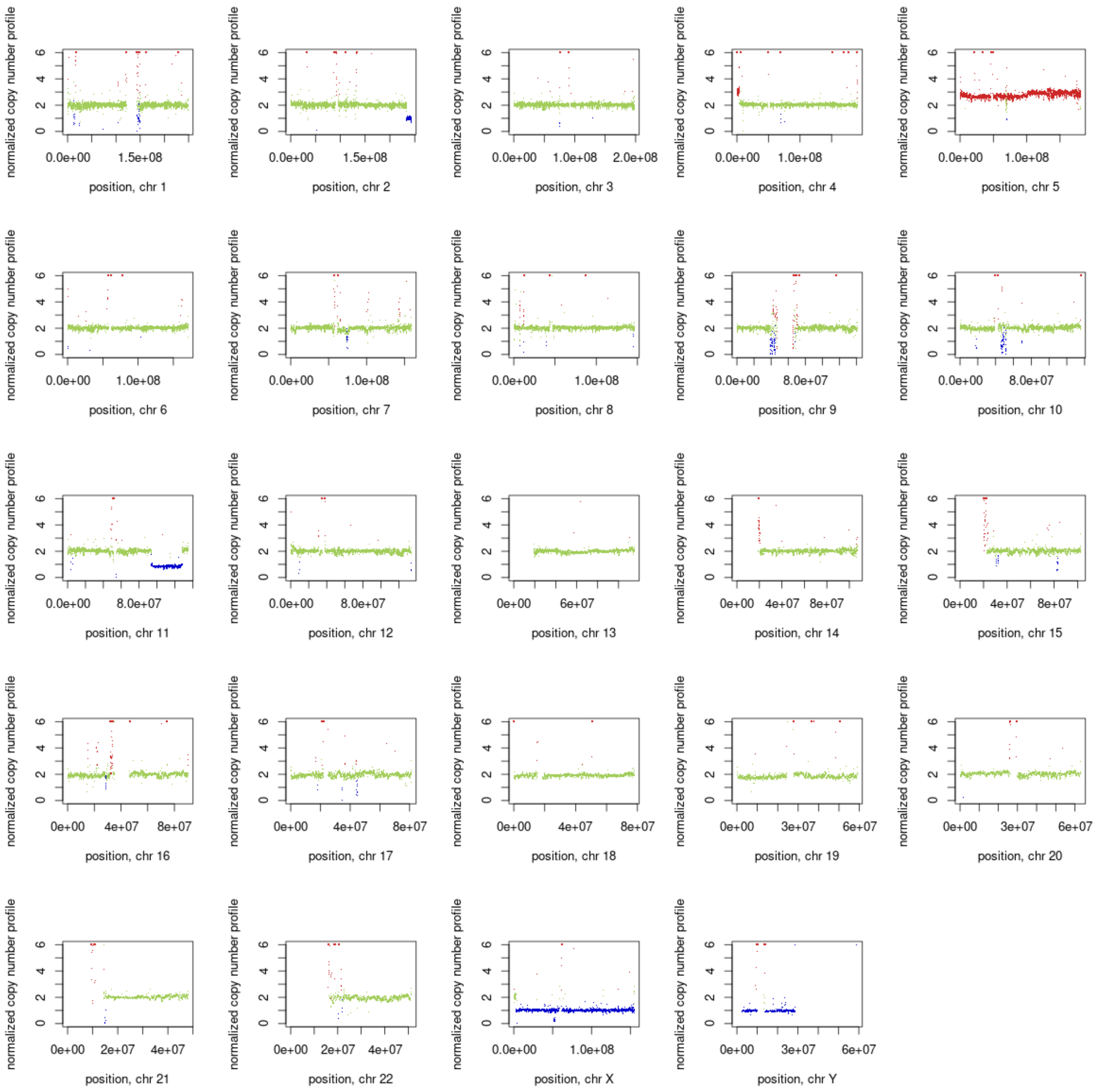
normalized copy number profile
0 2 4 6
0.0e+00 1.5e+08
position, chr 1
0.0e+00 1.5e+08
position, chr 2
0.0e+00 1.0e+08 2.0e+08
position, chr 3
0.0e+00 1.0e+08
position, chr 4
0.0e+00 1.0e+08
position, chr 5
0.0e+00 1.0e+08
position, chr 6
0.0e+00 1.0e+08
position, chr 7
0.0e+00 1.0e+08
position, chr 8
0.0e+00 8.0e+07
position, chr 9
0.0e+00 8.0e+07
position, chr 10
0.0e+00 8.0e+07
position, chr 11
0.0e+00 8.0e+07
position, chr 12
0e+00 6e+07
position, chr 13
0e+00 4e+07 8e+07
position, chr 14
0e+00 4e+07 8e+07
position, chr 15
0e+00 4e+07 8e+07
position, chr 16
0e+00 4e+07 8e+07
position, chr 17
0e+00 4e+07 8e+07
position, chr 18
0e+00 3e+07 6e+07
position, chr 19
0e+00 3e+07 6e+07
position, chr 20
0e+00 2e+07 4e+07
position, chr 21
0e+00 2e+07 4e+07
position, chr 22
0.0e+00 1.0e+08
position, chr X
0e+00 3e+07 6e+07
position, chr Y

11M

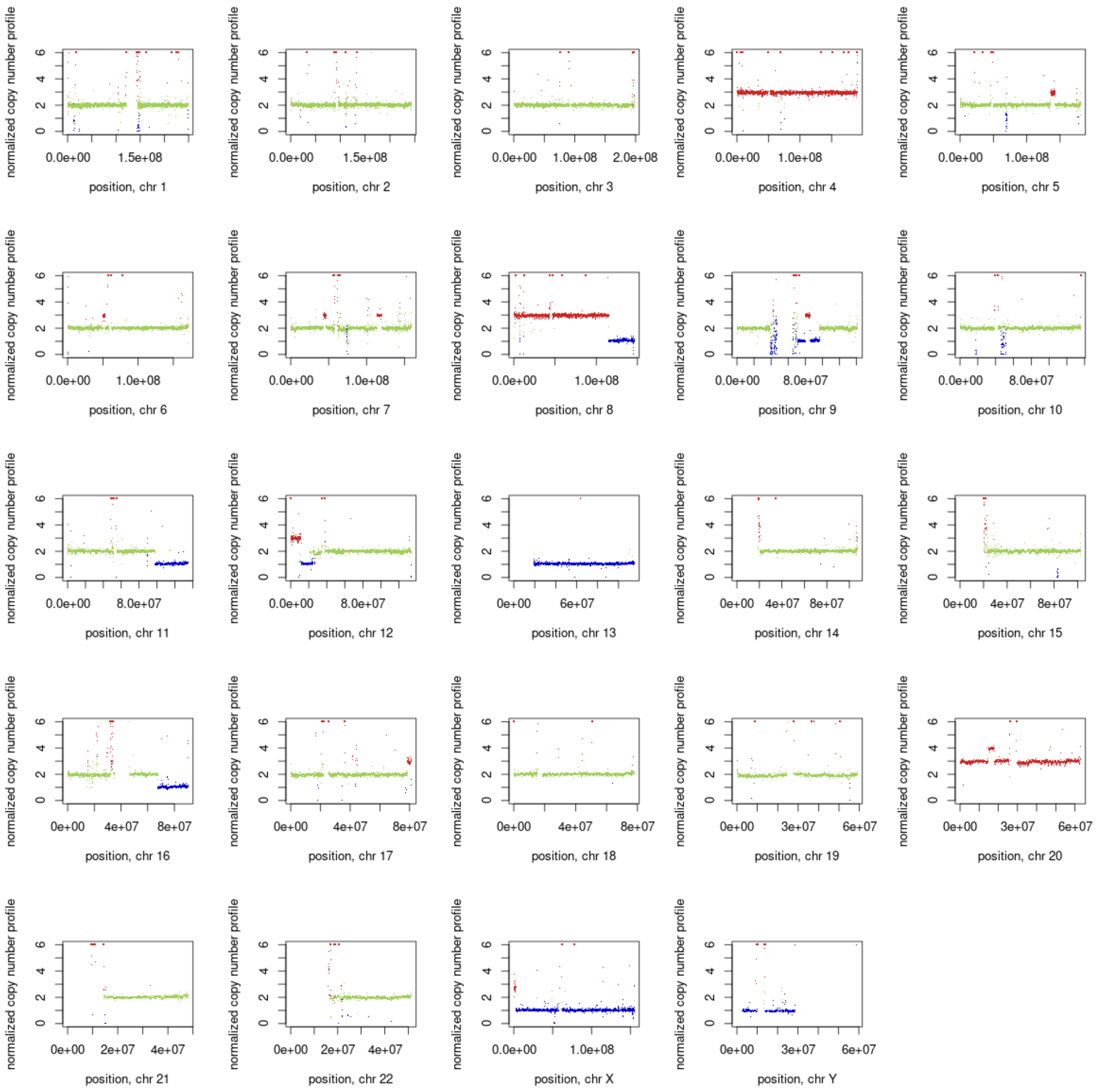
normalized copy number profile
0
2
4
6
0.0e+00 1.5e+08
position, chr 1
0.0e+00 1.5e+08
position, chr 2
0.0e+00 1.0e+08 2.0e+08
position, chr 3
0.0e+00 1.0e+08
position, chr 4
0.0e+00 1.0e+08
position, chr 5
0.0e+00 1.0e+08
position, chr 6
0.0e+00 1.0e+08
position, chr 7
0.0e+00 1.0e+08
position, chr 8
0.0e+00 8.0e+07
position, chr 9
0.0e+00 8.0e+07
position, chr 10
0.0e+00 8.0e+07
position, chr 11
0.0e+00 8.0e+07
position, chr 12
0e+00 6e+07
position, chr 13
0e+00 4e+07 8e+07
position, chr 14
0e+00 4e+07 8e+07
position, chr 15
0e+00 4e+07 8e+07
position, chr 16
0e+00 4e+07 8e+07
position, chr 17
0e+00 4e+07 8e+07
position, chr 18
0e+00 3e+07 6e+07
position, chr 19
0e+00 3e+07 6e+07
position, chr 20
0e+00 2e+07 4e+07
position, chr 21
0e+00 2e+07 4e+07
position, chr 22
0.0e+00 1.0e+08
position, chr X
0e+00 3e+07 6e+07
position, chr Y

4M

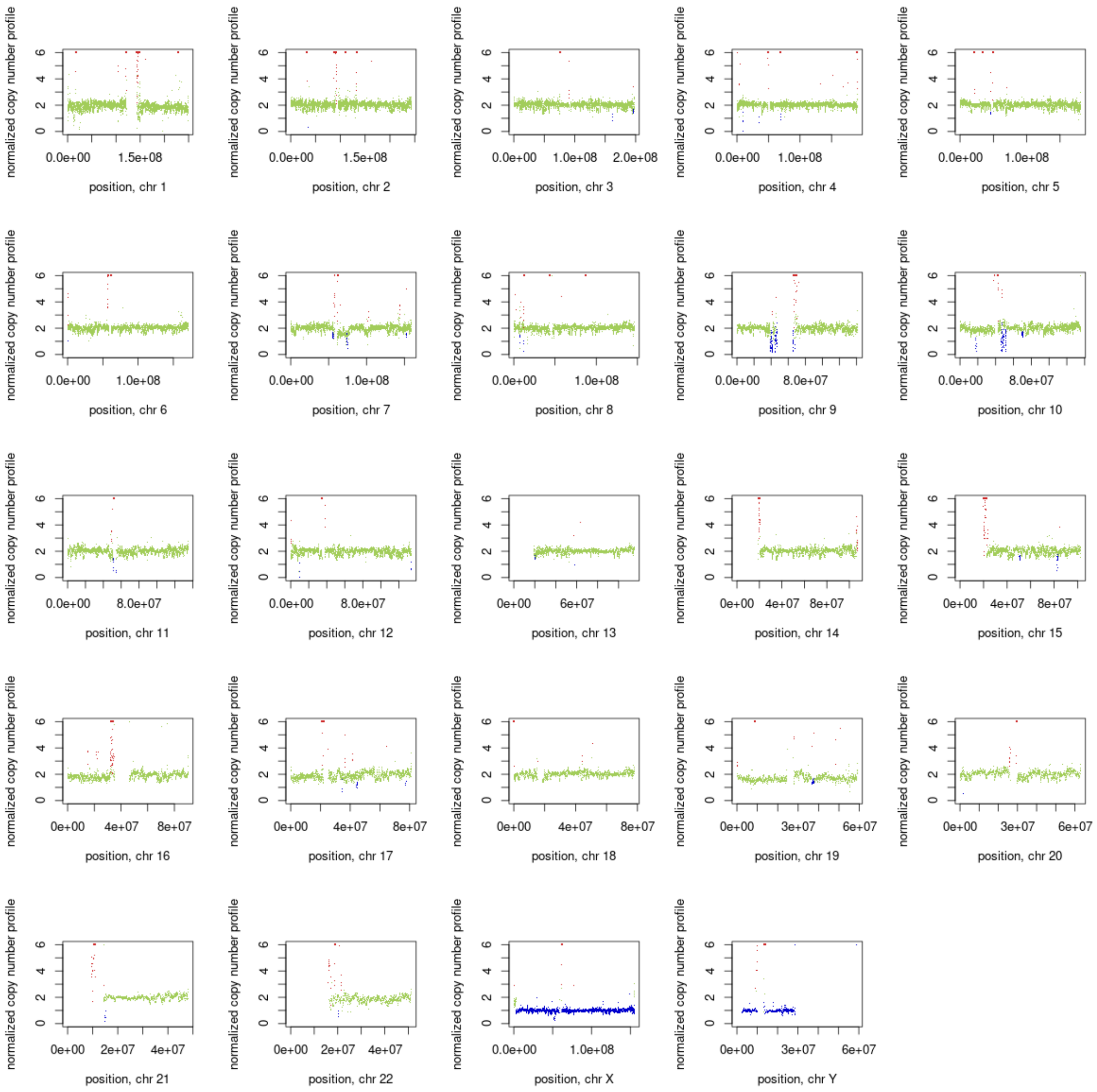
normalized copy number profile
0 2 4 6
0.0e+00 1.5e+08
position, chr 1
normalized copy number profile
0 2 4 6
0.0e+00 1.5e+08
position, chr 2
normalized copy number profile
0 2 4 6
0.0e+00 1.0e+08 2.0e+08
position, chr 3
normalized copy number profile
0 2 4 6
0.0e+00 1.0e+08
position, chr 4
normalized copy number profile
0 2 4 6
0.0e+00 1.0e+08
position, chr 5
normalized copy number profile
0 2 4 6
0.0e+00 1.0e+08
position, chr 6
normalized copy number profile
0 2 4 6
0.0e+00 1.0e+08
position, chr 7
normalized copy number profile
0 2 4 6
0.0e+00 1.0e+08
position, chr 8
normalized copy number profile
0 2 4 6
0.0e+00 8.0e+07
position, chr 9
normalized copy number profile
0 2 4 6
0.0e+00 8.0e+07
position, chr 10
normalized copy number profile
0 2 4 6
0.0e+00 8.0e+07
position, chr 11
normalized copy number profile
0 2 4 6
0.0e+00 8.0e+07
position, chr 12
normalized copy number profile
0 2 4 6
0e+00 6e+07
position, chr 13
normalized copy number profile
0 2 4 6
0e+00 4e+07 8e+07
position, chr 14
normalized copy number profile
0 2 4 6
0e+00 4e+07 8e+07
position, chr 15
normalized copy number profile
0 2 4 6
0e+00 4e+07 8e+07
position, chr 16
normalized copy number profile
0 2 4 6
0e+00 4e+07 8e+07
position, chr 17
normalized copy number profile
0 2 4 6
0e+00 4e+07 8e+07
position, chr 18
normalized copy number profile
0 2 4 6
0e+00 3e+07 6e+07
position, chr 19
normalized copy number profile
0 2 4 6
0e+00 3e+07 6e+07
position, chr 20
normalized copy number profile
0 2 4 6
0e+00 2e+07 4e+07
position, chr 21
normalized copy number profile
0 2 4 6
0e+00 2e+07 4e+07
position, chr 22
normalized copy number profile
0 2 4 6
0.0e+00 1.0e+08
position, chr X
normalized copy number profile
0 2 4 6
0e+00 3e+07 6e+07
position, chr Y

19M

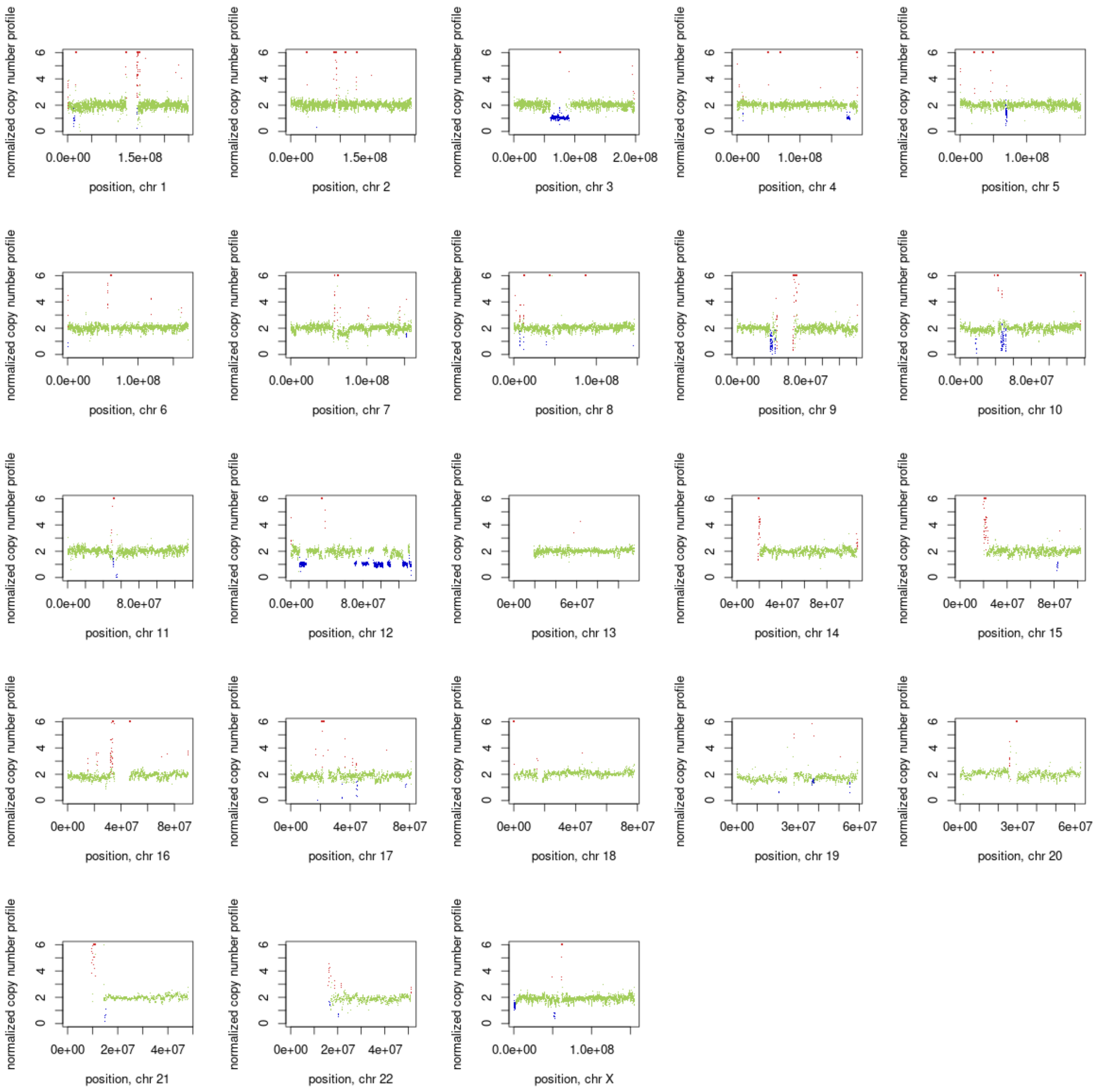
normalized copy number profile
0
2
4
6
position, chr 1
position, chr 2
position, chr 3
position, chr 4
position, chr 5
position, chr 6
position, chr 7
position, chr 8
position, chr 9
position, chr 10
position, chr 11
position, chr 12
position, chr 13
position, chr 14
position, chr 15
position, chr 16
position, chr 17
position, chr 18
position, chr 19
position, chr 20
position, chr 21
position, chr 22
position, chr X
0.0e+00
1.5e+08
1.0e+08
2.0e+08
8.0e+07
0e+00
6e+07
4e+07
8e+07
3e+07
2e+07

25P
